# Pharmacometric assessment of the *in vivo* antiviral activity of ivermectin in early symptomatic COVID-19

**DOI:** 10.1101/2022.07.15.22277570

**Authors:** William HK Schilling, Podjanee Jittamala, James A Watson, Maneerat Ekkapongpisit, Tanaya Siripoon, Thundon Ngamprasertchai, Viravarn Luvira, Sasithon Pongwilai, Cintia Cruz, James J Callery, Simon Boyd, Varaporn Kruabkontho, Thatsanun Ngernseng, Jaruwan Tubprasert, Mohammad Yazid Abdad, Nattaporn Piaraksa, Kanokon Suwannasin, Pongtorn Hanboonkunupakarn, Borimas Hanboonkunupakarn, Sakol Sookprome, Kittiyod Poovorawan, Janjira Thaipadungpanit, Stuart Blacksell, Mallika Imwong, Joel Tarning, Walter RJ Taylor, Vasin Chotivanich, Chunlanee Sangketchon, Wiroj Ruksakul, Kesinee Chotivanich, Mauro M Teixeira, Sasithon Pukrittayakamee, Arjen M Dondorp, Nicholas PJ Day, Watcharapong Piyaphanee, Weerapong Phumratanaprapin, Nicholas J White, the PLATCOV Collaborative Group

## Abstract

**Background:** There is no generally accepted methodology for *in vivo* assessment of antiviral activity in SARS-CoV-2 infection. Ivermectin has been recommended widely as a treatment of COVID-19, but whether it has significant antiviral activity *in vivo* is uncertain.

**Methods:** In a multicentre open label, randomized, controlled adaptive platform trial, adult patients with early symptomatic COVID-19 were randomized to one of six treatment arms including high dose ivermectin (600µg/kg daily for seven days), the monoclonal antibodies casirivimab and imdevimab (600mg/600mg), and no study drug. Viral clearance rates were derived from daily duplicate oropharyngeal quantitative PCR measurements. This ongoing trial is registered at ClinicalTrials.gov (NCT05041907).

**Results:** Randomization to the ivermectin arm was stopped after enrolling 205 patients into all arms, as the prespecified futility threshold was reached. Compared with the no study drug arm, the mean estimated SARS-CoV-2 viral clearance following ivermectin was 9.1% slower [95%CI -27.2% to +11.8%; n=45 versus n=41], whereas in a preliminary analysis of the casirivimab/imdevimab arm it was 52.3% faster [95%CI +7.0% to +115.1%; n=10 (Delta variant) versus n=41].

**Conclusions:** High dose ivermectin did not have measurable antiviral activity in early symptomatic COVID-19. Measured in this way viral clearance rate is a valuable pharmacodynamic measure in assessing antiviral COVID-19 therapeutics *in vivo*.

**Funding:** “Finding treatments for COVID-19: A phase 2 multi-centre adaptive platform trial to assess antiviral pharmacodynamics in early symptomatic COVID-19 (PLAT-COV)” is funded by the Wellcome Therapeutics Accelerator (223195/Z/21/Z).

**Impact:**

- Rate of viral clearance determined from daily duplicate oropharyngeal swabs over one week is an efficient measure of antiviral efficacy in early COVID-19 infection.
- High dose ivermectin did not demonstrate measurable antiviral activity in early symptomatic COVID-19 infection.

## Introduction

Effective, well-tolerated and inexpensive oral antiviral agents are needed for the early treatment of COVID-19. Monoclonal antibodies, mainly directed against the SARS-CoV-2 spike protein, have proved effective in preventing and treating COVID-19 (1, 2), but they are expensive, require parenteral administration, and are vulnerable to the emergence of spike protein mutations (3). Recently, large randomized controlled trials have shown clinical efficacy for the ribonucleoside analogue molnupiravir and the protease inhibitor nirmatrelvir (in combination with ritonavir) in the treatment of early COVID-19 (4, 5), but these drugs are not yet widely available, especially in low- and middle-Income (LMIC) settings. There have been no reported randomized comparisons between these expensive medicines. In the absence of comparative assessments, and uncertainty over antiviral efficacy, national treatment guidelines vary widely across the world.

Early in the COVID-19 pandemic considerable attention was focused on available drugs that might have useful antiviral activity (6). Notable and widely promoted repurposing candidates included hydroxychloroquine, remdesivir and ivermectin. The macrocyclic lactone endectocide ivermectin was pursued after a laboratory study suggested antiviral activity against SARS-CoV-2 (7). This *in vitro* activity, extensive experience in mass treatments, a well-established safety profile, and claims of clinical benefit, led to ivermectin being added to COVID-19 treatment guidelines in many countries, particularly in Latin America (8). Several small clinical trials have reported a survival benefit for ivermectin, although the quality of these trials has been questioned (9). Ivermectin’s relatively weak *in vitro* activity in relation to achievable levels *in vivo* has argued for the evaluation of maximum tolerated doses (c.600 µg/kg/day). Although the large TOGETHER platform trial showed no significant benefit with ivermectin in early COVID-19 infection on a composite outcome of hospitalization or lengthy (>six hours) emergency department visit, it used a lower dose of 400 µg/kg/day for only three days (10).

To resolve the uncertainty over efficacy, we measured the *in vivo* antiviral activity of high dose ivermectin in previously healthy adults with early symptomatic COVID-19 infection.

## Methods

PLATCOV is an ongoing open label, randomized, controlled adaptive platform trial designed to provide a standardized quantitative comparative method for *in vivo* assessment of potential antiviral treatments in early symptomatic COVID-19. The primary outcome is the rate of viral clearance measured as the change in the slope of the log_10_ oropharyngeal viral clearance curve (11). The trial was conducted in the Hospital for Tropical Diseases, Faculty of Tropical Medicine, Mahidol University, Bangkok, Thailand; Bangplee hospital, Samut Prakarn, Thailand; and Vajira hospital, Navamindradhiraj University, Bangkok, Thailand (full details in Supplementary materials). The trial was approved by local and national research ethics boards in Thailand (Faculty of Tropical Medicine Ethics Committee, Mahidol University (FTMEC Bangkok, Thailand, FTMEC Ref: TMEC 21-058) and the Central Research Ethics Committee (CREC, Bangkok, Thailand, CREC Ref: CREC048/64BP-MED34) and by the Oxford University Tropical Research Ethics Committee (OxTREC, Oxford, UK, OxTREC Ref: 24-21). All the patients provided fully informed written consent. The trial was coordinated and monitored by the Mahidol Oxford Tropical Medicine Research Unit (MORU), who conducted the trial, collected the data, and had full access to all the trial data and confirm the performance of the trial according to the protocol. The PLATCOV trial was overseen by a trial steering committee (TSC) and results were reviewed by a data and safety monitoring board (DSMB). The funders had no role in the design and conduct of the trial, data collection, management, nor analysis, neither the preparation, review, nor approval of the manuscript, nor the decision to submit the manuscript for publication. The trial is ongoing and is registered at ClinicalTrials.gov (NCT05041907).

### Participants

Patients presenting to the Acute Respiratory Infections (ARI) outpatient clinics for COVID-19 testing were pre-screened for study eligibility. Previously healthy adults aged between 18 and 50 years were eligible for the trial if they had early symptomatic COVID-19 (i.e. reported symptoms for not more than 4 days), oxygen saturation ≥96%, were unimpeded in activities of daily living, and were willing to give fully informed consent and adhere to the study protocol. SARS-CoV-2 positivity was defined either as a nasal lateral flow antigen test which became positive within two minutes (STANDARD™ Q COVID-19 Ag Test, SD Biosensor, Suwon-si, Republic of Korea) or a positive PCR test within the previous 24hrs with a cycle threshold value (Ct) of less than 25 (all viral gene targets), both suggesting high viral loads. Exclusion criteria included taking any potential antivirals or pre-existing concomitant medications, chronic illness or significant comorbidity, haematological or biochemical abnormalities, pregnancy (a urinary pregnancy test was performed in females), breastfeeding, or contraindication or known hypersensitivity to any of the study drugs.

### Randomization and interventions

Randomization was performed via a centralized web-app designed by MORU software engineers using RShiny®, hosted on a MORU webserver, although for all sites envelopes were also generated initially as back-up. The no study drug arm comprised a minimum proportion of 20% and uniform randomization ratios were applied across the treatment arms, (e.g. if six arms were active then the randomization ratios would be 5:4:4:4:4:4). All patients received standard symptomatic treatment. Ivermectin (600 μg/kg; Atlantic Laboratories, Thailand) was given once daily for seven days with food. The tablets were 6mg each (see Supplementary materials for dosing table) and patients were supplied with a hospital meal comprising of 500-600kcal and 20-25% fat. Casirivimab/imdevimab (600mg/600mg; Roche, Switzerland) was given once by intravenous infusion at randomization. During this period, patients were also randomized to remdesivir, favipiravir and fluoxetine.

### Trial procedures

Eligible patients were admitted to the study ward where a full clinical examination was performed. Baseline investigations included a rapid SARS-CoV-2 antibody test (BIOSYNEX COVID-19 BSS IgM/IgG, Illkirch-Graffenstaden, France), blood sampling for haematology and biochemistry, an electrocardiogram and a chest radiograph (following local guidance, but not a study requirement). After randomization, oropharyngeal swabs (two swabs from each tonsil) were taken in a standard manner as follows. A Thermo Fisher MicroTest™ flocked swab was rotated against the tonsil through 360° four times and placed in Thermo Fisher M4RT™ viral transport medium (3mL). Swabs were transferred at 4-8°C, aliquoted and then frozen at -80°C within 48hrs. Swabs were taken from the left and right tonsils daily from day 0 to day 7, and then after discharge on day 14.

The QuantCheck™ SARS-CoV-2 Quantitative Test (Applied Biosystems^®^, Thermo Fisher Scientific, Waltham, United States) was used to quantitate viral load (RNA copies per mL). The TaqCheck™ SARS-CoV-2 Fast PCR Assay, a multiplexed real-time PCR method, detects the SARS-CoV-2 N-gene and S-gene as well as human RNase P in a single reaction. RNase P was used to correct for variation in the sample human cell content. The viral load was quantified against known standards using the ATCC^®^ heat-inactivated SARS-CoV-2 VR-1986HK™ strain 2019-nCoV/USA-WA1/2020. Viral variants were identified using real-time PCR genotyping with the TaqMan™ SARS-CoV-2 Mutation Panel (see Supplementary materials). Plasma ivermectin concentrations were determined on days three and seven using validated high-performance liquid chromatography linked with tandem mass spectrometry (12, 13). Adverse events were graded according to the Common Terminology Criteria for Adverse Events v.5.0 (CTCAE). Adverse event summaries were generated if the adverse event was grade 3 or higher and the adverse event was new or increased in intensity from study drug administration until the end of the follow up period. Serious adverse events were recorded separately and reported to the DSMB.

### Outcome measures, stopping rules and statistical analysis

The primary outcome measure was the rate of viral clearance, expressed as a slope coefficient and presented as a half-life. This was estimated under a Bayesian hierarchical linear model fitted to the daily log_10_ viral load measurements between days 0 and 7 (18 measurements per patient). The viral clearance rate (i.e. slope coefficient from the model fit) is inversely proportional to the clearance half-life (t_1/2_ = log_10_ 0.5/slope). The treatment effect is defined as the change (%) in the viral clearance rate relative to the no study drug arm (i.e. how much the treatment accelerates viral clearance). A 50% increase in clearance rate is thus equal to a 33% reduction in the clearance half-life. All cause hospitalization for clinical deterioration (until day 28) was a secondary endpoint. For each intervention in the study the sample size is adaptive. The stopping rules were determined using a simulation approach, based on previously modelled serial viral load data (11), such that approximately 50 patients are needed to demonstrate increases in the rate of viral clearance of ∼50%, with control of both type 1 and type 2 errors at 10%. The prespecified decision criteria for stopping a treatment arm were either a model-based probability of <0.1 that the intervention did not accelerate viral clearance by >12.5% relative to no study drug (futility) or >0.9 that it did (success). The first interim analysis (n=50) was prespecified as unblinded in order to review the methodology and stopping rules. Following this analysis, the stopping threshold was increased from 5% to 12.5% because the treatment effect of casirivimab/imdevimab against the SARS-CoV-2 Delta variant was substantially larger than had been expected. After this first interim analysis trial investigators were blinded.

All analyses were done in a modified intention-to-treat (mITT) population, comprising patients who had a minimum of 3 days follow-up data. The safety population includes all patients who received at least one dose of the allocated intervention. A series of linear and non-linear Bayesian hierarchical models were fitted to the viral quantitative PCR (qPCR) data (see Supplementary materials for the full details). For the pharmacokinetic analysis a previously developed 2-compartment disposition model with 4 transit compartments adjusted for body weight was fitted to the plasma ivermectin levels (14). Drug exposure was summarized as the area under the concentration time curve up until 72 hours (AUC_0-72_) and the maximum peak concentration (C_max_).

This report describes the ivermectin results compared with no treatment and also includes the unblinded results for the first ten patients who received casirivimab/imdevimab (recruited until 16^th^ December 2021) to illustrate the sensitivity of the method. All data analysis was done in R version 4.0.2. Model fitting was done in *stan* via the *rstan* interface. All code and data are openly accessible via the following GitHub repository: https://github.com/jwatowatson/PLATCOV-Ivermectin.

## Results

The trial began recruitment on the 30^th^ of September 2021. On 18^th^ April 2022, enrolment into the ivermectin group was stopped as the prespecified futility margin had been reached. Of the 274 patients screened (Figure 1), 224 were randomized to either ivermectin (46 patients), casirivimab/imdevimab (40 patients; the unblinded first ten are reported here), no study drug (45 patients), or other interventions (93 patients: remdesivir, favipiravir, and fluoxetine). This analysis dataset therefore comprised 101 patients (46 ivermectin, 10 casirivimab/imdevimab, 45 no study drug), of whom five were excluded for either changing treatment before day 2 (n=3), withdrawing from the study (n=1), or because there was no detectable viral RNA at all timepoints (n=1), (Figure 1). In the mITT population (n=96), 60% were female and the median age was 27 (interquartile range 25-31) years. The median duration of illness at enrolment was 2 (interquartile range 2-3) days. Overall, 95% of all patients had received at least one dose of a COVID-19 vaccine (Table 1). The median (range) daily dose of ivermectin was 550μg/kg/day (490-600μg/kg/day). All patients recovered without complications. One patient in the no study drug arm was hospitalized for clinical reasons before day 28 (see safety and tolerability section). Virus variants spanned Delta (B.1.617.2), prevalent when the study began, Omicron BA.1 (B.1.1.529), and Omicron BA.2 (B.1.1.529) (Supplementary Figure 1).

**Table 1.** Summary of patient characteristics included in the mITT population (n= 96). HTD: Hospital for Tropical Diseases; BP: Bangplee Hospital; VJ: Vajira Hospital.

| Treatment arm | Number (total n=96) | Age median (range) | Baseline viral load mean log<sub>10</sub> copies per mL (range) | Vaccine doses median (range) | Antibody positive from rapid test (%)* | Male (%) | Sites |  |  |
| --- | --- | --- | --- | --- | --- | --- | --- | --- | --- |
|  |  |  |  |  |  |  | HTD (n=87) | BP (n=5) | VJ (n=4) |
| Casirivimab/imdevimab | 10 | 26.5 (18-31) | 5.5 (3.7-7.8) | 2 (0-3) | 50 | 20 | 10 | 0 | 0 |
| Ivermectin | 45 | 29 (19-45) | 5.7 (1.9-7.6) | 2 (0-4) | 78 | 47 | 41 | 2 | 2 |
| No study drug | 41 | 27 (20-43) | 5.5 (3-7.7) | 2 (2-4) | 90 | 44 | 36 | 3 | 2 |

\*Defined as IgM or IgG present on the rapid antibody test ((BIOSYNEX COVID-19 BSS IgM/IgG, Illkirch-Graffenstaden, France) used as per manufacturer's instructions

**Figure 1.**
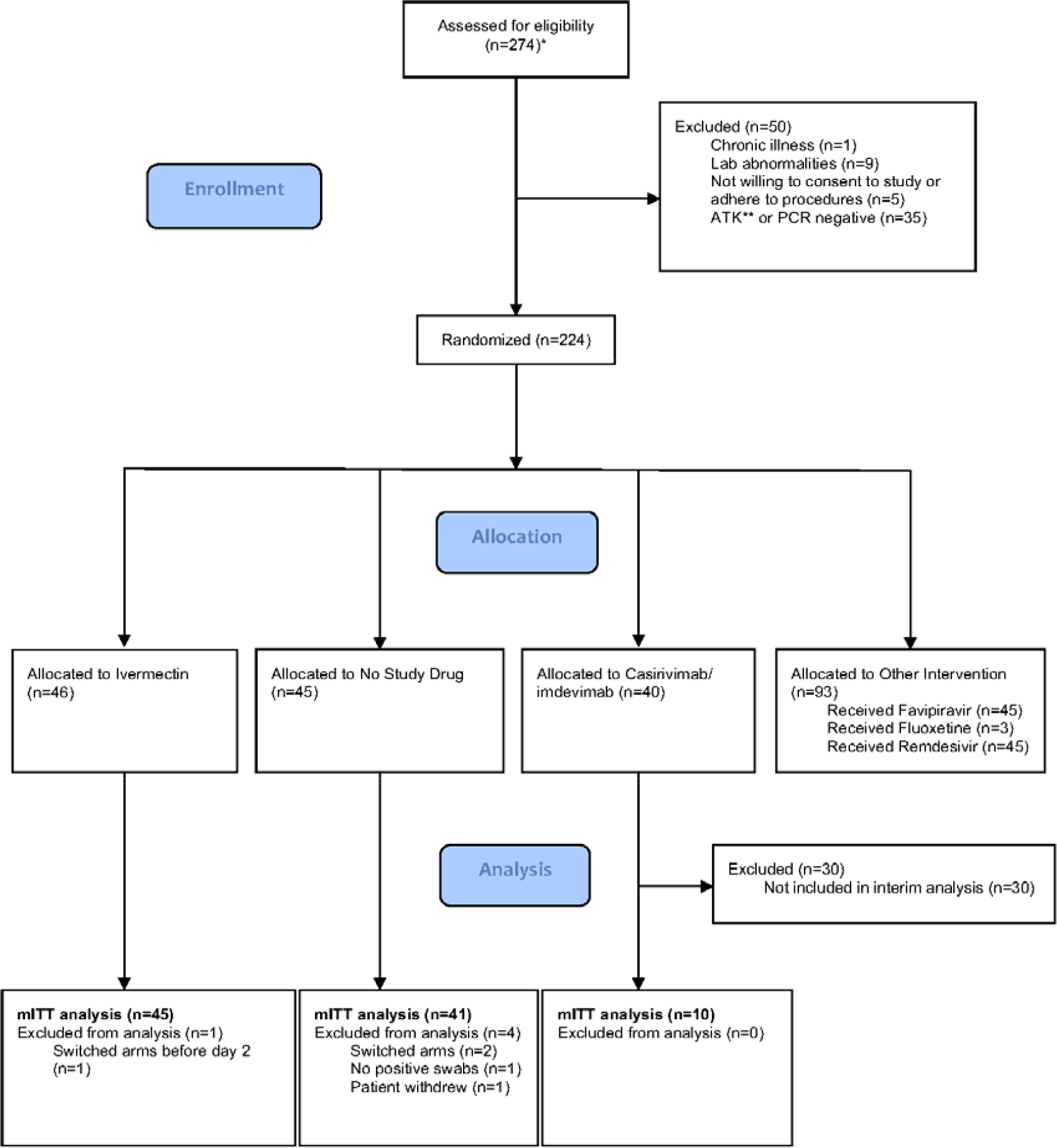
Study CONSORT diagram for the ivermectin analysis. *Pre-screening occurred in the hospitals’ Acute Respiratory Infection (ARI) units. Potentially eligible participants (based on age, duration of symptoms, reported co-morbidities and a willingness to consider study participation) were selected by the ARI Nurses to be contacted by the study team. As a result, a high proportion of those assessed for eligibility participated in the study. ** Antigen Test Kit (STANDARD™ Q COVID-19 Ag Test, SD Biosensor, Suwon-si, Republic of Korea)

### *In vivo* antiviral effects of ivermectin and casirivimab/imdevimab

The mITT population comprised 96 patients with a median of 18 viral load measurements each between days 0 and 7 (range 8-18). 7% (121/1700) of all measurements were below the lower limit of detection. The baseline geometric mean oropharyngeal viral load was 3.6 × 10^5^ RNA copies per ml (interquartile range 7.8 × 10^4^ to 2.8 × 10^6^), (Figure 2a). Viral loads declined substantially faster in casirivimab/imdevimab recipients compared to both the ivermectin and no study drug arms such that a greater than 10-fold difference in median viral loads was observed by day 3 (Figure 2b). Under a Bayesian hierarchical linear model, the population mean viral clearance half-life was estimated to be 19.2 hours (95%CI 14.8-23.9 hours) for the no study drug arm. Relative to the no study drug arm, patients randomized to ivermectin cleared oropharyngeal virus 9.1% slower (95%CI -27.2% to +11.8%), whereas patients randomized to casirivimab/imdevimab cleared oropharyngeal virus 52.3% faster (95%CI -7.0% to +115.1%), (Figure 3). This corresponded to a prolongation of virus clearance half-life of 1.9 hours (95%CI -2.1 to +6.6) for ivermectin and a shortening of 6.5 hours (95%CI -12.0 to -1.1) for casirivimab/imdevimab (Supplementary Figure 2). There was considerable inter-individual variability in viral clearance half-life-in patients in the no study drug arm with mean estimated values varying from 7 to 42 hours (Figure 4 and Supplementary Figure 2).

**Figure 2.**
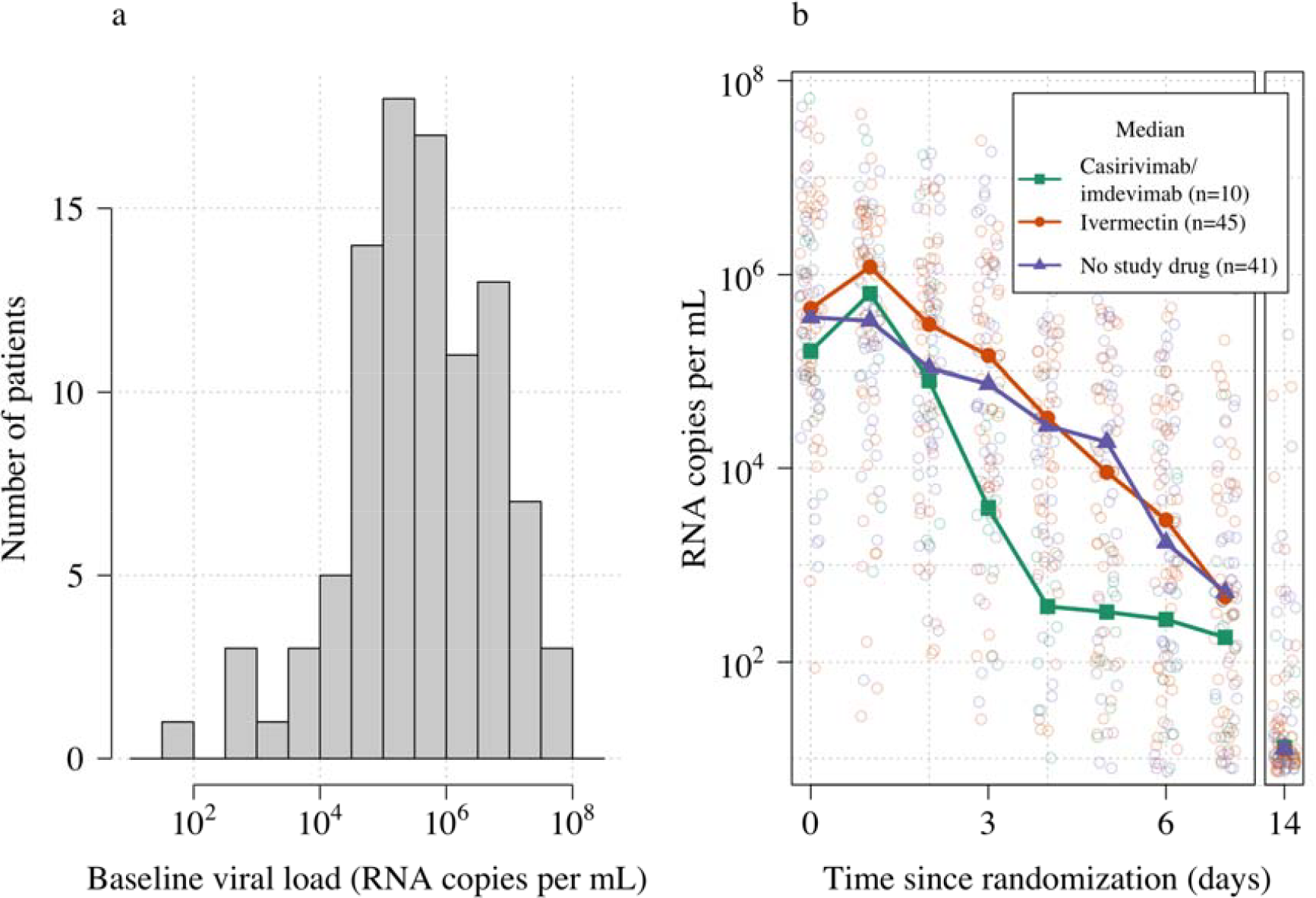
Summary of oropharyngeal viral load data in the analysis dataset (n=96). Panel a: distribution of viral loads at randomization (median of 4 swabs per patient); Panel b: individual viral load data with x-axis jitter. Median values by study arm are overlaid. The day 14 samples are not used in the primary analysis.

**Figure 3.**
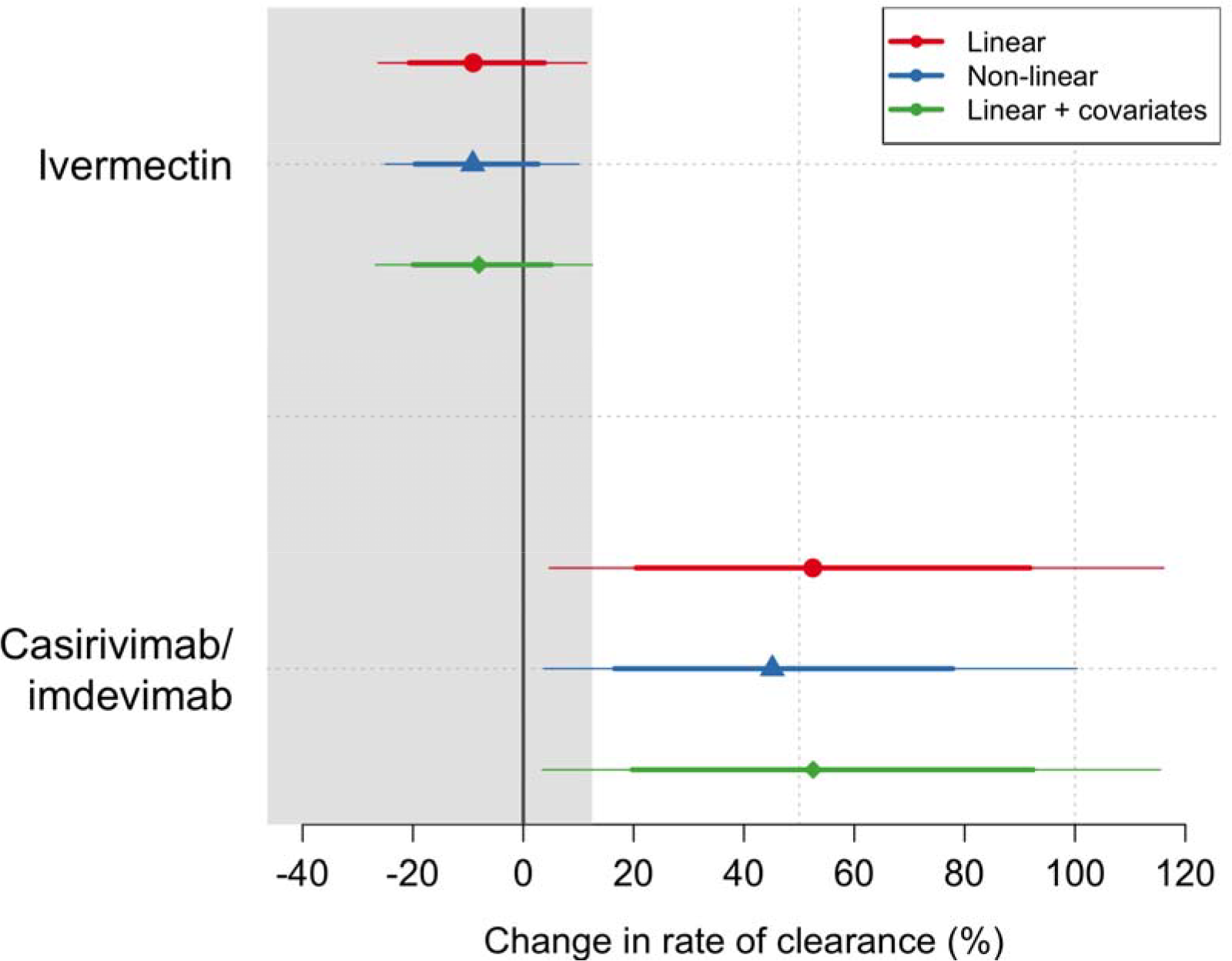
Treatment effects. Mean posterior estimates of the difference in the rate of viral clearance (thick dots) compared to the no study drug arm. 80% (thick lines) and 95% (thin lines) credible intervals under three hierarchical Bayesian models are shown. The grey area shows the futility zone (< 12.5% change in rate of viral clearance). The main model used to report effect estimates in the text is the linear model (red).

**Figure 4.**
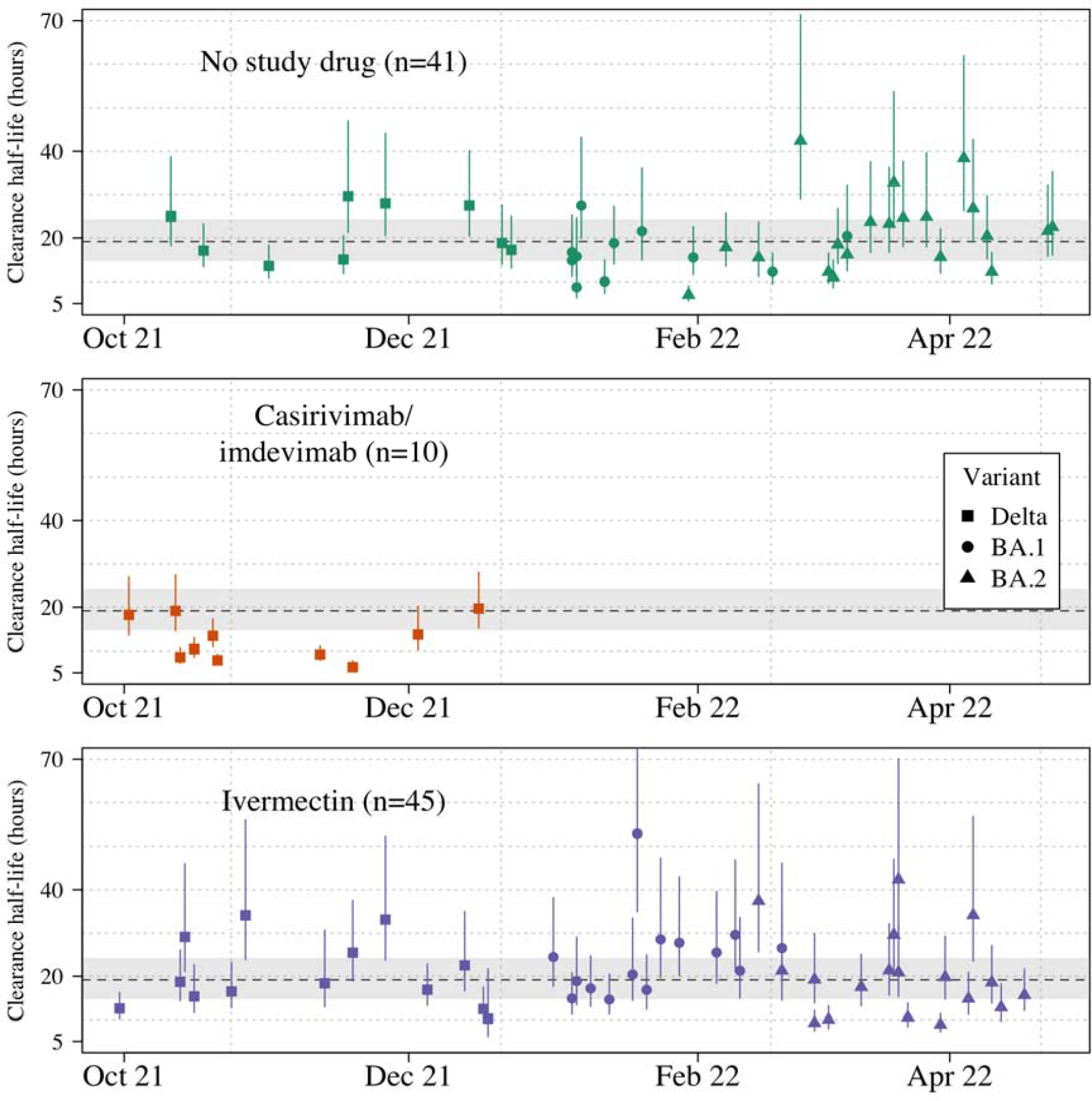
Individual patient virus clearance half-life estimates by study arm over time. The individual oropharyngeal virus clearance half-life mean posterior estimates with 95% credible intervals (lines) are shown (squares/circles/triangles corresponding to the virus genotype: temporarily Delta, Omicron BA.1, Omicron BA.2, respectively). The model estimated mean clearance half-life (95%CI) in untreated patients is shown by the grey line (dashed line-shaded area).

Targeted viral genotyping (see Supplementary materials) indicated that all 10 casirivimab/imdevimab recipients were infected with the SARS CoV-2 Delta variant (B.1.617.1) (Figure 4). The slope and intercept in all models were adjusted for site and virus variant. There was heterogeneity in the baseline viral loads and rates of clearance across recruitment sites: the two field hospitals in Thailand (Bangplee and Vajira) recruited patients with lower viral loads on average and patients in one also had faster viral clearance. However, both these sites contributed few patients (n=5 & 4, respectively). There were no apparent differences in viral clearance rates across the different virus variants/ subvariants; however, relative to the Delta variant, patients with the Omicron BA.2 subvariant had higher baseline viral loads (3.6-fold higher, 95%CI 1.4 to 9.4), and patients with the Omicron BA.1 subvariant had lower baseline viral loads (0.4-fold lower, 95%CI 0.1 to 1.1) (Supplementary Figures 3-4).

All analytical models of oropharyngeal virus clearance were in excellent agreement, giving near identical point estimates and credible intervals (Supplementary Figure 5). The best fitting model was the non-linear model which allows for some patients to have their viral load increase after randomization (i.e. patients enrolled before reaching peak viral load), followed by a log-linear decrease.

In the ivermectin recipients, there was no relationship between the viral clearance rate and the plasma AUC_0-72_ (p=0.8) or the plasma C_max_ (p=0.9). Drug exposures were high: all patients had significantly higher plasma concentrations than predicted under the pharmacokinetic model fitted to healthy volunteer data (14) (relative bioavailability of 2.6, see Supplementary Figure 6).

### Safety and Tolerability

The oropharyngeal swabbing and all treatments were well tolerated. There were three serious adverse events and these were all in the no study drug arm. Two patients had raised creatinine phosphokinase (CPK) levels greater than 10 times the upper limit of normal which were attributed to COVID-19-related rhabdomyolysis. Both patients’ laboratory tests improved with fluids and supportive management. The third patient was readmitted the day after discharge for investigation of chest pain and lethargy. Investigations were all normal and the patient was discharged the following day. Six patients reported transient visual disturbance after taking ivermectin (although not classified as grade 3 or above), and three of these withdrew from the treatment as a result (see Supplementary materials). All visual symptoms resolved quickly after the drug was stopped. All patients were referred for ophthalmology review which confirmed that no visual abnormality remained.

## Discussion

These first data from the PLATCOV adaptive platform study in early symptomatic COVID-19 show that ivermectin does not have a clinically significant antiviral effect. In contrast, the preliminary results from the casirivimab/imdevimab monoclonal antibody cocktail arm in patients infected with the SARS-CoV-2 Delta variant showed an approximate 50% increase in the rate of in viral clearance. This confirms that the study methodology can identify efficiently treatments which have clinically relevant antiviral effects *in vivo*.

It remains uncertain whether any of the proposed, and often recommended, repurposed potential antiviral treatments have significant *in vivo* antiviral activity in COVID-19. This continued uncertainty, well into the third year of the pandemic, highlights the limitations of the tools that have been used to assess antiviral activity in vivo. It is increasingly accepted that monoclonal antibodies and the newly developed specific antiviral drugs are most effective early in the course of COVID-19 infection (2, 4, 5, 15). Unfortunately, these efficacious medicines are not generally available outside high-income settings. Meanwhile repurposed therapeutics, which offered the prospect of affordable and generally available medicines, have been selected for clinical use based on their in vitro activity in cell cultures, sometimes animal studies, or clinical trials with subjective or infrequent endpoints (e.g. hospitalization or death). The large majority of these trials have been underpowered. For example, in a review of 1,314 registered studies in COVID-19, of which 1,043 (79%) were randomized controlled trials, the median (IQR) sample size was 140 patients (70-383) (16). These uncertainties have created confusion and, in the case of ivermectin, strongly polarized views.

The method of assessing antiviral activity in early COVID-19 reported in this study has the advantage of simplicity. It also avoids many of the limitations of unvalidated *in vivo* animal models (17). This approach requires a relatively small number of patients to identify antiviral activity *in vivo* (11). In this example, with 41 controls and 45 subjects receiving ivermectin, an acceleration in viral clearance of more than 12.5% could be excluded with high certainty; and with 41 controls and only 10 subjects receiving casirivimab/imdevimab an acceleration of at least 7% could be shown. These patient numbers are an order of magnitude smaller than required when the more commonly used end point of time to viral clearance (PCR negativity) is used (11). In addition, the procedures are well tolerated: daily oropharyngeal swabbing is much more acceptable than nasopharyngeal sampling.

Using less frequent sampling in large numbers of patients, clinical trials of monoclonal antibodies, and the specific antivirals molnupiravir and ritonavir-boosted nirmatrelvir, have shown that accelerated viral clearance is associated with improved clinical outcomes (4, 5). However, the quantitative relationship between the antiviral effect and clinical response varies, as host factors and viral virulence are both important determinants of outcome. Many of the studies which demonstrated improved clinical outcomes did so in high-risk, unvaccinated and previously uninfected adult individuals, with infections caused by virus variants which are no longer prevalent, and so they are sensitive to study design and temporal and epidemiological influences (18). As a result, direct comparisons between antiviral drugs using the clinical endpoints of the published phase III studies leave much uncertainty.

This trial has several limitations. It set a futility threshold of less than 12.5% acceleration of viral clearance, as currently available specific antiviral therapies suggest probable effect sizes of a 30 to 50% acceleration (4-6). It does not, therefore, exclude a smaller antiviral effect, or a benefit from a non-antiviral effect (e.g. an immunomodulatory effect). Whether a smaller antiviral effect would warrant recommendation in treatment is debatable, but it could still be very useful in prevention, which requires less potent viral suppression for a clinical benefit. It is also uncertain whether the daily sampling schedule is the optimal balance between statistical power and trial feasibility and acceptability. This could change with continued viral evolution and increasing vaccine coverage potentially affecting viral clearance parameters. There is substantial variability in estimated serial viral loads (see individual profiles shown in Supplementary Figure 7). Whether this variability can be reduced by adjusting for extracellular fluid content or different sampling techniques is uncertain. Finally, the viral qPCR measures viral genomes and does not distinguish between live (potentially transmissible) and dead virus.

High dose ivermectin did not have measurable antiviral activity in early symptomatic COVID-19. This study provides no support for the continued use of ivermectin in COVID-19. Efficient characterization and comparison of potential antiviral therapeutics in COVID-19 will be important for policy recommendations, particularly while cost and availability limit access. Use of the rate of oropharyngeal viral clearance as a metric for antiviral efficacy has applicability beyond COVID-19 including other respiratory illnesses, notably influenza (19), novel coronaviruses and future, as yet unknown, respiratory illnesses.

## Supporting information

Master Protocol v1.1

## Data Availability

All code and data are openly accessible via the following GitHub repository: https://github.com/jwatowatson/PLATCOV-Ivermectin.

## Acknowledgements

This research was funded by the Wellcome Trust. A CC BY, or equivalent licence, is applied to any author accepted manuscript arising from this submission, in accordance with the grant’s open access conditions.

## References

1. Weinreich DM, Sivapalasingam S, Norton T, Ali S, Gao H, Bhore R, Xiao J, Hooper AT, Hamilton JD, Musser BJ, Rofail D, Hussein M, Im J, Atmodjo DY, Perry C, Pan C, Mahmood A, Hosain R, Davis JD, Turner KC, Baum A, Kyratsous CA, Kim Y, Cook A, Kampman W, Roque-Guerrero L, Acloque G, Aazami H, Cannon K, Simon-Campos JA, Bocchini JA, Kowal B, DiCioccio AT, Soo Y, Geba GP, Stahl N, Lipsich L, Braunstein N, Herman G, Yancopoulos GD, Trial I. REGEN-COV Antibody Combination and Outcomes in Outpatients with Covid-19. N Engl J Med. 2021;385(23):e81.

2. O’Brien MP, Forleo-Neto E, Musser BJ, Isa F, Chan KC, Sarkar N, Bar KJ, Barnabas RV, Barouch DH, Cohen MS, Hurt CB, Burwen DR, Marovich MA, Hou P, Heirman I, Davis JD, Turner KC, Ramesh D, Mahmood A, Hooper AT, Hamilton JD, Kim Y, Purcell LA, Baum A, Kyratsous CA, Krainson J, Perez-Perez R, Mohseni R, Kowal B, DiCioccio AT, Stahl N, Lipsich L, Braunstein N, Herman G, Yancopoulos GD, Weinreich DM, Covid-19 Phase 3 Prevention Trial T. Subcutaneous REGEN-COV Antibody Combination to Prevent Covid-19. N Engl J Med. 2021;385(13):1184–95.

3. Bruel T, Hadjadj J, Maes P, Planas D, Seve A, Staropoli I, Guivel-Benhassine F, Porrot F, Bolland WH, Nguyen Y, Casadevall M, Charre C, Pere H, Veyer D, Prot M, Baidaliuk A, Cuypers L, Planchais C, Mouquet H, Baele G, Mouthon L, Hocqueloux L, Simon-Loriere E, Andre E, Terrier B, Prazuck T, Schwartz O. Serum neutralization of SARS-CoV-2 Omicron sublineages BA.1 and BA.2 in patients receiving monoclonal antibodies. Nat Med 28, 1297–1302 (2022).

4. Jayk Bernal A, Gomes da Silva MM, Musungaie DB, Kovalchuk E, Gonzalez A, Delos Reyes V, Martin-Quiros A, Caraco Y, Williams-Diaz A, Brown ML, D. J, Pedley A, Assaid C, Strizki J, Grobler JA, Shamsuddin HH, Tipping R, Wan H, Paschke A, Butterton JR, Johnson MG, De Anda C, Group MO-OS. Molnupiravir for Oral Treatment of Covid-19 in Nonhospitalized Patients. N Engl J Med. 2022;386(6):509–20.

5. Hammond J, Leister-Tebbe H, Gardner A, Abreu P, Bao W, Wisemandle W, Baniecki M, Hendrick VM, Damle B, Simon-Campos A, Pypstra R, Rusnak JM, Investigators E-H. Oral Nirmatrelvir for High-Risk, Nonhospitalized Adults with Covid-19. N Engl J Med. 2022;386(15):1397–408.

6. Robinson PC, Liew DFL, Tanner HL, Grainger JR, Dwek RA, Reisler RB, Steinman L, Feldmann M, Ho LP, Hussell T, Moss P, Richards D, Zitzmann N. COVID-19 therapeutics: Challenges and directions for the future. Proc Natl Acad Sci U S A. 2022;119(15):e2119893119.

7. Caly L, Druce JD, Catton MG, Jans DA, Wagstaff KM. The FDA-approved drug ivermectin inhibits the replication of SARS-CoV-2 in vitro. Antiviral Res. 2020;178:104787.

8. Mega ER. Latin America’s embrace of an unproven COVID treatment is hindering drug trials. Nature 2020. https://www.nature.com/articles/d41586-020-02958-2

9. Lawrence JM, Meyerowitz-Katz G, Heathers JAJ, Brown NJL, Sheldrick KA. The lesson of ivermectin: meta-analyses based on summary data alone are inherently unreliable. Nat Med. 2021;27(11):1853–4.

10. Reis G, Silva E, Silva DCM, Thabane L, Milagres AC, Ferreira TS, Dos Santos CVQ, Campos VHS, Nogueira AMR, de Almeida A, Callegari ED, Neto ADF, Savassi LCM, Simplicio MIC, Ribeiro LB, Oliveira R, Harari O, Forrest JI, Ruton H, Sprague S, McKay P, Guo CM, Rowland-Yeo K, Guyatt GH, Boulware DR, Rayner CR, Mills EJ, Investigators T. Effect of Early Treatment with Ivermectin among Patients with Covid-19. N Engl J Med. 2022;386(18):1721–31.

11. Watson JA, Kissler SM, Day NPJ, Grad YH, White NJ. Characterizing SARS-CoV-2 Viral Clearance Kinetics to Improve the Design of Antiviral Pharmacometric Studies. Antimicrob Agents Chemother. 2022 Jun 23:e0019222. doi: 10.1128/aac.00192-22. Epub ahead of print. PMID: 35736134.

12. Hanpithakpong W, Kamanikom B, Dondorp AM, Singhasivanon P, White NJ, Day NP, Lindegardh N. A liquid chromatographic-tandem mass spectrometric method for determination of artesunate and its metabolite dihydroartemisinin in human plasma. J Chromatogr B Analyt Technol Biomed Life Sci. 2008;876(1):61–8.

13. Lindegardh N, Annerberg A, White NJ, Day NP. Development and validation of a liquid chromatographic-tandem mass spectrometric method for determination of piperaquine in plasma stable isotope labeled internal standard does not always compensate for matrix effects. J Chromatogr B Analyt Technol Biomed Life Sci. 2008;862(1-2):227–36.

14. Kobylinski KC, Jittamala P, Hanboonkunupakarn B, Pukrittayakamee S, Pantuwatana K, Phasomkusolsil S, Davidson SA, Winterberg M, Hoglund RM, Mukaka M, van der Pluijm RW, Dondorp A, Day NPJ, White NJ, Tarning J. Safety, Pharmacokinetics, and Mosquito-Lethal Effects of Ivermectin in Combination With Dihydroartemisinin-Piperaquine and Primaquine in Healthy Adult Thai Subjects. Clin Pharmacol Ther. 2020;107(5):1221–30.

15. Gottlieb RL, Vaca CE, Paredes R, Mera J, Webb BJ, Perez G, Oguchi G, Ryan P, Nielsen BU, Brown M, Hidalgo A, Sachdeva Y, Mittal S, Osiyemi O, Skarbinski J, Juneja K, Hyland RH, Osinusi A, Chen S, Camus G, Abdelghany M, Davies S, Behenna-Renton N, Duff F, Marty FM, Katz MJ, Ginde AA, Brown SM, Schiffer JT, Hill JA, Investigators G-U-. Early Remdesivir to Prevent Progression to Severe Covid-19 in Outpatients. N Engl J Med. 2022;386(4):305–15.

16. McLean ARD, Rashan S, Tran L et al. The fragmented COVID-19 therapeutics research landscape: a living systematic review of clinical trial registrations evaluating priority pharmacological interventions. [version 1; peer review: 1 approved]. Wellcome Open Res 2022, 7:24

17. Munoz-Fontela C, Widerspick L, Albrecht RA, Beer M, Carroll MW, de Wit E, Diamond MS, Dowling WE, Funnell SGP, Garcia-Sastre A, Gerhards NM, de Jong R, Munster VJ, Neyts J, Perlman S, Reed DS, Richt JA, Riveros-Balta X, Roy CJ, Salguero FJ, Schotsaert M, Schwartz LM, Seder RA, Segales J, Vasan SS, Henao-Restrepo AM, Barouch DH. Advances and gaps in SARS-CoV-2 infection models. PLoS Pathog. 2022;18(1):e1010161.

18. Sigal A, Milo R, Jassat W. Estimating disease severity of Omicron and Delta SARS-CoV-2 infections. Nat Rev Immunol. 2022;22(5):267–9.

19. Yu H, Liao Q, Yuan Y, Zhou L, Xiang N, Huai Y, Guo X, Zheng Y, van Doorn HR, Farrar J, Gao Z, Feng Z, Wang Y, Yang W. Effectiveness of oseltamivir on disease progression and viral RNA shedding in patients with mild pandemic 2009 influenza A H1N1: opportunistic retrospective study of medical charts in China. BMJ. 2010;341:c4779.

