## Supplementary material for "Pharmacometric assessment of the *in vivo* antiviral activity of ivermectin in early symptomatic COVID-19": Master Protocol v1.1

**Finding treatments for COVID-19: A phase 2 multi-centre adaptive platform trial to assess antiviral pharmacodynamics in early symptomatic COVID-19 (PLATCOV)**

**Short title:** Finding treatments for COVID-19: A phase 2 platform trial of antiviral pharmacodynamics in early symptomatic COVID-19 (PLATCOV)

**Acronym:** PLATCOV

**OxTREC Ref:** 24-21

**Trial Registration Number**: NCT05041907

**Protocol number:** VIR21001

**Date and Version No:** Version MASTER 1.1/ 01 June 2022

| **Co-Principal Investigators:** | **Professor Sir Nicholas J White**  Mahidol Oxford Tropical Medicine Research Unit, Faculty of Tropical Medicine, Mahidol University  420/6 Ratchawithi Road,Ratchathewi Bangkok 10400 Thailand  **Dr William Schilling**  Mahidol Oxford Tropical Medicine Research Unit, Faculty of Tropical Medicine, Mahidol University  420/6 Ratchawithi Road,Ratchathewi Bangkok 10400 Thailand |
| --- | --- |
| **Co-Investigators:** | **Dr. Simon Boyd**  Mahidol Oxford Tropical Medicine Research Unit, Faculty of Tropical Medicine, Mahidol University  420/6 Ratchawithi Road,Ratchathewi Bangkok 10400 Thailand  **Dr James Callery**  Research Physician  Mahidol Oxford Tropical Medicine Research Unit, Faculty of Tropical Medicine, Mahidol University, Bangkok,Thailand  **Dr Cintia Cruz**  Paediatrician and Clinical Pharmacologist  Mahidol Oxford Tropical Medicine Research Unit, Faculty of Tropical Medicine, Mahidol University, Bangkok, Thailand  **Professor Nicholas PJ Day**  Director & Consultant Physician  Mahidol Oxford Tropical Medicine Research Unit, Faculty of Tropical Medicine, Mahidol University, Bangkok,Thailand  **Professor Arjen M Dondorp**  Deputy Director and Head of Malaria & Critical Illness  Mahidol Oxford Tropical Medicine Research Unit, Faculty of Tropical Medicine, Mahidol University, Bangkok,Thailand  **Assoc. Prof. Weerapong Phumratanaprapin**  Department of Clinical Tropical Medicine  Faculty of Tropical Medicine, Mahidol University  420/6 Ratchawithi Road,Ratchathewi Bangkok 10400 Thailand  **Dr Walter RJ Taylor**  Senior Clinical Researcher and Infectious Diseases Physician  Mahidol Oxford Tropical Medicine Research Unit, Faculty of Tropical Medicine, Mahidol University, Bangkok,Thailand  **Professor Mauro Teixeira**  Unidade de Pesquisa Clínica,  Centro de Terapias Avançadas e Inovadoras  Universidade Federal de Minas Gerais,  Belo Horizonte, MG, Brasil  **Dr James A Watson**  Statistician  Mahidol Oxford Tropical Medicine Research Unit, Faculty of Tropical Medicine, Mahidol University, Bangkok,Thailand |

**Investigators’ contributions:**

*NJW* conceived of the study and initiated the study design with other investigators.

**Confidentiality Statement**

This document contains confidential information that must not be disclosed to anyone other than the Sponsor, the Investigator Team, host organisation, and members of the Research Ethics Committee and Regulatory Authorities unless authorised to do so.

**Investigator Agreement and Declaration of Interests**

“The undersigned has read and understood the trial protocol detailed above and:

- agrees to conduct the trial in compliance with the protocol.
- agrees to comply with the principles of the International Conference on Harmonisation Tripartite Guideline on Good Clinical Practice.

and declare no conflict of interest, according to the current version of the Declaration of Helsinki”

| Dr William Schilling | 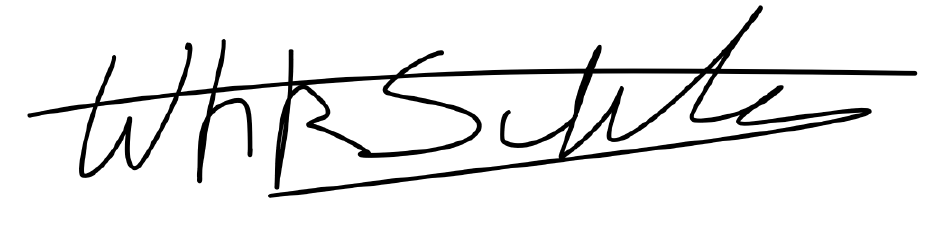 | …01/06/2022……… |
| --- | --- | --- |
| Co-Principal Investigator | Signature | Date |

| Professor Sir Nicholas J White | 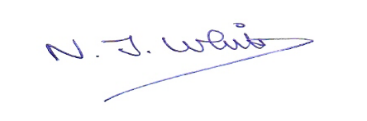  ………………………………..…… | …01/06/2022……… |
| --- | --- | --- |
| Co-Principal Investigator | Signature | Date |

### 1. **SYNOPSIS**

| **Study Title** | Finding treatments for COVID-19: A phase 2 multi-centre adaptive platform trial to assess antiviral pharmacodynamics in early symptomatic COVID-19 (PLATCOV) | |
| --- | --- | --- |
| **Protocol no.** | VIR21001 | |
| **Rationale** | Quantitative evidence of antiviral activity in patients with COVID-19 is required to justify phase 3 clinical trials of putative antivirals | |
| **Study Design** | Randomised, open label, group sequential adaptive platform trial | |
| **Inclusion and Exclusion Criteria** | **Inclusion criteria:**   - Patient understands the procedures and requirements and is willing and able to give informed consent for full participation in the study - Previously healthy adults, male or female, aged 18 to 50 years at time of consent with early symptomatic COVID-19 - SARS-CoV-2 positive by lateral flow antigen test OR a positive PCR test for SARS-CoV-2 within the last 24hrs with a Ct value of less than 25 (all viral targets) - Reported symptoms of COVID-19 (including fever, or history of fever) for less than 4 days (96 hours) - Oxygen saturation ≥96% measured by pulse oximetry at time of screening - Able to walk unaided and unimpeded in ADLs - Agrees and is able to adhere to all study procedures, including availability and contact information for follow-up visits     **Exclusion criteria:**   - Taking any concomitant medications or drugs (see appendix 4)† - Presence of any chronic illness/ condition requiring long term treatment, or other significant comorbidity (see appendix 4 for full list) - Laboratory abnormalities discovered at screening (see appendix 4) - For females: pregnancy, actively trying to become pregnant, or lactation - Contraindication to taking, or known hypersensitivity reaction to any of the proposed therapeutics (see appendix 4) - Currently participating in another COVID-19 therapeutic or vaccine trial - Evidence of pneumonia (although imaging is NOT required)   † healthy women on the oral contraceptive pill are eligible to join the study | |
| **Planned Sample Size** | Continuously running group sequential adaptive platform trial. There is no fixed sample size (see section 10.1 for further discussion) | |
| **Planned Study Period** | 2 years for total duration of the trial and 28 days for individual patients’ involvement | |
| **Interventions** | The platform trial will assess drugs with potential SARS-CoV-2 antiviral activity of three general types:   A. Newly available and repurposed potential antiviral drugs (see appendix 2) B. Positive control: monoclonal antibodies initially but subsequently any therapeutic that is shown to accelerate the rate of viral clearance C. Novel small molecule drugs that have gone through phase 1 testing | |
| **Control** | No antiviral treatment (although local hospital supportive treatment will remain the same e.g. antipyretics, anti-tussives, antihistamines, vitamins etc as per the treating Physician’s judgement) | |
| **Rationale** | Each intervention type has a different objective:   A. Newly available and repurposed drugs are already used and recommended in some countries. Showing that they do not have significant antiviral activity is as important as showing that they do. For the newly approved antivirals, comparing antiviral activities in-vivo will inform health authorities’ recommendations. B. Positive control: There is good evidence from phase 2 studies that monoclonal antibodies reduce viral load in COVID-19 with some evidence that they reduce hospitalisation in high-risk individuals. However, monoclonal antibodies are vulnerable to viral escape mutations. Tracking their performance over time is important to characterise the impact and inform the therapeutics of mutant SARS-CoV-2 strains. This will also be important for other antivirals. Monoclonal antibodies are expensive and cannot be produced at large scale currently, but this may change in the near future. These drugs may not be available early in the study, and will be included if there is local availability and regulatory approval. C. New small molecule drugs in development that pass phase 1 testing will have a stronger rationale (i.e. *a priori* are more likely to be effective) compared to repurposed drugs. Rapidly identifying the most effective candidate and the optimal dosing for phase 3 trials is the ultimate goal. | |
|  | **Objectives** | **Endpoint** |
| **Primary** | A. For newly available and repurposed drugs   To evaluate COVID-19 antiviral efficacy in-vivo   B. For positive controls (e.g. monoclonal antibodies)   To characterise in-vivo performance over time relative to the no antiviral treatment control   C. For novel small molecule drugs   To evaluate COVID-19 antiviral efficacy in-vivo (accelerated viral clearance) | Rate of viral clearance- estimated from the log_10_ viral density derived from qPCR of standardised duplicate oropharyngeal swabs/ saliva taken daily from baseline (day 0) to day 7 for each therapeutic arm compared with the no antiviral treatment control i.e. those not receiving study drug |
| **Secondary** | To characterise the determinants of viral kinetics in early COVID-19 disease | Rate of viral clearance (as for primary endpoint) |
|  | To determine optimal dosing regimens through pharmacometric assessment in antiviral drugs that are shown to be effective i.e. a positive signal (>90% probability of >12.5% acceleration in viral clearance) or evidence of antiviral effects from other studies (e.g. Molnupiravir, Nirmatrelvir/ritonavir) | Rate of viral clearance (as for primary endpoint) |
|  | To compare viral clearance of any discovered active antiviral drugs against the positive control (e.g. REGN-COV-2 a monoclonal antibody cocktail) or other licensed and available therapeutics with evidence of accelerated viral clearance | Rate of viral clearance (as for primary endpoint) |
| **Tertiary** | Characterise the relationship between viral clearance and hospitalisation (hospitalisation for clinical reasons) | Hospitalisation for clinical reasons up to day 28 |

### 2. **ABBREVIATION**

| ACT | Access to COVID-19 Tools |
| --- | --- |
| ADLs | Activities of daily living |
| AE | Adverse event |
| AR | Adverse reaction |
| ARI | Acute Respiratory Infection |
| COVAX | **COVID-19 Vaccines Global Access** |
| COVID-19 | Coronavirus disease of 2019 |
| CRF | Case Report Form |
| CTCAE | Common Terminology Criteria for Adverse Events |
| CTSG | Clinical Trials Support Group |
| DSMB | Data Safety Monitoring Board |
| EC | Ethic Committee |
| EDTA | Ethylenediaminetetraacetic acid |
| FBC | Full blood count |
| FDA | Food and Drug Administration |
| GCP | Good Clinical Practice |
| ICF | Informed Consent Form |
| ICH | International Conference on Harmonisation |
| IL | **Interleukin** |
| IM | Intramuscular |
| IMP | Investigational Medicinal Product |
| IV | Intravenous |
| LDH | Lactate dehydrogenase |
| LFT | Liver function test |
| MORU | Mahidol Oxford Tropical Medicine Research Unit |
| NGS | Next Generation Sequencing |
| OxTREC | Oxford Tropical Research Ethics Committee |
| PD | Pharmacodynamic |
| PI | Prinicipal investigator |
| PK | Pharmacokinetics |
| POC | Point-of-care |
| PPE | Personal protective equipment |
| RDT | Rapid diagnostic test |
| REGN | Regeneron |
| RT-qPCR | Real-time quantitative polymerase chain reaction |
| SAE | Serious Adverse Event |
| SAP | Statistical Analysis Plan |
| SAR | Serious Adverse Reaction |
| SARS-CoV2 | Severe acute respiratory syndrome coronavirus 2 |
| SC | Subcutaneous |
| SOP | Standard Operating Procedure |
| SUSAR | Suspected Unexpected Serious Adverse Reactions |
| TNF | Tumor necrosis factor |
| TMG | Trial Management Group |
| U&E | Urea and Electrolytes |
| VTM | Viral transport medium |
| WHO | World Health Organization |

### 3. BACKGROUND AND RATIONALE

There are many potential therapeutics for COVID-19 and a much larger number of vaccines are in development. Vaccines are being given to individuals with the primary aim of reducing morbidity and mortality and the secondary objective of generating herd immunity in the hope of bringing the pandemic to an end. However, there is widespread vaccine inequality. As of 13 January 2022, of the 194 Member States of the WHO, 36 have vaccinated less than 10% of their population, and 88 less than 40% (1) This demonstrates how many will remain vulnerable to infection for the foreseeable future. Vaccines are the solution but there are also concerns over incomplete protection, vaccine hesitancy, waning protective effects over time, and the rapid evolution of more transmissible and potentially vaccine resistant mutant strains (highlighted by the Omicron variant (B.1.1.529), and future variants (which may not be as comparatively mild). We know that vaccine and disease-induced immunity is imperfect and short-lived so identifying active antiviral drugs is extremely important. Many people over the next 2-3 years will get COVID-19 with substantial morbidity and hundreds of thousands of deaths. There will always be people, due to their age, comorbidities or being immunocompromised that will need early antiviral therapy. For all these reasons effective therapeutics are needed urgently.

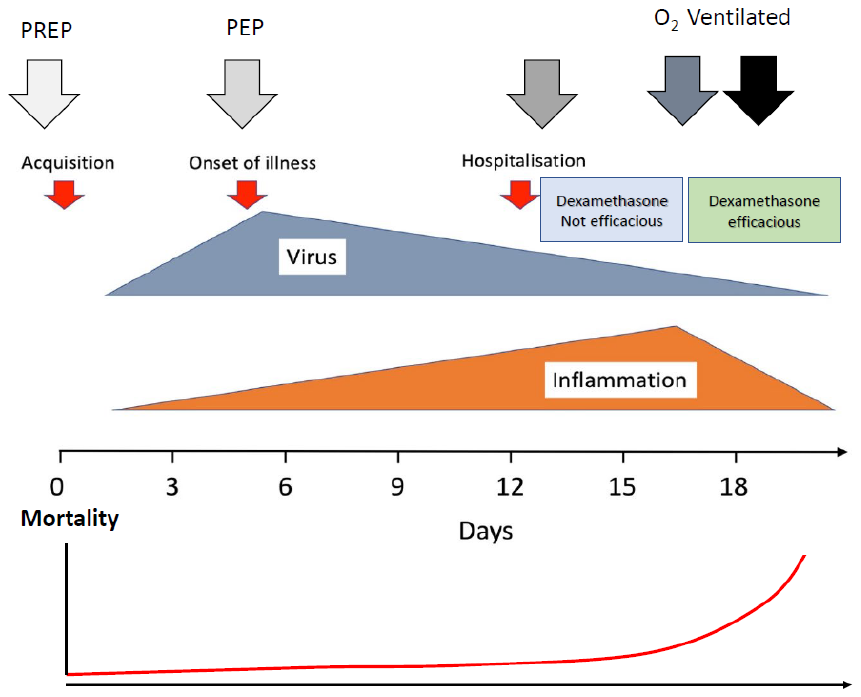

Figure 1 Simplified paradigm of COVID-19 infection and disease. There are two main stages of disease and antiviral therapies should be most effective early on.

Symptomatic COVID-19 infection has two overlapping phases (Figure 1). In the first phase, there is rapid viral replication with peak viral loads in the pharynx occurring approximately at the time of symptom onset. Thereafter viral burdens decline. In this second phase, the viral load decreases (the decline is first order, and the rate of decline is slower in sicker patients) (2-6). During the second phase, a small minority of individuals (particularly the elderly and those with co-morbidities) progress to severe illness (severe pneumonia) and some die. Hospital admission is usually about one week after the onset of illness (7). In those patients requiring respiratory support, low dose dexamethasone reduces mortality substantially (8). At this late stage of the illness inflammatory processes predominate, and antivirals are less likely to be of benefit (9). Antiviral treatments are most likely to be of benefit when given early in the illness. Accelerating viral clearance should reduce the subsequent inflammatory pathology, a hypothesis which is supported by studies of monoclonal antibody therapies and data from hospitalised patients (10, 11). At the time of writing the early protocol, there were promising monoclonal antibody therapies with evidence of viral clearance acceleration. Hospitalisation rates in RCTs were lower in the patients receiving these antibodies. However, monoclonal antibodies are currently expensive and difficult to deploy at scale (although this could change rapidly in the near future) and the continued efficacy of monoclonal antibodies is threatened by the evolution and spread of spike protein mutants (12, 13). This has been seen recently with the spread of the Omicron variant which has led, on the basis of in-vitro studies demonstrating decreased neutralization, to curtailment of their use. Two small molecule antivirals (molnupiravir, nirmatrelvir boosted by ritonavir) have completed phase 3 testing, and shown in-vivo benefits in COVID-19. They are already licensed for clinical use in some countries. However, further studies are needed, particularly comparative data on viral clearance between these two drugs. More antivirals will likely follow, and comparisons of their antiviral effects will help healthcare systems decide on their purchase and use.

There is no optimised or validated approach to assess rapidly potential antiviral therapeutics in COVID-19. Drugs are currently being selected for clinical study on the basis of activity in cell culture systems (in-vitro) and animal models in-vivo. Unfortunately the animal models are not sufficiently good to be included in the drug development critical pathway. Some manufacturers (e.g. Pfizer) have decided not to try and assess antiviral activity of candidate drugs in these animal models. Thus drugs currently under consideration are justified only on the basis of antiviral activity in cell cultures. In order to identify effective antivirals and optimise their dosing, and therefore ensure that phase 3 studies are designed appropriately, and progress is as rapid as possible, in-vivo antiviral effects must be characterized adequately. This can be achieved in natural COVID-19 infections at an early stage of the disease using the following design. The proposed trial will develop and validate a platform for quantitative assessment of antiviral effects (5) in low-risk patients with high viral burdens and uncomplicated COVID-19.

#### 3.1. Proposal

In this randomised open label, controlled, group sequential adaptive platform trial, we will assess the performance of three distinct types of intervention relative to control (no treatment):

A: Newly available and repurposed antiviral drugs; and if available:

B: "Positive control” (e.g. monoclonal antibodies); and later:

C: Small molecule drugs that pass phase 1 testing.

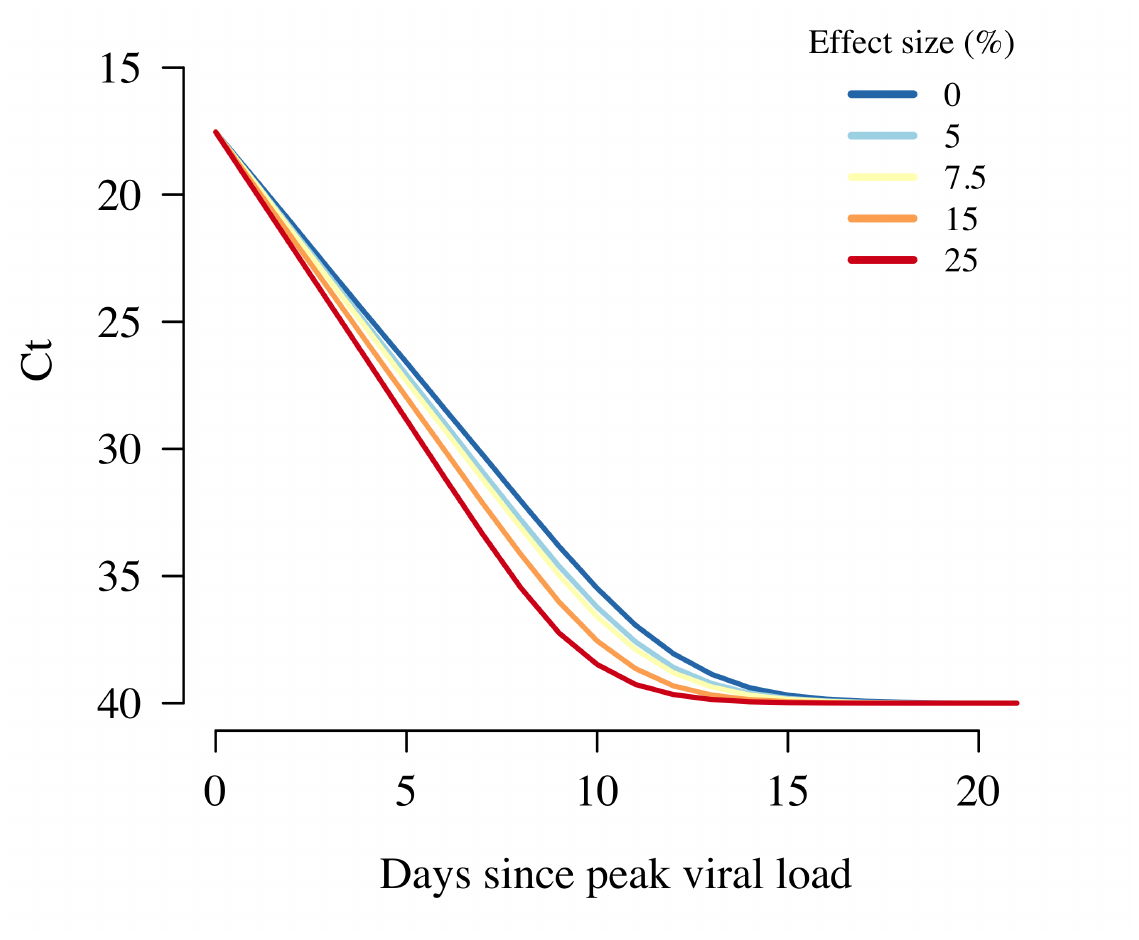
The trial initially used repurposed drugs as they were available at the time, and some are used widely without good evidence of benefit. Subsequently newly developed small molecule antiviral drugs have become available (e.g. Molnupiravir and Nirmatrelvir/ritonavir). For these newly available and repurposed drugs under consideration (to be defined as type A interventions), the platform trial is designed based on the premise that showing clear evidence of an antiviral effect is as important for medicine and public health as proving there is no significant effect (i.e. Type A adaptive design best suits the trial aims here). Antivirals shown to be beneficial in the study may subsequently be chosen to be Type B interventions (e.g. “Positive controls”). Proving that a drug does not accelerate viral clearance in-vivo should provide evidence to stop clinical recommendations based on claims of putative antiviral activity in ex-vivo cell based assays. Monoclonal antibodies (to be defined as type B interventions) were initially included as a “positive control”. This is beneficial to demonstrate that the method does identify active compounds. It will be important to calibrate and compare the effects of these validated effective therapies relative to the newly identified small molecule antiviral drugs. In addition, for monoclonal antibodies it is also important to characterise waning efficacy over time resulting from novel spike protein mutants (12,13)

Figure 2. Acceleration of viral clearance rates by different amounts

We aim to identify the best candidates rapidly of the novel small molecule drugs. The distinct objectives for each of these intervention types requires different stopping rules and design features. The rapid turn-round in quantitative PCR measurements is key to the success of this project. Analysis of these results will allow for frequent interim analyses with the aim of halting study arms early for futility, or triggering an intensive nested PK-PD study if there is evidence of clinically significant antiviral activity, although early interim analyses are not likely to yield efficacy assessments, and are aimed primarily at validation of the methodology. This will speed up the identification of antivirals to be assessed in phase III studies. The key metric is the rate of viral clearance (5) (Figure 2).

#### 3.2. Pharmacodynamic assessment

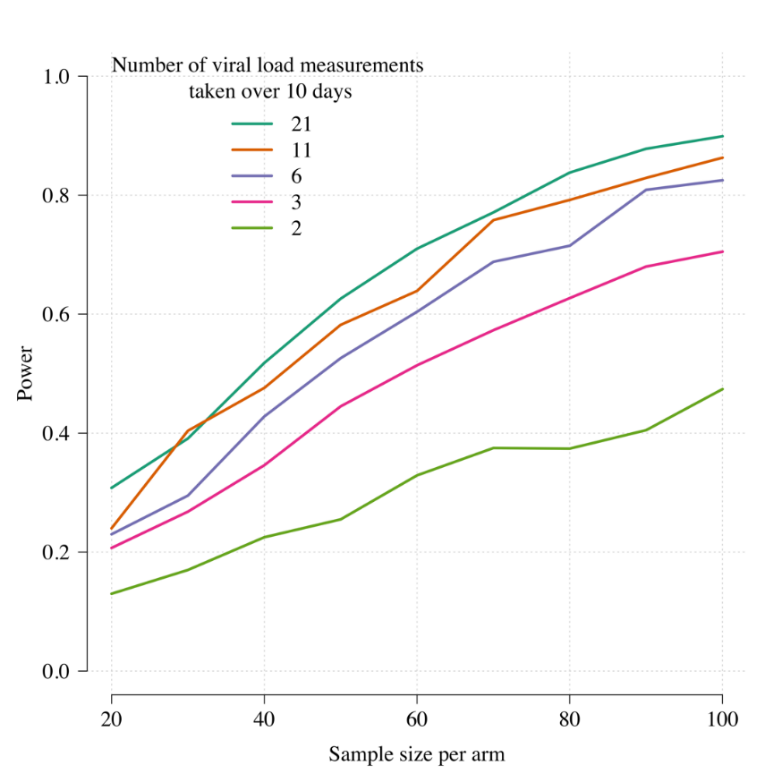
We have used prospectively collected serial viral load data from 46 individuals infected with SARS-CoV-2 and an accompanying pharmacodynamic model to simulate different trial designs for putative antiviral drugs using either time-to-clearance or rate-of-clearance as the primary trial endpoint (15). This model was then tested on data from a small pilot study of 24 patients with early symptomatic COVID-19 who were randomised to either ivermectin or placebo (16). **Rate-of-clearance was shown to be a uniformly better endpoint in terms of type 2 error (i.e. results in increased power) than the widely used and reported time-to-clearance (4).** Rate of decline in oropharyngeal qPCR estimates of viral density measured daily also allows shorter and more efficient evaluation of viral clearance (Figure 3). Our simulations (Appendix 3) show that the proposed trial design can identify an effective therapy rapidly in the setting of a platform trial and quickly discard ineffective therapies (both of which are the primary objectives of the trial).

Figure 3. The relationship between the number of qPCR viral load measurements taken over 10 days and the power (1-type 2 error) to detect an effect size of 10% increase in viral clearance rate relative to control. This assumes that patients are recruited at peak viral load (4).

### 4. OBJECTIVES AND ENDPOINTS

|  | **Objectives** | **Endpoint** | **Timepoint(s) of evaluation of this endpoint (if applicable)** |
| --- | --- | --- | --- |
| **Primary** | A. **For newly available and repurposed drugs**   **To evaluate COVID-19 antiviral efficacy**   B. For positive controls (e.g. monoclonal antibodies)   To characterise performance (over time) relative to the no antiviral treatment control   C. For novel small molecule drugs   To evaluate COVID-19 antiviral efficacy (accelerated viral clearance) | Rate of viral clearance (estimated from the log_10_ viral density derived from qPCR of standardised oropharyngeal swabs/ saliva taken daily from baseline (day 0) to day 7 for each therapeutic arm compared with the no antiviral treatment control i.e. patients not receiving study drug) | Days 0-7 |
| **Secondary** | To characterise the determinants of viral kinetics in early COVID-19 disease. | Rate of viral clearance | Days 0-7 |
|  | To determine optimal dosing regimens through pharmacometric assessment in antiviral drugs that are shown to be effective i.e. show a positive signal (>90% probability of >12.5% acceleration in viral clearance) or evidence of antiviral effects from other studies (e.g. Molnupiravir, Nirmatrelvir/ritonavir) | Rate of viral clearance | Days 0-7 |
|  | To compare viral clearance of any discovered active antiviral drugs against the positive control (e.g. REGN-COV2 (monoclonal antibody cocktail) or other licensed and available therapeutics with evidence of accelerated viral clearance | Rate of viral clearance | Days 0-7 |
| **Tertiary** | Characterise the relationship between viral clearance and hospitalisation (hospitalisation for clinical reasons) | Hospitalisation for clinical reasons up to day 28 | Days 0-28 |

### 5. STUDY DESIGN

The study is a randomised, open label, controlled adaptive platform trial that will be conducted in low-risk patients with COVID-19, recruited from outpatient COVID-19/ Acute Respiratory Infection (ARI) clinics or through other approved facilities, or from inpatient isolation facilities, or by patient self-referral to the study site, that is previously healthy patients 18 to 50 years old with early symptomatic COVID-19 and without co-morbidities (see appendix 4). The study has started in Thailand and Brazil.

After obtaining fully informed consent, we will recruit adult patients with early symptomatic COVID-19 (less than 4 days (96 hours) since the reported onset of symptoms), who are positive by a SARS-CoV-2 lateral flow antigen test (identifying those with higher starting viral densities) OR a positive PCR test for SARS-CoV-2 within the last 24hrs with a Ct value of less than 25 (all viral targets). A SARS-CoV-2 antibody RDT will be done and the result recorded (antibodies strongly correlate with higher viral loads (11)). Patients will be included who can be followed up reliably for 28 days (17-19). It is expected that vaccination coverage will increase during the study period as well as the cumulative number of natural infections.

The primary pharmacodynamic measure in this study is the rate of viral clearance (expressed as the slope of a fitted regression to the linear segment of the serial standardized oropharyngeal sample qPCR densities over 7 days) following treatment (5). Each site will include a no antiviral treatment control arm consisting of patients not receiving any antiviral treatment, although local hospital supportive treatment will remain unchanged for the patients including antipyretics, anti-tussives, antihistamines, vitamins etc and other required medications, in the clinical judgement of the treating Physician.

#### 5.1. Interventions

Intervention arms will be of three types:

A. Newly available and repurposed antiviral drugs. The initial list of intervention drugs is Hydroxychloroquine, Ivermectin, Lopinavir/Ritonavir, Fluoxetine, Fluvoxamine, Miglustat, Remdesivir, Nitazoxanide, Nebulised Unfractionated Heparin (UFH), Favipiravir, Molnupiravir and Nirmatrelvir/Ritonavir. Additional repurposed drugs can be added to the list. The interventions will be chosen in order of priority, as well as local feasibility at sites (availability of drugs, local EC and regulatory approvals).
B. Positive control: we will initially include the REGN-COV2 (monoclonal antibody cocktail); and later other antibodies when available.
C. Novel small molecule drugs: there are no candidates initially, but these can be added to the platform at any time after demonstrating success in phase 1 clinical trials.

At any given time in the study, it is possible that not all intervention arms are available. Randomisation ratios will be uniform across all available treatment arms within type A, B and the control arm in each site. If interventions of type C (novel drugs) are added to the platform, response adaptive randomisation will apply.

Randomisation will be continuously monitored by MORU. A positive SARS-CoV-2 lateral flow antigen test will be used as the main inclusion criteria. A positive lateral flow test result implies a relatively high viral load at the start of the study intervention (17-19). In order to assess viral clearance dynamics accurately over time initial high viral densities are required. In non-vaccinated patients, seronegativity implies there will be limited host control of the infection initially and was previously strongly correlated with higher viral loads (15), although this also depends on the variant.

We aim to identify interventions that accelerate viral clearance by a minimum of 12.5% (corresponding approximately to a reduction of 2 days in the mean time to viral clearance). Any intervention drug of type A or C that shows an acceleration of viral clearance, defined as a greater than 90% probability of accelerating viral clearance by more than 12.5%, or if there is evidence of accelerated viral clearance from other studies (e.g. Molnupiravir, Nirmatrelvir/ritonavir), will be selected for inclusion in a nested pharmacokinetic-pharmacodynamic study. In this nested study, frequent blood sampling will allow assessment of the relationships between plasma drug concentrations and viral clearance. The design of the pharmacokinetic study depends on the known pharmacokinetic properties of each drug. In each case no more than 10 blood samples (20mL) will be taken. Any intervention arm of type A, B or C that meets the futility endpoint (less than 10% probability of accelerating viral clearance by more than 12.5%) will be dropped from the study. New arms can be added at any time during the trial. Decisions concerning success and futility will be made sequentially at the pre-specified interim analysis to be conducted after 50 patients are enrolled, and thereafter after every 25 patients to ensure control of type 1 and type 2 error (Figure 4). The nested intensive PK-PD substudy will be triggered if the probability that viral clearance is accelerated by more than 12.5% goes above 90%, or if there is evidence of accelerated viral clearance from other studies (e.g. Molnupiravir, Nirmatrelvir/ritonavir).

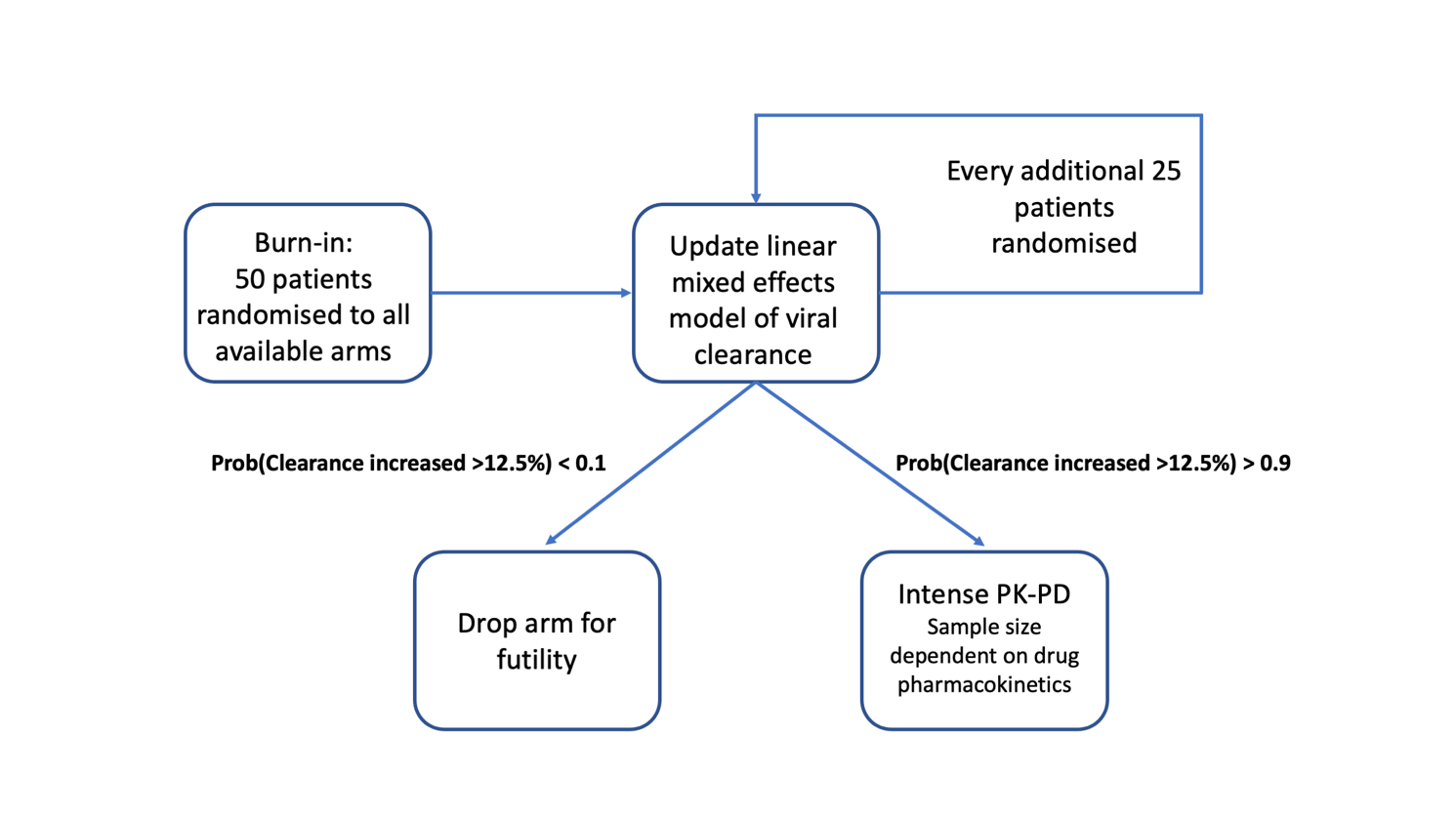

Figure 4. Flow diagram showing how each intervention arm can progress through the study. An interim analysis was conducted after qPCR data from the first 50 patients recruited into the study were available (burn-in). Ongoing internal monitoring will occur for every additional batch of 25 studied patients. Following the first interim analysis, the stopping rule threshold effect size was changed from 5% to 12.5 based on the observed effect of Regeneron and on sample predictions using observed uncertainty intervals. Futility occurs if the probability of success (increase in viral clearance > 12.5% relative to control) is less than 10%. Success occurs if the probability of success is greater than 90%, triggering an intensive pharmacokinetic-pharmacodynamic substudy for that arm.

### 6. PATIENT IDENTIFICATION AND RECRUITMENT

#### 6.1. Study Patients

Previously healthy adult patients with early symptomatic COVID-19.

#### 6.2. Inclusion Criteria

- Patient understands the procedures and requirements and is willing and able to give informed consent for full participation in the study.
- Previously healthy adults, male or female, aged 18 to 50 years at time of consent with early symptomatic COVID-19
- SARS-CoV-2 positive by lateral flow antigen test OR a positive PCR test for SARS-CoV-2 within the last 24hrs with a Ct value of less than 25 (all viral targets)
- Symptoms of COVID-19 (including fever, or history of fever) for less than 4 days (96 hours).
- Oxygen saturation ≥96% measured by pulse-oximetry at time of screening.
- Able to walk unaided and unimpeded in ADLs
- Agrees and is able to adhere to all study procedures, including availability and contact information for follow-up visits

#### 6.3. Exclusion Criteria

The patient may not enter the study if ANY of the following apply:

- Taking any concomitant medications or drugs (see appendix 4)†
- Presence of any chronic illness/ condition requiring long term treatment, or other significant comorbidity (see appendix 4 for the full list)
- Laboratory abnormalities discovered at screening (see appendix 4)
- For females: pregnancy, actively trying to become pregnant, or lactation
- Contraindication to taking, or known hypersensitivity reaction to any of the proposed therapeutics (see appendix 4)
- Currently participating in another COVID-19 therapeutic or vaccine trial
- Evidence of pneumonia (although imaging is NOT required)

† healthy women on the oral contraceptive pill are eligible to join the study

### 7. STUDY SET-UP AND PROCEDURES

Recruitment of patients will be from outpatient COVID-19/ ARI clinics or through other approved facilities, or from inpatient isolation facilities, or by patient self-referral to the study site. Patients with a positive SARS-CoV-2 rapid lateral flow antigen test OR a positive PCR test for SARS-CoV-2 within the last 24hrs with a Ct value of less than 25 (all targets) will be enrolled in the study after providing fully informed written consent. In each study site there will be a designated research ward for study procedures. In accordance with local and national infection control guidelines the initial assessments will be conducted in a designated research ward set-up to manage COVID-19 patients. From D1-D7, patients will receive care in either an inpatient or outpatient setting. All recruited patients will be visited at their home or place of residence, or be seen in the designated clinical trials unit or hospital ward. All staff will wear appropriate personal protective equipment (PPE). For home visits a trained member of staff will perform the swabbing (and may ask the patient to give a saliva sample).

The rapid lateral flow antigen test will be performed by trained study personnel. Patients will be eligible for the trial if the test is positive within a specified period of time (detailed in a separate SOP based upon manufacturer’s guidelines). A positive band appearing rapidly is correlated with a higher viral load in the swab sample (personal communication, Prof. Tim Peto). A digital photograph of the test result may be taken to allow for future analysis. The exact details of this will be detailed in the separate work instruction.

After enrolment into the study, the patient will be randomised to one of the study arms. Further details of the randomisation procedures are given below. An oropharyngeal swab will be obtained in duplicate by the study team using a standard operating procedure (SOP) after a full explanation is provided to the patient. Swabs will be in duplicate again prior to receiving the study drug. To minimise patient discomfort, all SARS-CoV-2 qPCRs will be performed on eluates from standard oropharyngeal swabs, taken in duplicate (20-23). In addition, some patients may be asked to give a sample of saliva (see section 7.1). The patient will undergo a physical examination and blood sampling for routine haematology and biochemistry and other baseline investigations (see Section 7.5). For oral study drugs, the first dose will be administered and observed at the research ward/ hospital. Subsequent doses will be administered at the patient’s home, or current residence, supervised by study personnel (if multiple doses, then one of the doses per day will be observed). In case a study drug requires parenteral administration, this will be provided under supervision by the study personnel in the research ward, or hospital, or the patient’s home.

#### 7.1.  Virological sampling

Serial oropharyngeal swabs will be taken in duplicate (one swab each tonsil) (24) according to the sample schedule. The swabs will be taken from the back of the throat (oropharynx; tonsillar fossa ) by trained study personnel according to the SOP abiding by strict PPE measures. Each swab will be placed in 3mL of viral transport medium (VTM) (Thermofisher M4RT®), snapped off and sealed. In addition, some patients may be asked to give a sample of saliva, either by spitting in a tube, or by sucking on a swab tip until it is saturated. The volume of saliva will be standardized. The initial purpose of this is to assess and validate a method of determining the viral density in the oropharynx in a less invasive manner or supplement the results from the oropharyngeal swabs to improve the characterization of the viral clearance. If these results are informative, they may subsequently be included in the SAP for endpoint analysis. Daily sampling will be performed by the study personnel who will place the swabs in the transport medium, and document the time. The samples will be kept cool, and subsequently stored at -80°C as detailed in the SOP. Transport times will be recorded, as well as the time at which the swab is frozen at -80°C. Viral genomes from the throat swabs will be quantitated by RT-qPCR, according to published methods (25). Each observed value will be recorded along with the RNAseP log_10_ density (representing human cell numbers). The exact details of the analysis are given in the SAP (26,27). The measurements for the patient over time will be used to estimate the rate of viral clearance under a Bayesian hierarchical linear mixed effects model. The concentration of urea in the VTM will also be measured and compared with the serum urea concentration in the baseline biochemistry sample. This comparison enables determination of the extracellular fluid content in the eluate volume of the oropharyngeal samples. The urea concentration in throat fluid is the same as in serum, so that the dilution factor of the throat sample in the VTM fluid can be easily calculated (28). Blood will also be taken for quantitative virology and other baseline assessments (see Section 7.5). All samples positive for SARS-CoV-2 on qPCR will be sequenced using Next Generation Sequencing (NGS), or canonical variant mutation analysis will be used to determine the virus genotype, in order to determine the effect of viral genetics on viral load and response to treatment.

#### 7.2. Recruitment

Potential patients with a positive SARS-CoV-2 test obtained during routine screening, or those with symptoms in keeping with COVID-19, even if they do not have the results of their SARS-CoV-2 test, or have not been tested yet, may be contacted by the research group. Alternatively they may contact the research group for participation in the study. Each will be provided with a written patient information sheet.

#### 7.3. Screening and Eligibility Assessment

Eligibility assessment will occur at the point of screening. If, based on the inclusion and exclusion criteria, the patient is eligible and willing to complete the full study, they will be included provided informed consent is obtained.

#### 7.4. Informed Consent

Written and verbal versions of the Patient Information and Informed Consent will be presented to the patients detailing the exact nature of the study; what it will involve for the patient; the implications and constraints of the protocol; and the known side effects of the medicines under evaluation and any risks involved in taking part. It will be stated clearly that the patient is free to withdraw from the study at any time for any reason without prejudice to future care, and with no obligation to give the reason for withdrawal.

The patient will be allowed as much time as required to consider the information as long as they remain eligible and within the time-frame for recruitment into the study, and the opportunity to question the Investigator or other independent parties to decide whether they will participate. Written Informed Consent will then be documented by a patient dated signature and dated signature of the person who presented and obtained the Informed Consent. The person who obtained the consent must be suitably qualified and experienced and have been authorised to do so by the site PI. A copy of the signed Informed Consent will be given to the patient. The original signed form will be retained at the study site.

#### 7.5. Baseline Assessments

A physical exam, vital signs, symptom review and basic demographics will be recorded. Oropharyngeal viral swabs taken by the study personnel in duplicate (swabs will be taken at two timepoints prior to study drug being administered) (and possibly a saliva sample see section 7.1). A total of 20 ml of venous blood for study testing will be taken at this point. Administration of the therapeutic may be delayed until the results of the FBC and Biochemistry will be available (although these will ideally be available within 1 hour of sample collection i.e. before randomization), to determine if there is any contraindication. If these bloods are not available in this timeframe, then the patient can be safely enrolled, randomized, and given their first dose, and can be excluded prior to their next dose if laboratory abnormalities resulting in a contraindication are present. If an FBC and Biochemistry has been taken in the previous 24hrs, the results of these can be used to determine whether there is a contraindication.

Laboratory tests at baseline

- Blood sample for
  - Full blood count (FBC) and differential count
  - Biochemistry (including U&Es, creatinine, LFTs and LDH)
  - Fibrinogen, Ferritin, TNF/IL-6/IL-1
  - Serology (antibody responses)
  - Stored sample
  - Drug level
  - Host genotyping to assess determinants of the therapeutic response
- Oropharyngeal swab in duplicate (and possibly a saliva sample see section 7.1)
- ECG

#### 7.6. Days of study

7.6.1 Randomisation D0

After enrolment, the patient will be randomized to one of the study arms using an online randomization application. The number of arms at a site can be limited by the approval and availability of the study drug at that site. Randomization to the no antiviral treatment control arm (no intervention) will be fixed at a minimum of 20% throughout the study. The randomization ratios will be uniform for all available interventions. The drug allocations are not blinded for the patient for practical reasons. For patients randomized to an active drug, the first treatment dose (hour 0) will be supervised by study team after a viral swab (and possibly a saliva sample see section 7.1) is taken again for qPCR (taken in duplicate) (see appendix 1 for schedule of activities). The patient will then be observed for a minimum of 1 hour in the ward.

For the therapeutic arms for which intensive PK has been triggered, allocated patients will stay for extra blood tests for drug monitoring levels, according to the schedule in the drug-specific appendix. For further details of the intensive PK schedules for the individual therapeutics please see the appendix 2.

7.6.2 Day 1- day 7

The following variables will be assessed daily:

- Eligibility check, and assessment of treatment (taking of medications taken will be observed by the study personnel and the time recorded. In participants who are outpatients, for medications taken more than once daily, the patient will record the time non-observed doses are taken).
- Oropharyngeal swab for qPCR taken in duplicate (and possibly a saliva sample see section 7.1) by the study personnel (time taken recorded by the study personnel)
- Temperature recorded twice daily by study staff or participant (information collected in a diary and reviewed by the study personnel daily)
- Study personnel completes a brief symptom questionnaire each day
- Two blood samples will be taken (ideally day 3 and 7) for antibody levels and for later pharmacokinetic analyses (pre- or post-dose depending on the therapeutic in question) and FBC and biochemistry, to assess potential drug adverse effects.

7.6.3 Day 14

All patients will have oropharyngeal swabs taken in duplicate and a blood test taken on day 14 (and possibly a saliva sample see section 7.1) either at home (if feasible) or at the research ward to follow-up blood abnormalities, if identified in the earlier tests, and to determine antibody responses (serology).

7.6.4 Day 28 assessment

The day 28 follow-up assessment will be through a telephone call or a visit to check whether the patient recovered uneventfully for whether there was progression to more severe illness or need for hospitalization for clinical reasons. This final assessment will have an allowed window -2/ +7 days based upon calculation from Day 0.

#### 7.7. Intensive pharmacokinetic sampling:

For those study drugs that are shown to be effective antivirals in other studies (e.g. Molnupiravir, Nirmatrelvir/ritonavir) or meet the success criteria of the study (defined as a greater than 90% probability of accelerating viral clearance by more than 12.5%), we will then conduct intensive pharmacokinetic (PK) sampling for further patients who are enrolled into that arm. The exact sampling schedule will depend on the study drug characteristics (see appendix 2), but in general the patient will stay at the ward for the remainder of the day to have further blood tests performed. A cannula will be inserted so that blood can be drawn without the need for repeated venepuncture. A further blood test will be performed the next day, either at the ward or at their home if feasible. The volume of blood from this will not exceed 20 mls.

#### 7.8. Management of patients who become ill

During the study, although this is unlikely, the patient may deteriorate clinically from their COVID-19 or may develop a new intercurrent illness or potentially a side-effect related to the study medication.

All patients who develop difficulty with activities of daily living or complain of shortness of breath will be assessed initially by the study personnel and their oxygen saturation measured and, if necessary, brought to the clinic research physician for further assessment. Based on this assessment, they may be referred to hospital, re-examined the next day, or asked to update the research team by mobile phone frequently on their well-being.

If a patient is referred to hospital, a clinical assessment will be made by the hospital physician and a diagnosis made. The decision to stop study drug will depend on that diagnosis and be made between the hospital team and the study PI.

The hospital physicians will have the responsibility for patient care but the research team will continue to follow the progress of the patients in hospital. Treatment for COVID-19 severe enough to warrant hospitalization will follow national guidelines. For those patients where treatment is changed (i.e. a new treatment is started for clinical reasons) an extra set of swabs in duplicate (and possibly a saliva sample see section 7.1) should be collected prior to the initiation of the new treatment (i.e. 2 sets of swabs will be taken in duplicate). This will only occur if the patient is within the first 7 days of the study.

The D0, D3, D7 and D14 blood samples will be tested for FBC and biochemistry (e.g. U&Es, LFTs) to assess for drug-related adverse effects or abnormalities which require a potential dose adjustment or stopping the therapeutic or clinical intervention. The responsibility for evaluating and acting upon the results of blood tests will belong to the study team at the local site. They will use their clinical judgement in the assessment of the patient, further tests and onwards referral. At any point of the study, extra laboratory tests may be conducted on the patient if felt to be clinically indicated by the research team, based on symptoms or previous laboratory abnormalities.

#### 7.9. Sample Handling and Retention

Samples will be transferred to designated testing facilities, where they will undergo testing in accordance with best practice laboratory measures and safety procedures.

Oropharyngeal swabs (and possibly a saliva sample see section 7.1) will be processed using validated quantitative Real-Time quantitative Polymerase Chain Reaction (qPCR) to detect SARS-CoV-2, according to the study SOP. A urea measurement will be measured on each and compared to the baseline blood concentration. Swabs will be tested in duplicate.

The samples will be retained as per University of Oxford and local site regulations. Consenting patients may rescind their consent up until the completion of the study. Unless requested the samples collected prior to date of withdrawal will be retained for study analysis.

Samples, including those for PK, may be transferred to MORU or other designated testing facilities outside the site country, with appropriate material transfer agreements (MTA) and associated approvals prior to shipment.

### 8.  Discontinuation/Withdrawal of Patients from Study

Patients may choose to stop treatment and/or study assessments but may remain on study follow-up. Patients may also withdraw their consent, meaning that they wish to withdraw from the study completely. In the case of withdrawal from both treatment and active follow up the following options for a tiered withdrawal from the study would apply a) Patients withdraw from the study but permit data and samples obtained up until the point of withdrawal to be retained for use in the study analysis. No further data or samples would be collected after withdrawal, b) Patients withdraw from active follow-up and further communication but allow the trial team to continue to access their medical records and any relevant hospital data that is recorded as part of routine standard of care; i.e., CT-Scans, blood results and disease progression data etc.

In addition, the Investigator may discontinue a patient from the trial treatment at any time if the Investigator considers it necessary for any reason including, but not limited to:

- Pregnancy
- Ineligibility (either arising during the study or retrospectively having been overlooked at screening)
- Significant protocol deviation
- Significant non-compliance with treatment regimen or trial requirements
- An adverse event which requires discontinuation of the trial medication or results in inability to continue to comply with trial procedures
- Disease progression which requires discontinuation of the trial medication or results in inability to continue to comply with trial procedures

The reason for discontinuation and/or withdrawal will be recorded in the Case Report Form. qPCR data from patients withdrawn from the study will still be analysed if at least three distinct timepoints are available for the estimation of a clearance slope. The sample size is adaptive so there is no need to replace withdrawn patients.

Consenting patients may rescind their consent up until the completion of the study. Unless requested, data and samples collected prior to the date of withdrawal will be retained in the study database and analysis.

#### 8.1. Definition of End of Study

The end of study is the date of the 28 day follow up visit of the last enrolled patient.

### 9. SAFETY REPORTING

#### 9.1. Definitions

| Adverse Event (AE) | Any untoward medical occurrence in a patient to whom a medicinal product has been administered, including occurrences which are not necessarily caused by or related to that product. |
| --- | --- |
| Adverse Reaction (AR) | An untoward and unintended response in a patient to an investigational medicinal product which is related to any dose administered to that patient.  The phrase “response to an investigational medicinal product” means that a causal relationship between a trial medication and an AE is at least a reasonable possibility, i.e. the relationship cannot be ruled out.  All cases judged by either the reporting medically qualified professional or the Sponsor as having a reasonable suspected causal relationship to the trial medication qualify as adverse reactions. |
| Serious Adverse Event (SAE) | A serious adverse event is any untoward medical occurrence that:   - results in death - is life-threatening - requires inpatient hospitalisation or prolongation of existing hospitalisation^ş^ - results in persistent or significant disability/incapacity - consists of a congenital anomaly or birth defect.   Other ‘important medical events’ may also be considered serious if they jeopardise the patient or require an intervention to prevent one of the above consequences.  NOTE: The term “life-threatening” in the definition of “serious” refers to an event in which the patient was at risk of death at the time of the event; it does not refer to an event which hypothetically might have caused death if it were more severe.  Hospitalisation is defined as an unplanned, formal inpatient admission, even if the hospitalisation is a precautionary measure for continued observation. The patient must be admitted overnight, a short stay of several hours to receive treatment is not considered hospitalisation. If a patient is admitted overnight or longer for social/economic or isolation reasons and is otherwise medically stable, this does not constitute a SAE. Other examples of visits to a hospital facility that are not considered hospitalisation are: Emergency room visits, outpatient surgery, pre-planned or elective procedures for a pre-existing condition (as long as that condition has not deteriorated while on trial treatment or brought forward because of worsening symptoms) and for the purpose of this study, being hospitalised for COVID-19 isolation. |
| Serious Adverse Reaction (SAR) | An adverse event that is both serious and, in the opinion of the reporting Investigator, believed with reasonable probability to be due to one of the trial treatments, based on the information provided. |

NB: to avoid confusion or misunderstanding of the difference between the terms “serious” and “severe”, the following note of clarification is provided: “Severe” is often used to describe intensity of a specific event, which may be of relatively minor medical significance. “Seriousness” is the regulatory definition supplied above.

Any pregnancy occurring during the clinical trial and the outcome of the pregnancy should be recorded and followed up for congenital abnormality or birth defect, at which point it would fall within the definition of “serious”.

#### 9.2. Causality

The relationship of each adverse event to the trial medication must be determined by a medically qualified individual according to the following definitions:

Definitely related:	There is clear evidence to suggest a causal relationship and other possible contributing factors can be ruled out.

Probably related:	There is evidence to suggest a causal relationship and the influence of other factors is unlikely.

Possibly related:	There is some evidence to suggest a causal relationship (e.g. because the event occurs within a reasonable time after administration of the trial medication). However, the influence of other factors may have contributed to the event (e.g. the patient’s clinical condition, other concomitant treatments).

Unlikely to be related: There is little evidence to suggest there is a causal relationship (e.g. the event did not occur within a reasonable time after administration of the trial medication), or there is another reasonable explanation for the event (e.g. the patient’s clinical condition, other concomitant treatment).

Not related:	There is no evidence of any causal relationship.

#### 9.3. Procedures for Recording Adverse Events

A symptom questionnaire will be performed daily until D7 with a final follow-up assessment on D28 to aid in the identification of adverse events. The severity of adverse events will be assessed following the Common Terminology Criteria for Adverse Events (CTCAE) v5.0:

1 = mild, 2 = moderate, 3 = severe, 4 = life-threatening, 5 = fatal.

Aes occurring in patients from enrolment and during trial participation that are observed by the Investigator or reported by the patient with severity **grade of 3 (severe) or higher** will be recorded on the CRF, whether or not attributed to trial medication.

The following information will be recorded: description, date of onset and end date, severity, assessment of relatedness to trial medication, other suspect drug or device and action taken. Follow-up information should be provided as necessary.

Aes considered related to the trial medication as judged by a medically qualified investigator or the Sponsor will be followed either until resolution, or the event is considered stable.

It will be left to the Investigator’s clinical judgment to decide whether or not an AE is of sufficient severity to require the patient’s removal from treatment. A patient may also voluntarily withdraw from treatment due to what he or she perceives as an intolerable AE. If either of these occurs, the patient must undergo an end of trial assessment and be given appropriate care under medical supervision until symptoms cease, or the condition becomes stable.

#### 9.4. Grading of laboratory abnormalities

Abnormal laboratory findings detected during the study or are present at baseline and worsen following the taking of study medication will be reported as Aes or SAEs. If found to be abnormal the values will be graded according to the CTCAE v5.0. Aes and SAEs of severity grade 3 or higher will be recorded in the CRF and will be followed up until grade 2 or below, returned to baseline, or deemed to be permanent. For laboratory results that are not available in the CTCAE v5.0, the site investigator should make a determination as to whether or not the laboratory abnormality is clinically significant. If the site investigator believes a laboratory abnormality is clinically significant, it should be reported as an adverse event and a grade should be identified to the best of their ability.

#### 9.5. Reporting Procedures for Serious Adverse Events

All SAEs detected by the site investigator must be reported to the PLATCOV safety team within 24 hours of site awareness. The Serious Adverse Event CRF documenting the SAE should be emailed to. The PLATCOV safety team will inform the DSMB within 10 days of initial notification of the SAE and keep the DSMB updated as needed.

Further reports should be submitted, if required, until the SAE is resolved, is deemed stable/ permanent or results in death. A final status should be determined for any SAEs ongoing at the study end date.

The site PI must also report the SAEs to the local ethics committee and the regulatory authority in accordance with local requirements.

9.5.1 Expectedness

Expectedness will be determined according to the Investigators’ Brochure/Summary of Product Characteristics.

9.5.2 SUSAR Reporting

All SUSARs will be reported by the site PI to the relevant Competent Authority and to the local Ethics Committee and other parties as applicable. For fatal and life-threatening SUSARS, this will be done no later than 7 calendar days after MORU is first aware of the reaction. Any additional relevant information will be reported within 8 calendar days of the initial report. All other SUSARs will be reported within 15 calendar days.

Site Pis will be informed of all SUSARs for the relevant IMP that occur at any of the PLATCOV study sites.

#### 9.6. Data Safety Monitoring Board (DSMB)

An independent Data Safety and Monitoring Board (DSMB) will be set up consisting of qualified volunteers with the necessary knowledge of clinical trials. The DSMB will receive summary reports from MORU as defined per charter or per ad-hoc request, prior to each meeting. All data reviewed by the DSMB will be in the strictest confidence. A DSMB charter will outline its responsibilities and how it will operate.

An interim report will be prepared by the Trial Statistician for the pre-specified interim analysis. In case of safety concerns, additional information or formal interim analyses can be requested by the DSMB.

The DSMB will meet formally at the following timepoints:

- before the study starts
- after the first 50 patients have been accrued into the study (10 per arm)
- At additional time-points as indicated by the DSMB after their review, if deemed necessary

All DSMB recommendations will be communicated to site Pis. The site PI will be responsible for submitting the written DSMB summary reports with recommendations as applicable to local/ national ethics committees and other applicable groups.

### 10. STATISTICS AND ANALYSIS

#### 10.1. Overview of adaptive study design and overall approach

This is a platform trial that will evaluate multiple antiviral treatments for COVID-19. The sample size is adaptive with multiple planned interim analyses. By specifiying an adaptive sample size, we can more rapidly identify treatments that work and possibly remove treatments that do not work. Clinical trials with adaptive designs are increasingly being used in research, including in such successful and prominent COVID-19 studies as the RECOVERY study. In addition the FDA and EMA supply guidance on studies with these designs^[1]^. They do not have fixed sample sizes as conventional RCTs. The number of patients recruited depends on the results (i.e. they are adaptive).The benefits of an adaptive sample size are increased efficiency and ethical considerations (i.e. they can stop ineffective arms earlier, identify effective arms earlier, and limit numbers in ineffective arms compared to fixed sample number studies). Because of these advantages this type of study is likely to become more prevalent, but it is important that the study is well-designed in advance, that the adaptive design is appropriate for the overall study design, and has mechanisms in place to control type I error.

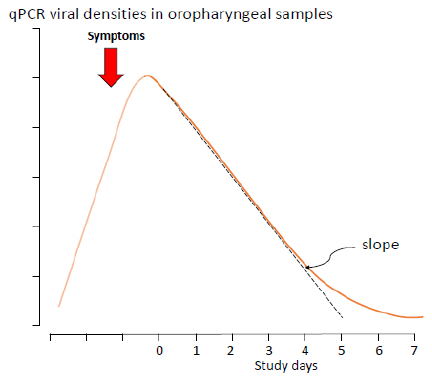
The main pharmacodynamic endpoint is the rate of viral clearance, estimated from serial qPCR measurements in each patient (4). There is considerable inter-individual variability in clearance rates which are first order and predominantly monoexponential. The measured end point is the slope of the the initial log linear decline in qPCR estimated viral densities (Figure 5). The clearance rate is correlated with both disease progression and the presence of symptoms. As this is a multi-centre trial, populations may differ between sites with respect to key covariates. Therefore, the main analytic approach will be to fit a hierarchical Bayesian model to the serial qPCR measurements (using default weakly informative Bayesian priors to aid model fitting), whereby the baseline viral loads (intercept) and the first-order clearance rates (slope of initial decline) can vary between sites and between individuals. The viral clearance slope is also dependent on the randomised study arm. Stopping an intervention arm (of type A, B or C) for futility or triggering an intensive PK-PD nested sub-study (type A or C) will be decided using the posterior probabilities that the arm results in an increased slope (increase set as 12.5%, following the results of the first interim analysis). The PK-PD nested sub-study will be also triggered if there is evidence of accelerated viral clearance from other studies (e.g. Molnupiravir, Nirmatrelvir/ritonavir).

Figure 5 qPCR viral densities in oropharyngeal samples over time, showing time of likely symptom onset relative to viral densities and study days

#### 10.2. Results of the first interim analysis

As pre-specified in the SAP, the first interim analysis, triggered once data from the first 50 patients were available, was presented to the Trial Steering Committee and the DSMB. This analysis showed that the positive control arm (REGN-CoV-2) had a substantially larger effect on viral clearance than anticipated, with over 50% increase in viral clearance relative to the negative control arm (Figure 3). In addition, the intra-individual variation in viral load estimation was greater than anticipated, resulting in larger uncertainty around effect sizes. For these reasons, we changed the threshold effect size defining futility and success from 5% increase in viral clearance to 12.5% increase in the viral clearance. Under this threshold, we would need an average of 50 patients randomised to an intervention that has no effect on viral clearance to demonstrate futility.

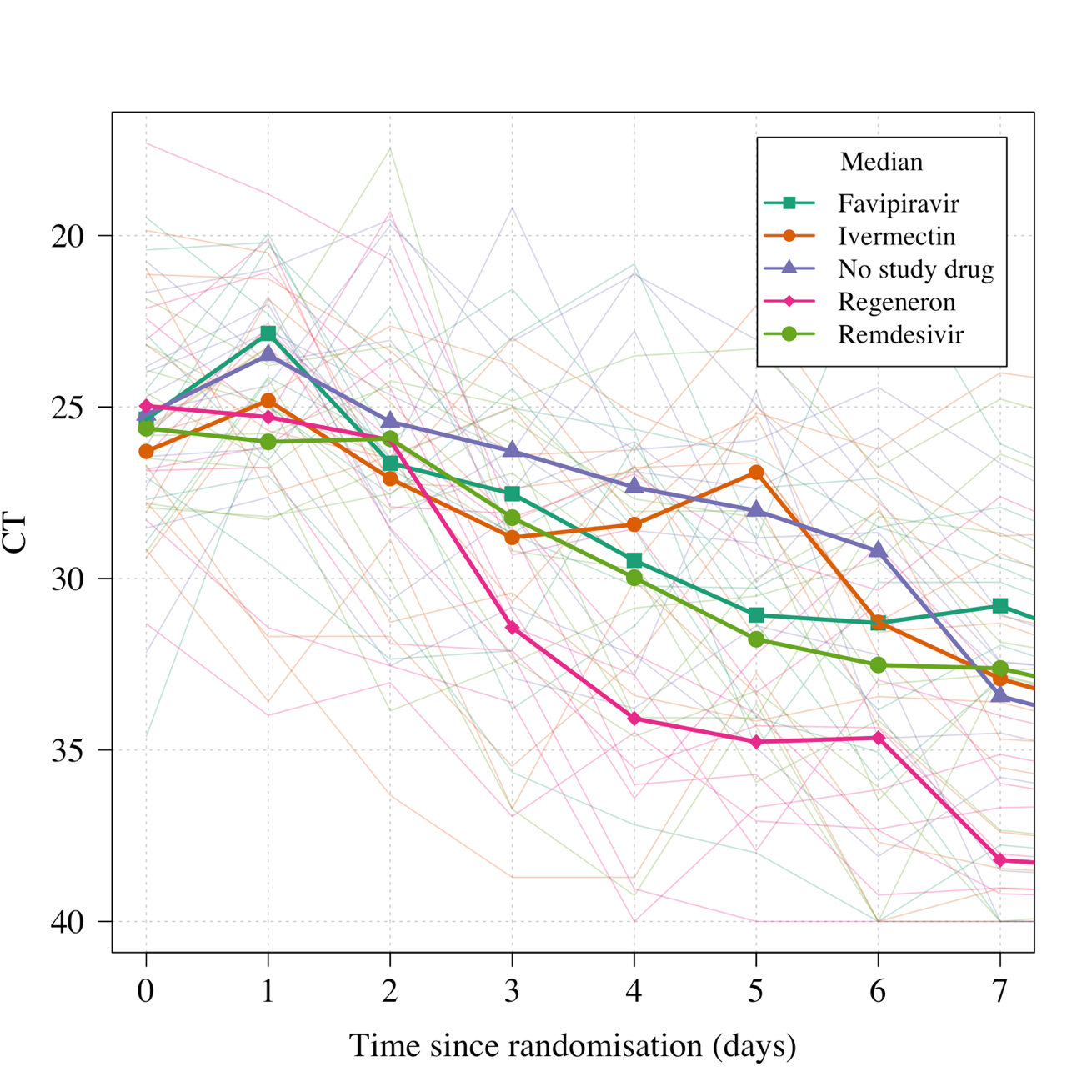

Figure 3 Data from the first interim analysis (first 50 patients recruited into the study).

#### 10.3. Bayesian hierarchical model of viral clearance

At each interim analysis we will fit a series of Bayesian hierarchical (mixed effects) model to the serial viral load measurements (modelled directly on the Ct scale). The main linear model will take the following form:

y_k,i,t_ ~ Student(α_0_ + α_k_ + α_i_ + (β_0_ + β_i_ + β_k_ + β_T(i)_)t , sigma^2^, dof)

where y_k,i,t_ is the observed log_10_ viral load for individual i at time t from site k (two values per timepoint), and T(i) is the randomised arm assigned to individual i. Sigma^2^ is the measurement error (residual error). The error model is a t-distribution with degrees of freedom inferred from the data (dof). The slope of viral clearance decomposes into 4 terms: the population mean slope β_0_; the site random effect β_k;_ the individual random effect β_i_; and the treatment effect (fixed across sites and individuals) β_T(i)_. The intercept term (baseline viral load) decomposes into 3 terms: the population intercept α_0_; the site random effect α_k_; and the individual random effect α_i_. All comparisons are made relative to the no antiviral treatment control arm, so we set β_control_ = 0. Adjustment will be made for human RNaseP concentration. The model will treat viral load corresponding to CT values of 40 as left censored. An equivalent non-linear model (allowing for an initial increase in viral load, followed by a linear decrease) will also be fitted to the data as a sensitivity analysis.

See the SAP (version 2.1) for full details of the analysis.

#### 10.4.  Time varying effects

Characterising and identifying time varying effects not specified in the main model is important for two reasons:

- Monoclonal antibodies (type B- see study design section)- where the efficacy can be reduced in infections with spike protein mutant SARS-CoV-2 strains. Therefore, it is possible that a waning effect is observed over time justifying early stopping.
- New interventions (type C- see Study design section) which are added later on during the trial cannot be compared directly to all data from the control arm without accounting for possible differences in population over time (as an epidemic evolves, included populations may differ in a single site over time).

We will use descriptive approaches to detect time-varying effects (residual plots over time, mean slopes in control individuals over time). If substantial time-varying effects are observed, we will add a smooth time varying spline component to the linear model. The base model will not include this for simplicity.

#### 10.5. Sample size estimation, randomisation and statistical considerations for the Bayesian group sequential design

The platform study has two main objectives:

- Identify interventions of type A or B that are unlikely to be of clinical benefit (i.e. demonstrate futility)
- Identify interventions of type A or C that could have clinical benefit and determine optimal dosing (from the nested PK-PD study) for phase 3 evaluations. For type C, these will have stronger *a priori* justification and choosing the best candidate is desired (rather than choosing all candidates with reasonable chance of success)

The required sample size depends on how stringent the thresholds are that define futility or success. Before the trial started we used simulation to determine the probability thresholds that result in control of the type 1 error at 10% and control of the type 2 error at 20% using as a minimum effect size a 5% increase in viral clearance. Following the first interim analysis at 50 patients, the minimum threshold was changed to 12.5%

For the following interim analyses, the decisions mde for a treatment arm *T* are as follows:

- Stopping for futility: if Probability(β_T_ > 0.125 * β_0_) < 0.1
- Triggering intensive PK-PD nested sub-study: if Probability(β_T_ > 0.125 * β_0_) > 0.9

The first interim analysis was carried out after 50 patients were enrolled. We will then carry out interim analyses every 25 patients. The initial thresholds for futility and success were chosen after simulating 6000 trials with all combinations of futility thresholds in the set {0.05, 0.1, 0.2} and success thresholds in the set {0.8, 0.9, 0.95, 0.99} (12 combinations in total, 500 simulations per combination). The maximum sample size was set at 400 patients in total with 6 arms (5 intervention arms and 1 control arm). Out of the 5 interventions, 1 was effective with an effect size of 10% and the other 4 were ineffective (no increase in slope). Type 1 error for a particular combination of futility and success thresholds was calculated as the average number of times any of the ineffective arms were declared effective divided by the number of decisions (note this the per decision type 1 error not the family wide error rate which is dependent on the number of arms). The type 2 error was calculated as the number of times the effective arm was declared ineffective or no decision was reached by 400 patients. Appendix 3 shows how type 1 and type 2 error varies across choices of thresholds. The thresholds we have chosen allow us to control type 1 errors at 10% and type 2 errors at 20%.

For these futility and success thresholds, the simulations demonstrate the number of patients randomised per arm in order to reach a decision is:

- 50 patients (average; median is 30) for each efficacious intervention (effect size of 10% increase in viral clearance)
- 40 patients (average; median is 25) for each inactive intervention (effect size of 0)

Therefore, supposing there were 5 interventions of type A in the platform, and assuming that only 1 of the 5 were in fact effective, we would need on average a total of 50 + 4*40 = 210 patients randomised to interventions of type A (approximately 66% of the total sample size: a minimum of 20% of samples are randomised to the negative control and thus ~13% of patients are randomised to each intervention arm and the positive control arm). This implies a total of ~320 patients for the first set of interventions identified (this does not include any intensive PK sub-studies).

This study is open label and so a centralized server-based randomization system will be used. Only designated study staff will be authorized to access the webapp for patient allocation. The webapp will be password protected. All randomisation activities will be traced within the webapp and attributed to a timestamp along with the anonymised patient study code, age and sex.

#### 10.6. Sample size for the intensive PK-PD sub-study

Any drugs of type A or C that with evidence of accelerated viral clearance from other studies (e.g. Molnupiravir, Nirmatrelvir/ritonavir) or which meet the success endpoint (>90% probability that clearance is accelerated by more than 12.5% relative to control) will be included in an intensive PK-PD study. The sample size calculation for each sub-study will be drug dependent and will use the information regarding the variability in clearance in the intervention arm versus the variability in clearance in the control arm (i.e. it will be determined by the results). Larger observed variability in the intervention arm would be expected if an active intervention had variation in exposures that corresponded to variation around the near maximal effect concentration (eg EC_95_).

We will use a PK simulation-based approach where we will simulate predicted drug AUC values for patients given the chacteristics of the patients in each site (weight and sex) and use the observed variability in viral clearance rates to estimate the necessary sample size to infer a dose-response relationship whereby a 10% increase in AUC resulted in a 10% increase in the slope of viral clearance with 80% power and 5% type 1 error.

#### 10.7. Final analysis of primary outcome

The final analysis will fit a series Bayesian hierarchical linear and non-linear models to the serial log_10_ viral load measurements (copies per mL). The primary estimates of interest are the relative changes in the rate of viral clearance for the intervention arms (including the positive control arm) compared to the no study drug arm. These will be reported as their mean posterior estimates along with credible intervals. All analyses will be done using *R* using the package *rstan* along with bespoke software. All analysis code is openly available via a github repository: <https://github.com/jwatowatson/PLATCOV-SAP>. Anonymised viral load data and key meta data (e.g. site, strata, randomised allocation) will be made available at publication of results.

### 11. DATA MANAGEMENT

#### 11.1. Access to Data

Direct access will be granted to authorised representatives from the University of Oxford and any host institution for monitoring and/or audit of the study to ensure compliance with regulations. Outcome data and treatment assignment data will be made available for analysis in real time.

#### 11.2. Data Handling and Record Keeping

Clinical study data will be recorded on CRFs and entered on to a password-protected database by the local study PI, research nurse or designee. The study database will be built in MACRO EDC, a clinical data management system that is compliant with ICH GCP and FDA 21 CFR Part 11 and will be hosted in a secure, access-restricted server in MORU. The study database will include internal quality checks to identify data that appear inconsistent, incomplete, or inaccurate.

Measures will be taken to ensure non-disclosure of information that is potentially harmful to patients. Paper records (for example, patient identifiable information for the purposes of follow-up, the screening logs and signed ICFs) will be kept in locked cabinets; electronic data will only be accessible to staff with user accounts and passwords. The database contains an audit trail that keeps record of changes to data and user activity within the database. All electronic data will be stored on secure servers that are backed up daily, with weekly off-site storage.

Patient records at site will, taking into account the ability of the sites, be stored in binders in the secured access-limited room or scanned and stored electronically. The records will be retained for five years following completion of the study, or according to local site regulation. The study database will be retained indefinitely.

With patient’s consent, clinical data and results from blood analyses stored in the database may be shared according to the terms defined in the MORU data sharing policy with other researchers to use in the future.

Data generated from this study will adhere to the 2016 “[Statement on data sharing in public health emergencies](https://wellcome.ac.uk/what-we-do/our-work/statement-data-sharing-public-health-emergencies)”(<https://wellcome.ac.uk/press-release/statement-data-sharing-public-health-emergencies>).

### 12. QUALITY CONTROL AND QUALITY ASSURANCE PROCEDURES

The study will be conducted in accordance with relevant regulations and standard operating procedures.

The study will be conducted in compliance with this protocol, International Conference on Harmonization (ICH) Guidelines for Good Clinical Practice (GCP) and any applicable regulatory requirement(s). Monitoring will be overseen by the MORU Clinical Trials Support Group (CTSG) according to a prespecified risk-based monitoring plan to ensure compliance to the study protocol and applicable guidelines and regulations. Biological specimens will be processed, stored and shipped in accordance with MORU SOPs.

Data validation will be performed to identify errors or discrepancies and thus ensure completeness, validity and accuracy of data.

### 13. Ethical and regulatory considerations

This study will be conducted in patients who would be unlikely to progress to severe illness. The investigated medications may benefit the patient; i.e. shorten the duration of symptoms or decrease their severity, but are unlikely to have a significant adverse effect on the patient’s illness and subsequent health. No drug intervention has been shown to be unequivocally beneficial at this point in the illness, in this population, and clinical equipoise exists between the interventions and the no antiviral treatment (although local supportive treatment will be fully provided including antipyretics (e.g. paracetamol), anti-tussives, antihistamines, vitamins etc and any treatments clinically indicated by the Physician looking after the patient. Funds will be set aside to cover hospital costs in the unlikely event of a drug adverse reaction.

Women who are pregnant, actively trying to become pregnant, or breast feeding will be excluded from this study as it is not known if any of the treatments being tested will have additional benefits that would outweigh any risks associated with pregnancy/breast feeding.

#### 13.1. Declaration of Helsinki

The Investigator will ensure that this study is conducted in accordance with the current revision of the Declaration of Helsinki.

#### 13.2. Guidelines for Good Clinical Practice

The Investigator will ensure that this study is conducted in accordance with relevant regulations and with Good Clinical Practice.

#### 13.3. Approvals

The protocol, informed consent form and participant information sheet will be submitted to OxTREC and local ethics committees for written approval.

The Investigator will submit and, where necessary, obtain approval from the above parties for all amendments to the original approved documents.

#### 13.4. Patient Confidentiality

The study staff will ensure that the patients’ anonymity is maintained. The patients will be identified only by a patient ID number on all study documents and any electronic database, with the exception of the CRF, where patient initials may be added. The name and any other identifying detail will NOT be included in any study data electronic file. All documents will be stored securely and only accessible by study staff and authorised personnel. The study will comply with the Data Protection Act 2018, which requires that personal data must not be kept as identifiable data for longer than necessary for the purposes concerned.

#### 13.5. Expenses

Patients will be financially compensated for their time in the study, in accordance with local EC guidance and approval.

#### 13.6. Risk

All of the treatments being tested *initially (see Appendix 2 for study drugs)* are generally well tolerated with the main adverse effects related to gastrointestinal symptoms (abdominal pain, diarrhoea, nausea and vomiting). In the case of intravenously/subcutaneously administed treatments there may be discomfort, bleeding or bruising of the skin at the site of needle puncture. Further information regarding specific side effects for each of the drugs used in the study can be found in Appendix 2. These are usually mild and settle without the need for medical intervention.

The risks associated with blood withdrawl during the study include discomfort, occasional bleeding or bruising at the site of needle puncture, and very rarely infection. The risks associated with nose and throat swabbing are limited to some slight discomfort.

#### 13.7. Benefits

It is not yet known if any of the treatments being tested will have additional benefits for the patient in the management of COVID-19. Although an individual patient may not personally benefit, this study should help future COVID-19 patients by discovering treatments that work in early disease and ruling out those do not. Remuneration will be provided to the patients for the period they are enrolled in the study. Patients will be reimbursed for costs associated with traveling to the study site and loss of work time. The amount will be determined per local allowed guidelines and ethics committee policies .

#### 13.8. Reporting

The PI shall submit an Annual Progress Report to OxTREC on the anniversary of the date of approval of the study. In addition, the PI shall submit an End of Study Report to OxTREC within 12 months of completion of the study.

#### 13.9. Finance and insurance

13.9.1 Funding: ACT- Accelerator

This trial is funded by the COVID-19 Therapeutics Accelerator (CTA), managed with MORU through the Wellcome Trust. CTA is a philanthropic collaboration supporting efforts to research, develop and bring effective treatments against COVID-19 to market quickly and accessibly. CTA and Wellcome Trust have had no role in the design of this study and will not have any role during its execution, analyses, interpretation of the data, or decision to submit the results.

13.9.2 Insurance:

The University has a specialist insurance policy in place which would operate in the event of any patient suffering harm as a result of their involvement in the research (Newline Underwriting Management Ltd, at Lloyd’s of London).

#### 13.10. Data ownership

The data generated in this study belongs to the study group as a whole. The final database will be shared amongst the site PI and key members of the research team.

The database may be shared with researchers not directly involved in this study but only after the main paper has been published and in accordance with MORU guidelines on data sharing. The database will only be shared if future publications are not compromised. The criteria for authorship will be consistent with the international guidelines (<http://www.icmje.org/#author>).

#### 13.11. Publication policy

The Investigators will be involved in reviewing drafts of the manuscripts, abstracts, press releases and any other publications arising from the study. Authors will acknowledge that the study was funded by <*insert name*>. Authorship will be determined in accordance with the International Committee of Medical Journal Editors (ICMJE) guidelines and other contributors will be acknowledged.

The results of the study will be summarised in lay language, in both English and the language(s) commonly spoken at the study sites, and disseminated to key stakeholders, user communities and patients.

### 14. References

1. Organisation WH. COVAX delivers its 1 billionth COVID-19 vaccine dose 2022 [Available from: COVAX delivers its 1 billionth COVID-19 vaccine dose (who.int).
2. He X, Lau EHY, Wu P, Deng X, Wang J, Hao X, et al. Temporal dynamics in viral shedding and transmissibility of COVID-19. Nat Med. 2020;26(5):672-5.
3. Wölfel R, Corman VM, Guggemos W, Seilmaier M, Zange S, Müller MA, Niemeyer D, Jones TC, Vollmar P, Rothe C, Hoelscher M, Bleicker T, Brünink S, Schneider J, Ehmann R, Zwirglmaier K, Drosten C, Wendtner C. Virological assessment of hospitalized patients with COVID-2019. Nature. 2020; 588 (7839):E35. May;581(7809):465-469.
4. Hillen T, Contreras C, Newby JM. Personalized Virus Load Curves of SARS-CoV-2 Infection medRxiv preprint doi: <https://doi.org/10.1101/2021.01.21.21250268>
5. Watson JA, Kissler SM , Day NPJ, Grad Y, White NJ. Optimal design for phase 2 studies of SARS-CoV-2 antiviral drugs medRxiv preprint doi: <https://www.medrxiv.org/content/10.1101/2021.01.06.21249368v1.full.pdf>
6. Gastine S, Pang J, Boshier FAT 2, Carter SJ, Lonsdale DO, Cortina-Borja M, Hung IFN, Breuer J, Kloprogge F, Standing JF. Systematic review and patient-level meta-analysis of SARS-CoV-2 viral dynamics to model response to antiviral therapies. medRxiv preprint doi: https://doi.org/10.1101/2020.08.20.20178699
7. Wang D, Hu B, Hu C, Zhu F, Liu X, Zhang J, et al. Clinical Characteristics of 138 Hospitalized Patients With 2019 Novel Coronavirus-Infected Pneumonia in Wuhan, China. JAMA. 2020. 323(11):1061-1069.
8. RECOVERY Collaborative Group, Horby P, Lim WS, Emberson JR, Mafham M, Bell JL, Linsell L, Staplin N, Brightling C, Ustianowski A, Elmahi E, Prudon B, Green C, Felton T, Chadwick D, Rege K, Fegan C, Chappell LC, Faust SN, Jaki T, Jeffery K, Montgomery A, Rowan K, Juszczak E, Baillie JK, Haynes R, Landray MJ. Dexamethasone in Hospitalized Patients with Covid-19 - Preliminary Report. N Engl J Med. 2020 NEJMoa2021436. doi: 10.1056/NEJMoa2021436.
9. Abani O, Abbas A, Abbas F, Abbas M, Abbasi S, Abbass H, et al. Casirivimab and imdevimab in patients admitted to hospital with COVID-19 (RECOVERY): a randomised, controlled, open-label, platform trial. The Lancet. 2022;399(10325):665-76.
10. Weinreich DM, Sivapalasingam S, Norton T, Ali S, Gao H, Bhore R, et al. REGN-COV2, a Neutralizing Antibody Cocktail, in Outpatients with Covid-19. New England Journal of Medicine. 2020;384(3):238-51.
11. Néant N, Lingas G, Le Hingrat Q, Ghosn J, Engelmann I, Lepiller Q, Gaymard A, Ferré V, Hartard C, Plantier JC, Thibault V, Marlet J, Montes B, Bouiller K, Lescure FX, Timsit JF, Faure E, Poissy J, Chidiac C, Raffi F, Kimmoun A, Etienne M, Richard JC, Tattevin P, Garot D, Le Moing V, Bachelet D, Tardivon C, Duval X, Yazdanpanah Y, Mentré F, Laouénan C, Visseaux B, Guedj J; French COVID Cohort Investigators and French Cohort Study groups. Modeling SARS-CoV-2 viral kinetics and association with mortality in hospitalized patients from the French COVID cohort. Proc Natl Acad Sci U S A. 2021 Feb 23;118(8):e2017962118. doi:10.1073/pnas.2017962118. PMID: 33536313.
12. Boschi C, Colson P, Bancod A, Moal V, La Scola B. Omicron Variant Escapes Therapeutic Monoclonal Antibodies (mAbs) Including Recently Released Evusheld®, Contrary to 8 Prior Main Variant of Concern (VOC). Clinical Infectious Diseases. 2022.
13. Zhou H, Tada T, Dcosta BM, Landau NR. Neutralization of SARS-CoV-2 Omicron BA. 2 by therapeutic monoclonal antibodies. Biorxiv. 2022.
14. Feld JJ, Kandel C, Biondi MJ, Kozak RA, Zahoor MA, Lemieux C, Borgia SM, Boggild AK, Powis J, McCready J, Tan DHS, Chan T, Coburn B, Kumar D, Humar A, Chan A, O'Neil B, Noureldin S, Booth J, Hong R, Smookler D, Aleyadeh W, Patel A, Barber B, Casey J, Hiebert R, Mistry H, Choong I, Hislop C, Santer DM, Lorne Tyrrell D, Glenn JS, Gehring AJ, Janssen HLA, Hansen BE. Peginterferon lambda for the treatment of outpatients with COVID-19: a phase 2, placebo-controlled randomised trial. Lancet Respir Med. 2021 Feb 5:S2213-2600(20)30566-X. doi: 10.1016/S2213-2600(20)30566-X. Epub ahead of print. PMID: 33556319.
15. Kissler SM, Fauver JR, Mack C, Tai C, Shiue KY, Kalinich CC, et al. Viral dynamics of SARS-CoV-2 infection and the predictive value of repeat testing. medRxiv. 2020:2020.10.21.20217042.
16. Chaccour C, Casellas A, Blanco-Di Matteo A, Pineda I, Fernandez-Montero A, Ruiz-Castillo P, et al. The effect of early treatment with ivermectin on viral load, symptoms and humoral response in patients with non-severe COVID-19: A pilot, double-blind, placebo-controlled, randomized clinical trial. EClinicalMedicine. 100720. doi: 10.1016/j.eclinm.2020.100720.
17. Pekosz A, Parvu V, Li M, Andrews JC, Manabe YC, Kodsi S, Gary DS, Roger-Dalbert C, Leitch J, Cooper CK. Antigen-Based Testing but Not Real-Time Polymerase Chain Reaction Correlates With Severe Acute Respiratory Syndrome Coronavirus 2 Viral Culture. Clin Infect Dis. 2021 Jan 20: ciaa1706. doi: 10.1093/cid/ciaa1706. Epub ahead of print. PMID: 33479756.
18. Kohmer N, Toptan T, Pallas C, Karaca O, Pfeiffer A, Westhaus S, Widera M, Berger A, Hoehl S, Kammel M, Ciesek S, Rabenau HF. The Comparative Clinical Performance of Four SARS-CoV-2 Rapid Antigen Tests and Their Correlation to Infectivity In Vitro. J Clin Med. 2021;10(2):E328. doi: 10.3390/jcm10020328. PMID: 33477365.
19. Rastawicki W, Gierczyński R, Juszczyk G, Mitura K, Henry BM. Evaluation of PCL rapid point of care antigen test for detection of SARS-CoV-2 in nasopharyngeal swabs. J Med Virol. 2021 Jan 6. doi: 10.1002/jmv.26765.
20. To KK, Tsang OT, Leung WS, Tam AR, Wu TC, Lung DC, et al. Temporal profiles of viral load in posterior oropharyngeal saliva samples and serum antibody responses during infection by SARS-CoV-2: an observational cohort study. Lancet Infect Dis. 2020;20(5):565-74.
21. Sri Santosh T, Parmar R, Anand H, Srikanth K, Saritha M. A Review of Salivary Diagnostics and Its Potential Implication in Detection of Covid-19. Cureus. 2020;12(4):e7708.
22. Williams E, Bond K, Zhang B, Putland M, Williamson DA. Saliva as a Noninvasive Specimen for Detection of SARS-CoV-2. J Clin Microbiol. 2020;58(8).
23. Thompson MG, Ferber JR, Odouli R, David D, Shifflett P, Meece JK, et al. Results of a pilot study using self-collected mid-turbinate nasal swabs for detection of influenza virus infection among pregnant women. Influenza Other Respir Viruses. 2015;9(3):155-60.
24. van Wesenbeeck L, Meeuws H, D'Haese D, Ispas G, Houspie L, Van Ranst M, et al. Sampling variability between two mid-turbinate swabs of the same patient has implications for influenza viral load monitoring. Virol J. 2014;11:233.
25. Corman VM, Landt O, Kaiser M, Molenkamp R, Meijer A, Chu DK, et al. Detection of 2019 novel coronavirus (2019-nCoV) by real-time RT-PCR. Euro Surveill. 2020;25(3).
26. Grein J, Ohmagari N, Shin D, Diaz G, Asperges E, Castagna A, et al. Compassionate Use of Remdesivir for Patients with Severe Covid-19. N Engl J Med. 2020;382(24):2327-36.
27. Piralla A, Daleno C, Pariani E, Conaldi P, Esposito S, Zanetti A, et al. Virtual quantification of influenza A virus load by real-time RT-PCR. J Clin Virol. 2013;56(1):65-8.
28. Kaulbach HC, White MV, Igarashi Y, Hahn BK, Kaliner MA. Estimation of nasal epithelial lining fluid using urea as a marker. J Allergy Clin Immunol. 1993;92(3):457-65.

**Appendices**

### 15. Appendix 1: Schedule of activities

| ***Procedures*** | | **D0** | | | **D1** | **D2** | **D3** | **D4** | **D5** | **D6** | **D7** | **D14** | **D28 (-2/ +7)** | **If new Tx** |
| --- | --- | --- | --- | --- | --- | --- | --- | --- | --- | --- | --- | --- | --- | --- |
|  |  | **Pre-1^st^ dose** | **H0** | **H1** |  |  |  |  |  |  |  |  |  |  |
| *Screening incl. Lateral Flow Ag test and urine pregnancy test* | | X |  |  |  |  |  |  |  |  |  |  |  |  |
| *Eligibility assessment* | | X |  |  |  |  |  |  |  |  |  |  |  |  |
| *Informed consent* | | X |  |  |  |  |  |  |  |  |  |  |  |  |
| *Randomisation* | | X |  |  |  |  |  |  |  |  |  |  |  |  |
| *Baseline information captured (inc symptoms, physical exam, vital signs, ECG and POC antibody test)* | | X |  |  |  |  |  |  |  |  |  |  |  |  |
| *Symptom checklist* | |  |  | X^1^ | X | X | X | X | X | X | X | X | X |  |
| *Observed dose* | |  | X^1^ |  | X^1^ | X^1^ | X^1^ | X^1^ | X^1^ | X^1^ |  |  |  |  |
| *Observation post dose* | |  |  | X^1^ |  |  |  |  |  |  |  |  |  |  |
| *Temperature** | |  |  | X | X | X | X | X | X | X | X | X |  |  |
| ***Virology*** | |  | | | | | | | | | | | | |
| *Oropharyngeal swabs*** | | X | X |  | X | X | X | X | X | X | X | X |  | X |
| ***Bloods*** | |  | | | | | | | | | | | | |
| *FBC+diff* | | X^2^ |  |  |  |  | X |  |  |  | X | X |  |  |
| *Biochemistry (including U&Es, LFTs LDH)* | | X^2^ |  |  |  |  | X |  |  |  | X | X |  |  |
| *Fibrinogen/*  *FDP* | | X |  |  |  |  | X |  |  |  | X | X |  |  |
| *Ferritin* | | X |  |  |  |  | X |  |  |  | X | X |  |  |
| *TNF/IL-6/IL-1* | | X |  |  |  |  | X |  |  |  | X | X |  |  |
| *Serology and stored sample* | | X |  |  |  |  | X |  |  |  | X | X |  |  |
| *Drug level* | | X |  |  |  |  | X |  |  |  | X | X |  |  |
| *Host genotyping* | | X |  |  |  |  |  |  |  |  |  |  |  |  |
|  | * Performed twice a day. For those taking drugs which are more than once per day, the patient will also record the time the dose was taken for any non-observed doses.  ** Taken in duplicate. Time swab done recorded. Some patients may be asked to give a saliva sample as well (see section 7.1)  X^1^Depends on dosing schedule of the drug (see appendix 2). The first dose will be given at the designated trial facility. For drugs where further doses are required, at least one dose per day will be observed by the study team. The time the drug is taken will be recorded by the study team.  X^2^ Results will be used to detect laboratory abnormalities included in the exclusion criteria (see appendix 4 exclusion criteria)  The final column, If new Tx, refers to if the treatment is changed while the patient is within the 1^st^ 7 days of the study.  **Note**: PK sampling schedules for determination of antiviral drug level are mentioned in Appendix 2. | | | | | | | | | | | | | |

### 16. Appendix 2: Study drugs

The maximum duration of treatment is 7 days so many of the toxicity concerns related to chronic dosing are not relevant to this short course treatment

#### 16.1. Hydroxychloroquine

Rationale

Although hydroxychloroquine and chloroquine have shown anti-SARS-CoV-2 activity *in vitro* (1-4), hydroxychloroquine in hospitalised patients did not result in clinical benefit in two large randomised controlled trials (5,6). Although this is strong evidence that it is ineffective in late stage disease, the evidence earlier in the disease process is equivocal including in early treatment. Meta-analysis of RCTs in pre-exposure prophylaxis is in the direction of benefit. There is therefore substantial uncertainty whether hydroxychloroquine has significant antiviral activity against SARS-CoV-2 in humans.

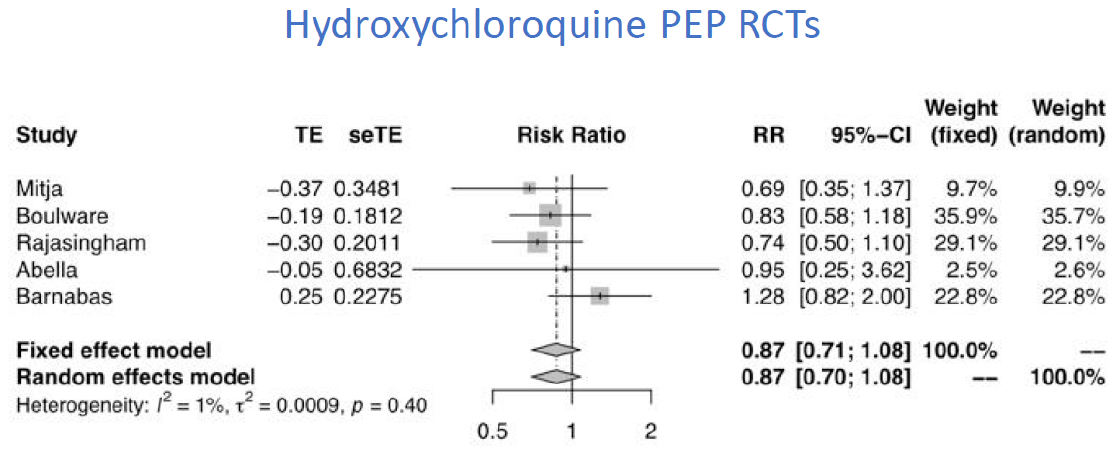

Composition and dose

The reference form of hydroxychloroquine is the Sanofi product, Plaquenil® (<https://www.medicines.org.uk/emc/product/1764/smpc>).

Hydroxychloroquine will either be obtained by the sponsor or the local teams from a reliable manufacturer. One tablet of hydroxychloroquine sulphate contains 200 mg of hydroxychloroquine sulphate which is equivalent to 155 mg hydroxychloroquine base. Hydroxychloroquine will be given for 7 days.

**The dosing schedule of hydroxychloroquine is:**

Day 0

- 2 tablets at time 0
- 2 tablets 12 hours later, then

Days 1 – 6

2 tablets per day

That is a total of 800 mg of hydroxychloroquine sulphate (salt) on the first day of treatment and 400 mg of hydroxychloroquine sulphate as a maintenance dose. The doses will be given with food to reduce gastric upset.

Pharmacokinetic characteristics

Hydroxychloroquine is generally well-absorbed, reaches a Cmax at a mean of ~2-3 hours, and following distribution and a multiexponential disposition profile, has a long terminal elimination half-life of ~54 days (5). The disposition of hydroxychloroquine in healthy volunteers is shown in Figure 3 (6).

Pharmacokinetic sampling schedule: (only if there is evidence of accelerated viral clearance)

All patients will need to be admitted for 12 hours for intense venous blood sampling around the **loading dose**.

The timings are:

- Pre-dose D0H0, then
- 1, 2, 3, 4, 6, 9, 12 and 24h post dose

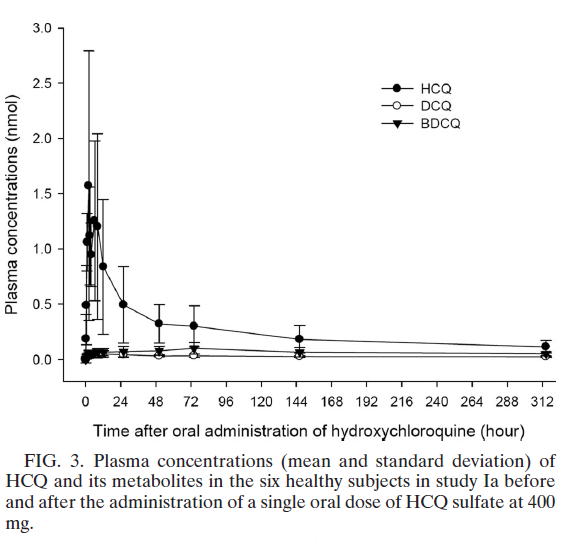
Note that blood will be taken on D3 and D7 for routine haematology and biochemistry. The EDTA sample will be used to measure the drug concentrations.

Toxicity

Hydroxychloroquine is well-tolerated and has been used for many years to treat patients with rheumatological diseases. The toxicity associated with long term use e.g. retinal disease, myopathy is not seen with short term use.

The most common side effects include dyspepsia, nausea, vomiting, headache and blurred vision; the latter is dose related and usually transient but patients should be warned about driving and operating machinery. Postural hypotension may occur in patients with a fever.

Cardiac toxicity

Hydroxychloroquine (and chloroquine) cause dose related QRS and QT prolongation but these are only clinically important in:

- Overdose (4)
- those with pre-existing QT prolongation
- patients with QT prolonging diseases e.g. myxoedema, ischaemic heart disease, hypokalaemia
- patients on QT prolonging drugs.

The large randomised controlled trials in COVID-19 (RECOVERY and SOLIDARITY) did not find an excess of arrhythmias in patients receiving high dose hydroxychloroquine.

More details regarding concomitant medication can be found at:

- <https://www.bnf.org/products/bnf-online/> - available in the UK
- <https://www.uptodate.com/drug-interactions/?source=responsive_home#di-druglist> – this requires a subscription
- <https://compendium.ch/fr/Patient> - A Swiss website which deals with drug interactions
- <https://crediblemeds.org/>

Skin

These include:

exacerbation of psoriasis (case reports)

acute generalised exanthematous pustulosis (AGEP) that may be associated with fever and neutrophilia and resolves with discontinuation of the offending drug

itching / prickly sensation that is more common in dark skinned individuals

Contraindications

In our study, hydroxychloroquine is contraindicated in:

those with known hypersensitivity to hydroxychloroquine or other 4-aminoquinolines (e.g. amodiaquine)

patients with pre-existing retinopathy of the eye

severe liver disease, severe renal disease, epilepsy, porphyria and myasthenia gravis are relative contraindications but such patients would not be enrolled in this trial

Cautions

Long term use of hydroxychloroquine has caused hypoglycaemia in type II diabetes. Diabetic individuals will not be recruited.

Patients with a history of unexplained syncope and/ or family history of sudden unexplained cardiac death, should have an ECG is performed and should be enrolled only if the Bazett QTc <430 ms in males and <450 ms in females.

Relevant drug interactions

Antacids may reduce absorption of hydroxychloroquine so it is advised that a 4-hour interval be observed between hydroxychloroquine sulphate and antacid dosing

References

1. Liu J, Cao R, Xu M, Wang X, Zhang H, Hu H, et al. Hydroxychloroquine, a less toxic derivative of chloroquine, is effective in inhibiting SARS-CoV-2 infection in vitro. Cell Discov. 2020;6:16.
2. Yao X, Ye F, Zhang M, Cui C, Huang B, Niu P, et al. In Vitro Antiviral Activity and Projection of Optimized Dosing Design of Hydroxychloroquine for the Treatment of Severe Acute Respiratory Syndrome Coronavirus 2 (SARS-CoV-2). Clin Infect Dis. 2020.
3. Wang M, Cao R, Zhang L, Yang X, Liu J, Xu M, et al. Remdesivir and chloroquine effectively inhibit the recently emerged novel coronavirus (2019-nCoV) in vitro. Cell Res. 2020;30(3):269-71.
4. White NJ, Watson JA, Hoglund RM, Chan XHS, Cheah PY, Tarning J. COVID-19 prevention and treatment: A critical analysis of chloroquine andhydroxychloroquine clinical pharmacology. PLoS Med 2020; 17: e1003252.
5. RECOVERY group, Horby P, Mafham M, Linsell L, Bell JL, Staplin N, et al. Effect of Hydroxychloroquine in Hospitalized Patients with Covid-19. N Engl J Med. 2020;383(21):2030-40.
6. Consortium WHO Solidarity Trial, Pan H, Peto R, Henao-Restrepo AM, Preziosi MP, Sathiyamoorthy V, et al. Repurposed Antiviral Drugs for Covid-19 - Interim WHO Solidarity Trial Results. N Engl J Med. 2020; 384(6):497-511.

#### 1.1. Remdesivir

Rationale

Remdesivir is a nucleoside analogue prodrug that shows *in vitro* inhibition of the viral RNA polymerase of SARS-CoV (1) and the Middle East Respiratory Syndrome (MERS-CoV) (2) and SARS-CoV-2 (1). In clinical trials, remdesivir has been associated with earlier clinical improvement,hospital discharge and reduced mortality (2-7). A retrospective study showed that administration of remdesivir in hospitalized patients significantly reduced viral load (8).

Composition and dose

Remdesivir will be either be obtained by the sponsor or the local teams. Much of the following is taken from the Gilead summary of product characteristics (SPC) (<https://www.medicines.org.uk/emc/product/11597/smpc/print>).

Each vial contains remdesivir 100 mg of remdesivir concentrate for solution for infusion. After reconstitution, each vial contains 5 mg/mL of remdesivir solution. The recommended dosage of remdesivir in patients 12 years of age and older and weighing at least 40 kg is:

- Day 1 – single loading dose of remdesivir 200 mg given by intravenous infusion
- Day 2 onwards – 100 mg given once daily by intravenous infusion

The SPC says “The total duration of treatment should be at least 5 days and not more than 10 days.” In our trial we will give remdesivir **for 5 days**.

Remdesivir is given by intravenous infusion in 0.9% saline; there are several options.

| Infusion Bag Volume | Infusion Time | Rate of Infusion |
| --- | --- | --- |
| 250 mL | 30 min | 8.33 mL/min |
|  | 60 min | 4.17 mL/min |
|  | 120 min | 2.08 mL/min |
| 100 mL | 30 min | 3.33 mL/min |
|  | 60 min | 1.67 mL/min |
|  | 120 min | 0.83 mL/min |

In our trial, we will give the:

- loading dose in 210 mL of N saline over 60 minutes
- maintenance doses in 230 mL of N saline

Care should be taken to prevent contamination when preparing the remdesivir dose. Use **aseptic technique** and administer IV medicines immediately after preparation.

Recommended dilution instructions – Reconstituted remdesivir powder for concentrate for solution for infusion.

| Remdesivir dose | Sodium chloride 9 mg/mL (0.9%) infusion bag volume to be used | Volume to be withdrawn and discarded from sodium chloride 9 mg/mL (0.9%) infusion bag | Required volume of reconstituted remdesivir |
| --- | --- | --- | --- |
| 200 mg  (2 vials) | 250 mL | 40 mL | 2 × 20 mL |
| 100 mg  (1 vial) | 250 mL | 20 mL | 20 mL |

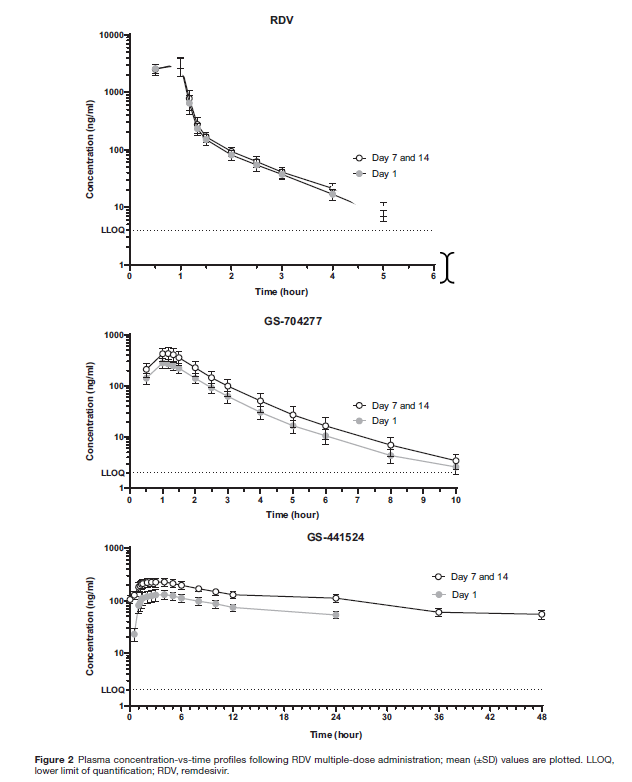
Pharmacokinetic properties

The pharmacokinetic profile for remdesivir (RDV) and two of its metabolites are shown below (5). Peak plasma concentrations of remdesivir occur at the end of the infusion, declining rapidly thereafter with a half-life of approximately 1 hour. Peak plasma concentrations of the principal biologically active metabolite GS-441524 were observed at 1.5 to 2.0 hours post start of a 30 minute infusion. GS-441524 has a median elimination half-life of 27h.

Remdesivir undergoes extensive intracellular metabolism by esterases to produce several active antiviral metabolites.

Renal clearance is the major elimination pathway for GS-441524 whilst only 10% of RDV is found in urine. With its long half-life, GS-441524 accumulates ~1.9-fold after daily dosing (5).

Pharmacokinetic sampling schedule: (only if there is evidence of accelerated viral clearance)

All patients will need to be admitted for 12 hours for intense venous blood sampling around the **loading dose**.

The timings are:

- Pre-dose D0H0, then
- 1, 2, 3, 4, 6, 9, 12 and 24h post dose

Note that blood will be taken on D3 and D7 for routine haematology and biochemistry. The EDTA sample will be used to measure the drug concentrations.

Toxicity

Remdesivir is generally well-tolerated. The main toxicities are listed below.

Hypersensitivity including infusion-related and anaphylactic reactions

Signs and symptoms may include hypotension, hypertension, tachycardia, bradycardia, hypoxia, fever, dyspnoea, wheezing, angioedema, rash, nausea, vomiting, diaphoresis, and shivering.

Stop the infusion and treat as clinically indicated.

A hypersensitivity reaction contraindicates additional doses.

Transaminase elevations

Transaminase elevations have been observed in healthy volunteers and patients with COVID-19 (9).

The SPC says remdesivir should be discontinued in patients who develop:

- ALT ≥ 5 times the upper limit of normal during treatment with remdesivir
- ALT elevation accompanied by signs or symptoms of liver inflammation or increasing conjugated bilirubin, alkaline phosphatase, or international normalised ratio (INR)

In our study, remdesivir will be stopped if:

- ALT ≥ 5 x ULN
- ALT > 3 x ULN & total bilirubin > 2 x ULN (meeting the Hy’s Law criteria) (9)
- Total bilirubin > 2 x ULN (suggesting a cholestatic liver injury)

Common symptoms & signs

These include:

- Headache
- Nausea
- Rashes e.g. maculopapular rash

Contraindications

Known hypersensitivity.

Renal impairment

Patients with eGFR ≥ 30 mL/min/1.73m^2^ have received remdesivir for treatment of COVID-19 with no dose adjustment. Remdesivir should not be used in patients with eGFR <30 mL/min/1.73m^2^. This is because remdesivir’s main metabolite GS-441524 and the excipient, betadex sulfobutyl ether sodium, are renally cleared and accumulate in patients with decreased renal function which may also be related to a reduced volume of distribution (10).

Hepatic impairment

Although the pharmacokinetic characteristics of remdesivir have not been evaluated in liver disease, it should not be used in patients with impaired liver function given its potential liver toxicity.

Relevant drug interactions

No clinical interaction studies have been performed with remdesivir.

*In vitro*, remdesivir is a substrate for esterases in plasma and tissue, drug metabolising enzymes CYP2C8, CYP2D6, and CYP3A4, and is a substrate for Organic Anion Transporting Polypeptides 1B1 (OATP1B1) and P-glycoprotein (P-gp) transporters but there are no studies of *in vitro* drug interactions.

References

1. Brown AJ, Won JJ, Graham RL, Dinnon KH, 3rd, Sims AC, Feng JY, et al. Broad spectrum antiviral remdesivir inhibits human endemic and zoonotic deltacoronaviruses with a highly divergent RNA dependent RNA polymerase. Antiviral Res. 2019;169:104541.
2. Sheahan TP, Sims AC, Leist SR, Schafer A, Won J, Brown AJ, et al. Comparative therapeutic efficacy of remdesivir and combination lopinavir, ritonavir, and interferon beta against MERS-CoV. Nat Commun. 2020;11(1):222.
3. Grein J, Ohmagari N, Shin D, Diaz G, Asperges E, Castagna A, et al. Compassionate Use of Remdesivir for Patients with Severe Covid-19. N Engl J Med. 2020;382(24):2327-36.
4. Beigel JH, Tomashek KM, Dodd LE, Mehta AK, Zingman BS, Kalil AC, et al. Remdesivir for the Treatment of Covid-19 - Final Report. N Engl J Med. 2020;383(19):1813-26.
5. Humeniuk R, Mathias A, Cao H, Osinusi A, Shen G, Chng E, et al. Safety, Tolerability, and Pharmacokinetics of Remdesivir, An Antiviral for Treatment of COVID-19, in Healthy Subjects. Clin Transl Sci. 2020;13(5):896-906.
6. Diaz G, Christensen A, Pusch T, Goulet D, Chang S, Grunkemeier G et al. Remdesivir and Mortality in Patients With Coronavirus Disease 2019. Clinical Infectious Diseases. 2021;.
7. Gottlieb R, Vaca C, Paredes R, Mera J, Webb B, Perez G et al. Early Remdesivir to Prevent Progression to Severe Covid-19 in Outpatients. New England Journal of Medicine. 2022;386(4):305-315.
8. Biancofiore, A., Mirijello, A., Puteo, M., Di Viesti, M., Labonia, M., Copetti, M., De Cosmo, S. and Lombardi, R., 2022. Remdesivir significantly reduces SARS‐CoV‐2 viral load on nasopharyngeal swabs in hospitalized patients with COVID‐19: A retrospective case–control study. Journal of Medical Virology, pp.1-6.
9. Navarro VJ, Senior JR. Drug-related hepatotoxicity. N Engl J Med. 2006;354(7):731-9.
10. Sorgel F, Malin JJ, Hagmann H, Kinzig M, Bilal M, Eichenauer DA, et al. Pharmacokinetics of remdesivir in a COVID-19 patient with end-stage renal disease on intermittent haemodialysis. J Antimicrob Chemother. 2021; 76(3):825-827.

#### 16.2. Lopinavir/ritonavir

Rationale

Lopinavir is a human immunodeficiency virus 1 (HIV-1) protease inhibitor, which is combined with ritonavir to increase exposure to lopinavir. It is licensed in adults and children. It has been widely used including in pregnant women. Lopinavir has *in vitro* inhibitory activity against SARS coronavirus (SARS-CoV) and MERS-CoV (1). In common marmosets infected with MERS-CoV, animals treated with lopinavir/ritonavir had improved clinical, radiological, and pathological outcomes and reduced viral loads compared with untreated animals. The RECOVERY trial showed no beneficial effect of lopinavir/ritonavir in patients hospitalised with COVID-19, consistent with an earlier study in China, and closed randomisation to that treatment arm (2). However, the main therapeutic window for efficacy of an antiviral drug is earlier in the disease, and there remains uncertainty as to whether an antiviral effect for this drug exists.

Composition & dose

Kaletra® contains 200 mg of liponavir and 50 mg of ritonavir.

Suitable tablets will be obtained by the sponsor or the local teams from a reliable manufacturer.

The dosing schedule is: **2 tablets bd for 7 days**.

Dosing and Pharmacokinetic characteristics

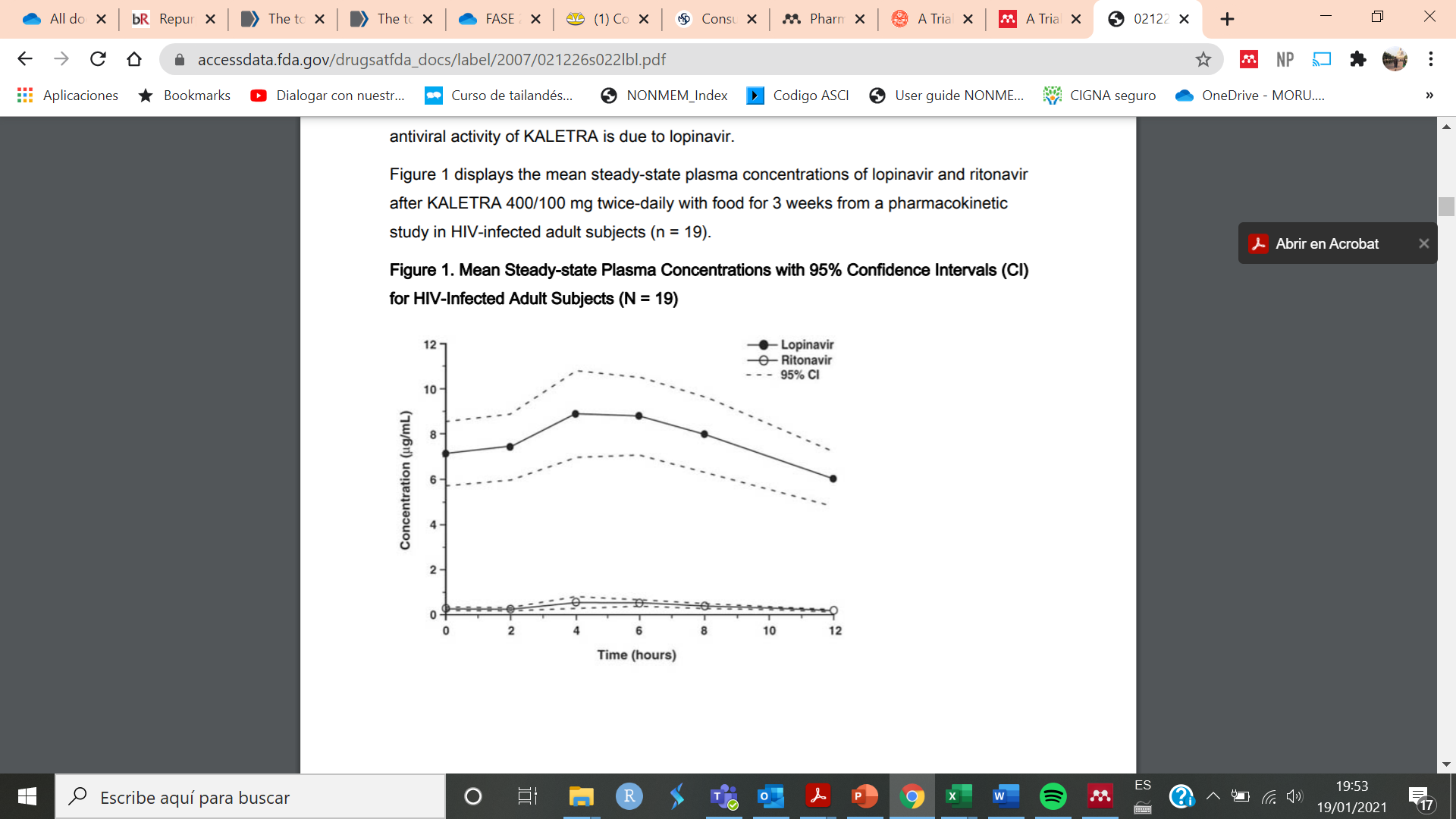
The Pharmacokinetic properties of lopinavir/ritonavir have been evaluated in healthy adult volunteers and in HIV-infected patients; no substantial differences were observed between the two groups. Lopinavir is essentially completely metabolised by CYP3A. Ritonavir inhibits the metabolism of lopinavir, thereby increasing the plasma levels of lopinavir. Administration of lopinavir/ritonavir 400/100 mg twice-daily yields mean steady-state lopinavir plasma concentrations 15- to 20-fold higher than those of ritonavir in HIV-infected patients. The plasma levels of ritonavir are less than 7% of those obtained after the ritonavir dose of 600 mg twice-daily. Figure 1 displays the mean steady-state plasma concentrations of lopinavir and ritonavir after lopinavir/ritonavir 400/100 mg twice-daily with food for 3 weeks from a Pharmacokinetic study in HIV-infected adult subjects (n = 19). This is also the most commonly used and studied lopinavir/ritonavir dosing regimen for COVID-19 treatment (2).

**Figure 1. Mean Steady-state Plasma Concentrations with 95% Confidence Intervals (CI) for HIV-Infected Adult Subjects (N = 19)**

In a pharmacokinetic study in HIV-positive subjects (n = 19), multiple dosing with 400/100 mg KALETRA twice-daily with food for 3 weeks produced a mean ± SD lopinavir peak plasma concentration (Cmax) of 9.8 ± 3.7 µg/mL, occurring approximately 4 hours after administration. The mean steady-state trough concentration prior to the morning dose was 7.1 ± 2.9 µg/mL and minimum concentration within a dosing interval was 5.5 ± 2.7 µg/mL. Lopinavir AUC over a 12-hour dosing interval averaged 92.6 ± 36.7 µg• h/mL.

Lopinavir is extensively metabolised by the hepatic cytochrome P450 system, almost exclusively by the CYP3A isozyme. Ritonavir is a potent CYP3A inhibitor which inhibits the metabolism of lopinavir, and thereby increases its plasma levels (exposure). Ritonavir has also been shown to induce metabolic enzymes, resulting in the induction of its own metabolism. Pre-dose lopinavir concentrations decline with time during multiple dosing, stabilising after approximately 10 to 16 days.

Pharmacokinetic sampling schedule: (only if there is evidence of accelerated viral clearance)

All patients will need to be admitted for 12 hours for intense sampling.

The timings are:

- Pre-dose D0H0, then
- 1, 2, 3, 4, 6, 9, 12 and 24h post dose.

Note that blood will be taken on D3 and D7 for routine haematology and biochemistry. The EDTA sample will be used to measure the drug concentrations.

Toxicity

Adverse effects of lopinavir/ritonavir include gastrointestinal distress such as nausea and diarrhoea (up to 28%) and hepatotoxicity (2%-10%). In patients with COVID-19, these adverse effects may be exacerbated by viral infection because approximately 20% to 30% of patients have elevated transaminases at presentation with COVID-19.

The following adverse reactions, commonly related to the chronic use of the drug, are discussed in greater detail below:

- Lipid elevations
- Hyperglycaemia
- Pancreatitis
- Hepatotoxicity
- QT Interval Prolongation
- PR Interval Prolongation

Lipid elevations

Treatment with lopinavir/ritonavir has resulted in large increases in the concentration of total cholesterol and triglycerides.

Hyperglycaemia

New onset diabetes mellitus, exacerbation of pre-existing diabetes mellitus, and hyperglycaemia have been reported during post-marketing surveillance in HIV-1 infected patients receiving protease inhibitor therapy. Some patients required either initiation or dose adjustments of insulin or oral hypoglycaemic agents for treatment of these events. In some cases, diabetic ketoacidosis has occurred. In those patients who discontinued protease inhibitor therapy, hyperglycaemia persisted in some cases.

Pancreatitis

Pancreatitis should be considered if clinical symptoms (nausea, vomiting, abdominal pain) or abnormalities in laboratory values (such as increased serum lipase or amylase values) suggestive of pancreatitis occur. Although a causal relationship to lopinavir/ritonavir has not been established, marked triglyceride elevations are a risk factor for development of pancreatitis.

Hepatotoxicity

There have been post marketing reports of hepatic dysfunction, including some fatalities. These have generally occurred in patients with advanced HIV-1 disease taking multiple concomitant medications in the setting of underlying chronic hepatitis or cirrhosis. A causal relationship with lopinavir/ritonavir therapy has not been established.

Elevated transaminases with or without elevated bilirubin levels have been reported in HIV-1 mono-infected and uninfected patients as early as 7 days after the initiation of lopinavir/ritonavir in conjunction with other antiretroviral agents. In some cases, the hepatic dysfunction was serious; however, a definitive causal relationship with lopinavir/ritonavir therapy has not been established. Appropriate laboratory testing should be conducted prior to initiating therapy with lopinavir/ritonavir and patients should be monitored closely during treatment.

QT prolongation

Post marketing cases of QT interval prolongation and Torsade de Pointes have been reported although causality of lopinavir/ritonavir could not be established. Avoid use in patients with congenital long QT syndrome, those with hypokalaemia, and with other drugs that prolong the QT interval.

PR interval prolongation

Lopinavir/ritonavir prolongs the PR interval in some patients. Cases of second- or third-degree atrioventricular block have been reported. Hence, these drugs should be used with caution in patients with underlying structural heart disease, pre-existing conduction system abnormalities, ischemic heart disease or cardiomyopathies, as these patients may be at increased risk for developing cardiac conduction abnormalities. The impact on the PR interval of co-administration of lopinavir/ritonavir with other drugs that prolong the PR interval (including calcium channel blockers, beta-adrenergic blockers, digoxin and atazanavir) has not been evaluated. As a result, co-administration of lopinavir/ritonavir with these drugs should be undertaken with caution, particularly with those drugs metabolized by CYP3A. Clinical monitoring is recommended.

Contraindications

In our study, lopinavir/ritonavir is contraindicated in:

hypersensitivity to lopinavir/ritonavir or any of its ingredients, including ritonavir.

co-administration with drugs highly dependent on CYP4503A.

co-administration with potent CYP450 3A inducers

More details regarding concomitant medication can be found at:

<https://www.covid19-druginteractions.org/checker>

<https://crediblemeds.org/>

<https://compendium.ch/fr/Patient> - this is a Swiss website dealing with drug interactions

<https://www.bnf.org/products/bnf-online/> - available only in the UK

References

1. Chu CM, Cheng VC, Hung IF, Wong MM, Chan KH, Chan KS, et al. Role of lopinavir/ritonavir in the treatment of SARS: initial virological and clinical findings. Thorax. 2004;59(3):252-6.
2. Cao B, Wang Y, Wen D, Liu W, Wang J, Fan G, et al. A Trial of Lopinavir-Ritonavir in Adults Hospitalized with Severe Covid-19. N Engl J Med. 2020;382(19):1787-99.

#### 16.3. Miglustat

Rationale

Miglustat is a drug that is in current clinical use for lysosomal storage disorders and has shown to be active against a variety of viral infections. A key mechanism by which iminosugars act as antivirals is their ability to disrupt glycoprotein folding, arising from the inhibition of ER α-glu I and II (1). Miglustat has been shown to inhibit SARS-CoV-2 in different cell types at concentrations compatible with those obtained for the treatment of Gaucher’s disease and Niemann-Pick type C patients (2), although more recent evidence in the calu-3 cell line suggests that higher doses (those used in the treatment of HIV may be required to reach *in vivo* antiviral drug concentrations) (unpublished communication). Time of addition studies indicate that the inhibitory activity is at the post-entry level and affects release of infectious virus. The proper folding and release of the Spike protein, and progeny virus appear mostly affected.

Composition & dose

Zavesca® capsules contain 100 mg of miglustat.

Suitable tablets will be obtained by the sponsor or the local teams from the manufacturer.

The approved doses are:

- 100 mg TDS and 200 mg TDS are approved doses (3) but doses of up to 3000mg/ day have been used in HIV patients for up to 6 months.

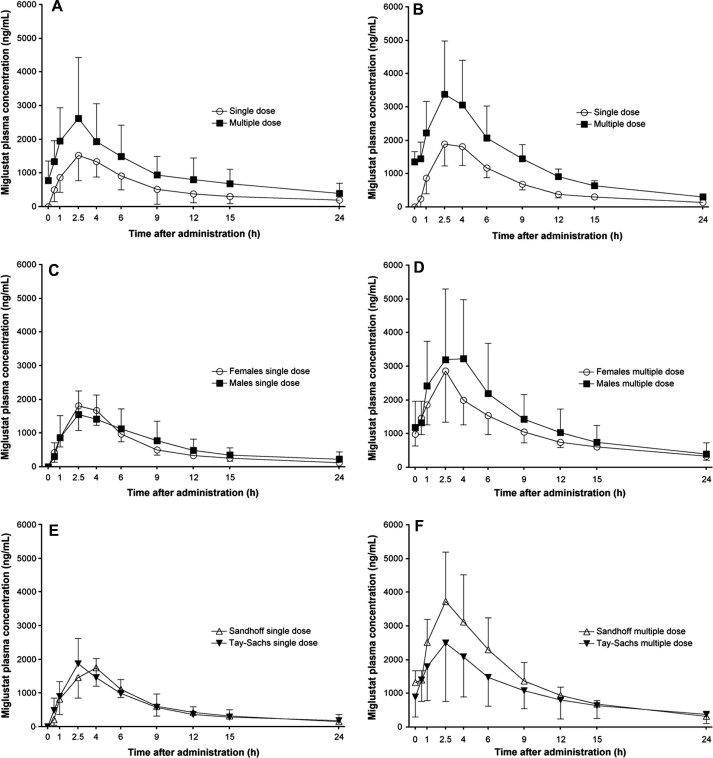
In our study the dose will be using the highest dose of **1g TDS PO for 7 days**

Pharmacokinetic characteristics

In a study of 28 adult patients with Gaucher’s disease receiving 100mg three times daily the time of maximum plasma concentration and half-life were 2.5 and 6.3 h, respectively, with a maximum concentration of 0.86 μg/mL. Steady-state plasma concentrations were achieved after 4–6 weeks of treatment (3).

Miglustat plasma exposure reaches steady state at about 72 h and the ratio trough/peak is overall between 0.6 and 0.7.

Pharmacokinetic sampling schedule: (only if there is evidence of accelerated viral clearance)

All patients will need to be admitted for 12 hours for intense sampling.

The timings are:

- Pre-dose D0H0, then
- 1, 2, 3, 4, 6, 9, 12 and 24h

Note that blood will be taken on D3 and D7 for routine haematology and biochemistry. The EDTA sample will be used to measure the drug concentrations.

Toxicity

This drug is commonly used in chronic conditions, and the toxicities described reflect that situation. It was also evaluated at doses of up to 3000 mg/day (the dose being evaluated in this study for 7 days) for up to six months in Human Immunodeficiency Virus (HIV)-positive patients. In these studies, the adverse events included granulocytopenia, dizziness, and paraesthesia. Leukopenia and neutropenia have also been observed in a similar group of patients receiving 800 mg/day or above.

- Gastrointestinal disturbances

Gastrointestinal events, mainly osmotic diarrhoea, have been observed in more than 80% of patients, either at the outset of treatment or intermittently during it. The mechanism is most likely inhibition of intestinal disaccharidases in the gastrointestinal tract. In clinical practice, miglustat-induced gastrointestinal events have been observed to respond to individualised diet modification, to taking it between meals, and/or to use anti-diarrhoeal medicinal products such as loperamide. In some patients, temporary dose reduction may be necessary (4).

- Fertility

After a single study in rodents suggested that miglustat could interfere with spermatogenesis there were concerns over reproductive toxicity. However these concerns have been allayed by studies in humans which have not shown effects on spermatogenesis (5). Contraceptive measures should be used by women of childbearing potential as miglustat crosses the placenta and should not be used during pregnancy. Studies in animals have shown reproductive toxicity, including dystocia. The potential risk for humans is unknown. It should also not be taken during breastfeeding.

- Renal impairment

Due to limited experience, miglustat should be used with caution in patients with renal or hepatic impairment. This drug is not recommended in patients with severe renal impairment (creatinine clearance < 30 ml/min/1.73m^2^).

Contraindications

Known hypersensitivity.

Cautions

Female patients should use contraception while receiving treatment and continue for one week after finishing. There are no relevant drug interactions

References

1. Alonzi DS, Scott KA, Dwek RA, Zitzmann N. Iminosugar antivirals: the therapeutic sweet spot. Biochem Soc Trans. 2017;45(2):571-82.
2. Williams SJ, Goddard-Borger ED. Alpha-glucosidase inhibitors as host-directed antiviral agents with potential for the treatment of COVID-19. Biochem Soc Trans. 2020;48(3):1287-95.
3. Maegawa GH, van Giersbergen PL, Yang S, Banwell B, Morgan CP, Dingemanse J, et al. Pharmacokinetics, safety and tolerability of miglustat in the treatment of pediatric patients with GM2 gangliosidosis. Mol Genet Metab. 2009;97(4):284-91.
4. Kuter DJ, Mehta A, Hollak CE, Giraldo P, Hughes D, Belmatoug N, et al. Miglustat therapy in type 1 Gaucher disease: clinical and safety outcomes in a multicenter retrospective cohort study. Blood Cells Mol Dis. 2013;51(2):116-24.
5. Amory JK, Muller CH, Page ST, Leifke E, Pagel ER, Bhandari A, Subramanyam B, Bone W, Radlmaier A, Bremner WJ. Miglustat has no apparent effect on spermatogenesis in normal men. Hum Reprod. 2007 Mar;22(3):702-7. doi: 10.1093/humrep/del414. Epub 2006 Oct 25. PMID: 17067996.

#### 1.2. Ivermectin

Rationale

Ivermectin is a macrocyclic lactone and broad-spectrum antiparasitic, antibacterial, and antiviral activity. It has an excellent safety profile and with 3.5 billion treatments distributed by mass drug administration for onchocerciasis and lymphatic filariasis elimination in Africa and Latin America since 1987 (Mectizan Donation Program). The side effects of ivermectin largely relate to the body’s reaction to the death of parasites (Mazzotti-type reaction) but is otherwise generally well-tolerated (1,2). Ivermectin has demonstrated broad-spectrum *in vitro* antiviral activity and is being used on a number of continents in the management of COVID-19. There are also reports from trials suggesting benefit and given the low cost, safety, tolerability and availability of this drug, it would be an attractive candidate in the prevention and treatment of COVID-19, if shown to have benefit in this illness.

Ivermectin has *in vitro* activity against the novel coronavirus SARS-CoV-2, which causes coronavirus disease (COVID-19). Ivermectin also inhibits influenza, HIV, Dengue, West Nile Virus, Yellow Fever virus, Japanese encephalitis, tick-borne encephalitis virus and Venezuelan equine encephalitis virus *in vitro* (2,3). The EC_50_ (the concentration to decrease the cytopathic effect of the virus by 50%) for SARS-CoV-2 on modified African green monkey kidney (Vero/hSLAM) cells was reportedly ~2µM and a 99.8% reduction in viral RNA at 48hrs in samples treated with 5µM concentrations (2), although the relevance of this *in vitro* cell line to human disease, like with other *in vitro* models, remains uncertain.

During the COVID-19 pandemic, due to early pre-print publications of the benefit of Ivermectin, this drug has been used widely at different stages of the disease, at different doses, for different durations and outside of clinical trials. RCTs of questionable quality seem to suggest improvements in virologic and clinical parameters, some demonstrating very large benefits.

Composition & dose

One tablet of ivermectin as STROMECTOL® contains 3 mg of ivermectin (<https://mri.cts-mrp.eu/Human/Downloads/NL_H_3952_001_FinalSPC.pdf>) where 3mg tablets are not available a corresponding dosing table will be created based on the dosing of 600mcg/kg bodyweight.

Ivermectin will either be obtained by the sponsor or the local teams from a reliable manufacturer.

The dose in this study is **600 micrograms/kg body weight given once daily for 7 days** (taken with food).

| **Weight in kg** | **Number of 6 mg tablets** | **Dose in mg** | **mg/kg** |
| --- | --- | --- | --- |
| 40 - <50 | 4 | 24 | 0.49 - 0.6 |
| 50 - <60 | 5 | 30 | 0.51 - 0.6 |
| 60 - <70 | 6 | 36 | 0.52 - 0.6 |
| 70 - <80 | 7 | 42 | 0.53 - 0.6 |
| 80 - <90 | 8 | 48 | 0.54 - 0.6 |
| 90 - <100 | 9 | 54 | 0.55 - 0.6 |
| ≥ 100 | 10 | 60 | ≤ 0.6 |

Pharmacokinetic characteristics

Ivermectin is incompletely absorbed (~50% bioavailable relative to an oral hydroalcoholic solution) following oral doses of ivermectin tablets, with a Tmax of ~4 hours.

Ivermectin has a molar mass of 875.1g/mmol, and as such 2µM (concentration at which anti-SARS-CoV-2 activity has been demonstrated) corresponds to a concentration of 1750 µg/L. Following oral administration of ivermectin with a standard oral dose of 12 mg (mean <200 µg/kg) of oral Ivermectin producing a C_max_ of 23.5-50µg/L (4,5). At this dose the C_max_ in plasma is some 35 times lower than the EC_50_ of the virus and an appreciable *in vivo* effect would not be expected to occur. However, in other experiments, *in vitro* effects of ivermectin do not adequately account for the *in vivo* effects observed. Baby hamster kidney (BHK-21) cells infected with pseudorabies virus exhibited an ivermectin EC_50_ of ~2µM at 72hours, which is similar to that of SARS-CoV-2, and would lead one to expect that a therapeutic response would not be possible. However, published in the same paper, single dose ivermectin (200 µg/kg) administered concomitantly or 12 hours after pseudorabies virus inoculation was able to suppress viral DNA synthesis and save 60% of infected mice. A 200 µg/kg ivermectin dose in mice has a human equivalent dosing (HED) of 16.3 µg/kg. The reason behind this *in vitro* to *in vivo* disparity is unclear but may relate to a more active ivermectin metabolite as is seen with this drug in other indications, as was suggested by a paper showing a 20- to 35-fold increase in mortality of mosquitoes which fed on blood from single dose ivermectin (400 µg/kg) treated volunteers compared to *in vitro* ivermectin spiked blood (6).

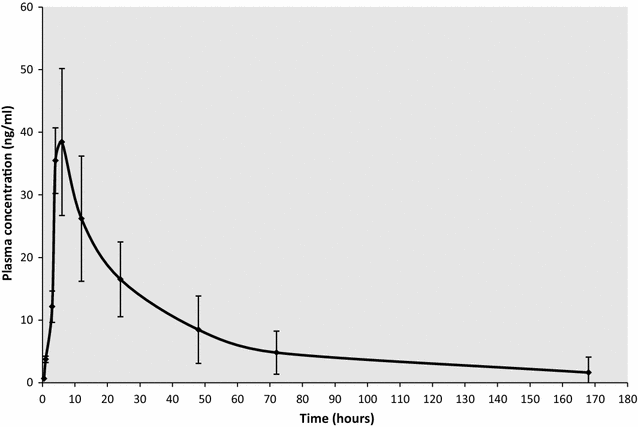
In humans, a phase III randomised, placebo-controlled, double blind trial showed a statistically significant difference in Dengue NS1 clearance times between ivermectin (400 or 600 µg/kg for three days) and placebo, although this did not translate into clinical efficacy (7). Again, this antiviral effect was seen despite *in vitro* EC_50_s against dengue which would suggest adequate *in vivo* levels would not be achieved (an order of magnitude lower).

Administration of 30 mg (333 to 600 μg/kg) ivermectin following a high-fat meal resulted in an approximate 2.5-fold increase in bioavailability compared to fasted healthy volunteers.

Ivermectin is metabolised in humans, and ivermectin and/or its metabolites are excreted almost exclusively in the faeces over an estimated 12 days with less than 1% of the administered dose being excreted in the urine. The plasma half-life of ivermectin in man is about 12 hours (9.8-14.3 h) and that of the metabolites is about 3 days.

Pharmacokinetic sampling schedule: (only if there is evidence of accelerated viral clearance)

All patients will need to be admitted to hospital for 12 hours for intense sampling. The timings are:

- Pre-dose D0H0, then
- 1, 2, 3, 4, 6, 9, 12 and 24h

Note that blood will be taken on D3 and D7 for routine haematology and biochemistry. The EDTA sample will be used to measure the drug concentrations.

Toxicity

Risks related to ivermectin are very low. The side effects of ivermectin largely relate to the body’s reaction to the death of parasites (Mazzotti-type reaction) with Onchocerciasis but is otherwise generally well-tolerated, and would not be an issue if studied in a non-endemic region.

Contraindications

Hypersensitivity reaction to ivermectin

Cautions

Ivermectin has not been studied in patients with impaired hepatic function or impaired renal function.

Relevant Drug interactions

None

References

1. Zoure HG, Wanji S, Noma M, Amazigo UV, Diggle PJ, Tekle AH, et al. The geographic distribution of Loa loa in Africa: results of large-scale implementation of the Rapid Assessment Procedure for Loiasis (RAPLOA). PLoS Negl Trop Dis. 2011;5(6):e1210.
2. Crump A, Omura S. Ivermectin, 'wonder drug' from Japan: the human use perspective. Proc Jpn Acad Ser B Phys Biol Sci. 2011;87(2):13-28.
3. Mastrangelo E, Pezzullo M, De Burghgraeve T, Kaptein S, Pastorino B, Dallmeier K, et al. Ivermectin is a potent inhibitor of flavivirus replication specifically targeting NS3 helicase activity: new prospects for an old drug. J Antimicrob Chemother. 2012;67(8):1884-94.
4. Caly L, Druce JD, Catton MG, Jans DA, Wagstaff KM. The FDA-approved Drug Ivermectin inhibits the replication of SARS-CoV-2 in vitro. Antiviral Res. 2020:104787.
5. Gonzalez Canga A, Sahagun Prieto AM, Diez Liebana MJ, Fernandez Martinez N, Sierra Vega M, Garcia Vieitez JJ. The pharmacokinetics and interactions of ivermectin in humans--a mini-review. AAPS J. 2008;10(1):42-6.
6. Kobylinski KC, Jittamala P, Hanboonkunupakarn B, Pukrittayakamee S, Pantuwatana K, Phasomkusolsil S, et al. Safety, Pharmacokinetics, and Mosquito-Lethal Effects of Ivermectin in Combination With Dihydroartemisinin-Piperaquine and Primaquine in Healthy Adult Thai Subjects. Clin Pharmacol Ther. 2019; 107(5):1221-1230.
7. Yamasmith, E., et al., Efficacy and Safety of Ivermectin against Dengue Infection: A Phase III, Randomized, Double-blind, Placebo-controlled Trial, in the 34th Annual Meeting The Royal College of Physicians of Thailand- ‘Internal Medicine and One Health'. 2018: Chonburi, Thailand.

#### 1.3. Nitazoxanide

Rationale

Nitazoxanide is a broad-spectrum antiprotozoal and antiviral drug first discovered in the 1980s. It has an excellent safety profile, the only contraindication to nitazoxanide being a hypersensitivity reaction to nitazoxanide. Nitazoxanide has demonstrated broad-spectrum *in vitro* antiviral activity including against SARS-CoV-2. There are also reports from trials suggesting benefit and given the low cost, safety, tolerability and availability of this drug, it would be an attractive candidate in the prevention and treatment of COVID-19, if shown to have benefit in this illness. Nitazoxanide has *in vitro* activity against the novel coronavirus SARS-CoV-2, which causes coronavirus disease (COVID-19). The EC50 on Vero E6 cells (the concentration to decrease the cytopathic effect of the virus by 50%) was reportedly 2.12µM (1). Nitazoxanide has a molar mass of 307.283g/mmol, and as such 2.12µM corresponds to a concentration of 0.65mg/L.

Although the above study on SARS-CoV-2 used nitazoxanide, the active metabolite tizoxanide inhibits influenza, [respiratory syncytial virus](https://www.sciencedirect.com/topics/medicine-and-dentistry/human-respiratory-syncytial-virus), [parainfluenza](https://www.sciencedirect.com/topics/medicine-and-dentistry/parainfluenza-virus-infection), coronavirus, [rotavirus](https://www.sciencedirect.com/topics/medicine-and-dentistry/rotavirus), [norovirus](https://www.sciencedirect.com/topics/medicine-and-dentistry/norovirus), hepatitis B, hepatitis C, [dengue](https://www.sciencedirect.com/topics/medicine-and-dentistry/dengue-fever), yellow fever, [Japanese encephalitis virus](https://www.sciencedirect.com/topics/medicine-and-dentistry/japanese-encephalitis-virus) and human immunodeficiency virus *in vitro* (2). Nitazoxanide was able to protect 90% of mice from death in a lethal challenge model of Japanese Encephalitis (3). The exact mechanism by which nitazoxanide exhibits its antiviral effects are uncertain, but augmented host responses as opposed to direct antiviral effects are postulated. Nitazoxanide is able to increase interferon responses in HCV/HIV coinfected individuals (4).

A double-blind randomised placebo-controlled trial of nitazoxanide in acute uncomplicated influenza found a statistically decreased duration of symptoms in the 600mg BD nitazoxanide group, but not the 300mg BD one (5). Additionally, viral titres were statistically decreased in the 600mg BD nitazoxanide group. Another study using the same doses in cases of severe respiratory infection caused by all respiratory viruses, did not show a statistical effect (6). Nitazoxanide is being assessed in clinical trials in COVID-19, and published results suggest an increased time to viral clearance, although this result may be confounded by differences in initial viral loads (7).

Composition & dose

One tablet of nitazoxanide as ALINIA® tablets contain 500 mg of nitazoxanide (<https://www.accessdata.fda.gov/drugsatfda_docs/label/2016/021497s001,021498s004lbl.pdf>).

Nitazoxanide will either be obtained by the sponsor or the local teams from a reliable manufacturer.

High dose nitazoxanide is given as 1.5 g bd.

The dose in this trial is: **1.5g PO twice daily for 7 days** (taken with food).

Pharmacokinetic characteristics

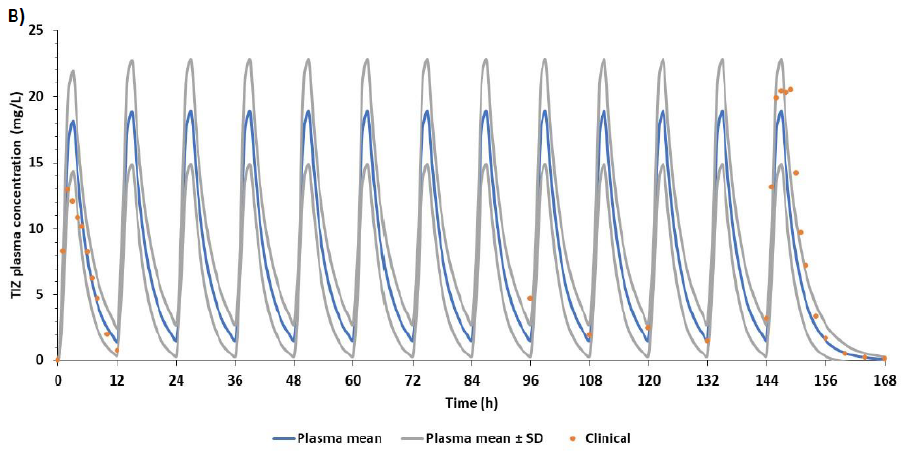
Following oral administration of nitazoxanide, the drug is rapidly hydrolysed to its active constituent, tizoxanide, and then conjugated to produce active tizoxanide glucuronide; nitazoxanide is not detected in plasma and > 99% of tizoxanide is bound to proteins. The bioavailabilities of tizoxanide and tizoxanide glucuronide are increased 2-fold with food. The mean Tmax of tizoxanide, is 2.5h with a half-life of 1 to 2h (8). The mean Tmax of the tizoxanide glucuronide is 4 to 6h. A dose of 1g PO BD of nitazoxanide produces a Cmax of 24mg/ml of tizoxanide and 26.4mg/ml tizoxanide glucuronide and the half-lives increase to 6.4 and 3.5 hours, respectively. Tizoxanide is excreted in the urine, bile and faeces, and tizoxanide glucuronide is excreted in urine and bile (its concentration is x10 that of plasma).

Pharmacokinetic sampling schedule: (only if there is evidence of accelerated viral clearance)

All patients will need to be admitted for 12 hours for intense sampling. The timings are:

- Pre-dose D0H0, then
- 1, 2, 3, 4, 6, 9, 12 and 24h

Note that blood will be taken on D3 and D7 for routine haematology and biochemistry. The EDTA sample will be used to measure the drug concentrations.

Toxicity

Nitazoxanide is a safe and well-tolerated drug. Reported toxicities include:

Gastrointestinal disorders: diarrhoea, gastroesophageal reflux disease

Nervous System disorders: dizziness

Respiratory, thoracic and mediastinal disorders: dyspnoea

Skin and subcutaneous tissue disorders: rash, urticaria

Risks related to nitazoxanide are very low. This is a very safe and generally well tolerated medication but adverse reactions relating to the central nervous system (headache and rare reports of dizziness), the gastrointestinal system (abdominal pain and nausea and rare reports of diarrhoea and gastro-oesophageal reflux), and genitourinary system (urine discolouration) have been described as well as post-marketing cases of dyspnoea, skin rash and urticaria. In one study, side effects of nitazoxanide did not differ from those of the placebo when used for the treatment of giardiasis, and does not cause side effects when used in healthy adults (9). This lack of significant side-effects is seen in doses up to 4g in humans. The LD50 of various animals is 10g/Kg. A study of 365 patients with AIDS receiving nitazoxanide 500-1500mg twice daily for a median duration of 62 days (1-1528 days) concluded that “No safety issues were identified at doses up to 3000 mg/day or for long durations of treatment” (10). These risks will be mitigated by excluding participation if people have had a previous serious adverse reaction to nitazoxanide.

Nitazoxanide may be used after the first trimester of pregnancy with severe symptoms of cryptosporidiosis. Adverse events have not been observed in animal reproduction studies although human data are not available. It is not known whether nitazoxanide enters into breast milk, and the manufacturer suggests balancing risk of exposure to the infant and benefit to the mother. We will exclude pregnant women and women breastfeeding from this study and ask female patients of child-bearing age to undergo a pregnancy test and take precautions not to become pregnant (contraception). Those who do become pregnant will be followed up. However, given the lack of evidence of harmful effects and its use later in pregnancy, the risk to a foetus seems low. The mitigating factors of a pregnancy test and advising contraception will make the likelihood of pregnancy within the trial and potential risk, even lower.

Contraindications and cautions

Known hypersensitivity. The Pharmacokinetics of nitazoxanide in patients with compromised renal or hepatic function have not been studied.

Relevant Drug interactions

None

References

1. Rossignol JF. Nitazoxanide: a first-in-class broad-spectrum antiviral agent. Antiviral Res. 2014;110:94-103.
2. Shi Z, Wei J, Deng X, Li S, Qiu Y, Shao D, et al. Nitazoxanide inhibits the replication of Japanese encephalitis virus in cultured cells and in a mouse model. Virol J. 2014;11:10.
3. Petersen T, Lee YJ, Osinusi A, Amorosa VK, Wang C, Kang M, et al. Interferon Stimulated Gene Expression in HIV/HCV Coinfected Patients Treated with Nitazoxanide/Peginterferon-Alfa-2a and Ribavirin. AIDS Res Hum Retroviruses. 2016;32(7):660-7.
4. El-Kowrany SI, El Ghaffar AEA, Shoheib ZS, Mady RF, Gamea GAM. Evaluation of nitazoxanide as a novel drug for the treatment of acute and chronic toxoplasmosis. Acta tropica. 2019;195:145-54.
5. Haffizulla J, Hartman A, Hoppers M, Resnick H, Samudrala S, Ginocchio C, et al. Effect of nitazoxanide in adults and adolescents with acute uncomplicated influenza: a double-blind, randomised, placebo-controlled, phase 2b/3 trial. Lancet Infect Dis. 2014;14(7):609-18.
6. Gamino-Arroyo AE, Guerrero ML, McCarthy S, Ramirez-Venegas A, Llamosas-Gallardo B, Galindo-Fraga A, et al. Efficacy and Safety of Nitazoxanide in Addition to Standard of Care for the Treatment of Severe Acute Respiratory Illness. Clin Infect Dis. 2019;69(11):1903-11.
7. Rocco PRM, Silva PL, Cruz FF, Junior M, Tierno P, Moura MA, et al. Early use of nitazoxanide in mild Covid-19 disease: randomised, placebo-controlled trial. Eur Respir J. 2021. Jan 14:2003725. doi: 10.1183/13993003.03725-2020.
8. Stockis A, Deroubaix X, Lins R, Jeanbaptiste B, Calderon P, Rossignol JF. Pharmacokinetics of nitazoxanide after single oral dose administration in 6 healthy volunteers. Int J Clin Pharmacol Ther. 1996;34(8):349-51.
9. Stockis A, Allemon AM, De Bruyn S, Gengler C. Nitazoxanide pharmacokinetics and tolerability in man using single ascending oral doses. Int J Clin Pharmacol Ther. 2002;40(5):213-20.
10. Rossignol JF. Nitazoxanide in the treatment of acquired immune deficiency syndrome-related cryptosporidiosis: results of the United States compassionate use program in 365 patients. Aliment Pharmacol Ther. 2006;24(5):887-94.

#### 1.4. REGN-COV2

Rationale

REGN-COV2 is an antibody cocktail of two anti-SARS-CoV-2 neutralising antibodies Casirivimab and Imdevimab, which has been shown to reduce viral load in patients with COVID-19 (1). On the 21^st^ of November 2020 the US Food and Drug Administration (FDA) issued an Emergency Use Authorization (EUA) in people who are older than 12 years, weigh more than 40Kg and were at high risk of progression to severe disease. The drug is currently being evaluated in the RECOVERY trial in late-stage hospitalised patients, but as with all antiviral drugs, the likely window of benefit is earlier in disease, when viral burdens are higher and before people are hospitalised, when inflammation predominates. Although REGN-COV-2 is currently expensive and requires IV administration- thereby limiting its use globally, this antibody cocktail currently has the best evidence for an antiviral effect, and will form the positive control for other interventions (i.e. interventions can be assessed in relation to the likely antiviral effect observed in this positive control arm). The hope would be that the cost, availability and feasibility of administration will improve and that this may become a more widely available intervention for COVID-19, which can also be used in LMICs. Unfortunately, the potential benefit of antibody therapy, including REGN-COV-2 is threatened by the appearance of mutant viruses, and there is now evidence of decreased binding affinity for the receptor binding domain (RBD) of Casirivimab, of the South Africa variant with the K417N and/or E484K mutations. As such the study will additionally allow monitoring of the effect of this antibody cocktail on viral clearance over time and with the appearance of different viral mutants, informing its on-going utility.

Composition & dose

The dosage in adults and in pediatric patients (12 years of age and older weighing at least 40 kg) is 600 mg of casirivimab and 600 mg of imdevimab administered together as a single intravenous infusion over at least 60 minutes. Casirivimab and imdevimab solutions must be diluted prior to administration. Casirivimab and imdevimab should be given together as soon as possible after positive results of direct SARS-CoV-2 viral testing and within 10 days of symptom onset.

The dose in this trial is: **600mg casirivimab/ 600mg imdevimab given once on D0**

REGN-COV2 will either be obtained by the sponsor or the local teams from the manufacturer.

For preparation and administration instructions please see: <https://www.fda.gov/media/143892/download>

Pharmacokinetic characteristics

Pharmacokinetic profiles of casirivimab and imdevimab are expected to be consistent with the profile of other IgG1 mAbs.

Specific Populations- the effect of different covariates (e.g., age, sex, race, body weight, disease severity, hepatic impairment) on the pharmacokinetics of casirivimab and imdevimab is unknown. Renal impairment is not expected to impact the pharmacokinetics of casirivimab and imdevimab, since mAbs with molecular weight >69 kDa are known not to undergo renal elimination.

Drug-Drug Interactions- casirivimab and imdevimab are mAbs which are not renally excreted or metabolized by cytochrome P450 enzymes; therefore, interactions with concomitant medications that are renally excreted or that are substrates, inducers, or inhibitors of cytochrome P450 enzymes are unlikely

Pharmacokinetic schedule: only if there is evidence of accelerated viral clearance

As a type B intervention (positive control), intensive pharmacokinetic sampling will not be conducted on those receiving REGN-COV2.

Toxicity

There is a potential for serious hypersensitivity reaction, including anaphylaxis, with administration of casirivimab and imdevimab. If signs or symptoms of a clinically significant hypersensitivity reaction or anaphylaxis occur, immediately discontinue administration and initiate appropriate medications and/or supportive therapy. Infusion-related reactions have been observed with administration of casirivimab and imdevimab. Signs and symptoms of infusion-related reactions may include: fever, chills, nausea, headache, bronchospasm, hypotension, angioedema, throat irritation, rash including urticaria, pruritus, myalgia, dizziness. If an infusion-related reaction occurs, consider slowing or stopping the infusion and administer appropriate medications and/or supportive care.

Contraindications and Cautions

There are no contraindications.

Reference

1. Weinreich DM, Sivapalasingam S, Norton T, Ali S, Gao H, Bhore R, et al. REGN-COV2, a Neutralizing Antibody Cocktail, in Outpatients with Covid-19. New England Journal of Medicine. 2020;384(3):238-51.

#### 16.8 Nebulised Unfractionated Heparin (UFH)

Rationale

Nebulised UFH has demonstrated antiviral, anti-inflammatory, anticoagulant and mucolytic effects and is being trialled in various phases of COVID-19. In this trial, we aim to primarily establish the antiviral effect of this drug *in vivo*, although other effects of this drug may also be clinically relevant in COVID-19. In laboratory experiments, UFH blocked the binding and infectivity of SARS-CoV-2 to human bronchial epithelial cells and in a Vero E6 cell line, inhibition of SARS-CoV-2 infection occurred at therapeutically relevant concentrations. Use of UFH via the nebulisation allows for higher concentrations to be delivered to infected tissues, without concerns re: systemic effects of anticoagulation. UFH is a cheap and widely available drug which can be given nebulised in the community. If shown to be effective, this treatment may be studied in phase 3 studies to determine the effect on clinical endpoints, and affordable and widely available nebulising systems can be developed.

Composition & dose

25000 IU UFH every 6 hours using a vibrating mesh nebuliser for 7 days.

Pharmacokinetic characteristics

UFH is rapidly taken up by respiratory endothelium and clinically significant systemic effects are not expected.

Pharmacokinetic schedule: only if there is evidence of accelerated viral clearance

As we are using the nebulised route of administration, high levels of drug will be expected in the oropharynx and respiratory tract, but negligible levels in the systemic circulation. An intensive pharmacokinetic schedule will not be conducted.

Toxicity

Toxicity relating to use of UFH normally relates to systemic effects of the drug on coagulation (through prolongation of the aPTT and increased likelihood of bleeding) and reactions to the drug. At the proposed dose used in this study, nebulised UFH only modestly increases peak aPTT in patients who concomitantly receive IV or SC UFH and has no effect on the peak APTT in patients who receive IV or SC LMWH.

Contraindications (1)

- Heparin allergy or Heparin-induced thrombocytopaenia
- Known prolonged APTT > 120ms or bleeding diathesis
- Pulmonary bleeding or uncontrolled bleeding
- Platelet count < 20 x 10^9^ /L
- Pregnant

Cautions

In addition to those with the above contraindications, caution should be taken by those using the nebulised system, as this may cause aerosolisation of virus and potentially increase the likelihood of transmission. As such, nebulisation if it occurs at the ward should occur in a negative pressure room with appropriate infection control procedures. If nebulisation occurs at home, this should be done in a room with adequate ventilation to the outside, where the patient can guarantee that the room will not be entered by any other persons for at least 2 hours after nebulisation occurs. Ideally this will be conducted outside, at least 50 metres from the nearest person.

Drug interactions

The listed drug interactions for UFH relate to systemic use and are not relevant to its use as nebulised therapy. Those taking drugs with an increased risk of bleeding should be assessed and potentially excluded.

Reference: <https://www.fda.gov/media/143892/download>

Reference

1. van Haren FMP, Richardson A, Yoon HJ, Artigas A, Laffey JG, Dixon B, Smith R, Vilaseca AB, Barbera RA, Ismail TI et al: INHALEd nebulised unfractionated HEParin for the treatment of hospitalised patients with COVID-19 (INHALE-HEP): Protocol and statistical analysis plan for an investigator-initiated international metatrial of randomised studies. Br J Clin Pharmacol 2020.

#### 16.5 Favipiravir

Rationale

Favipiravir is a small molecule antiviral which was developed in 2002 as an inhibitor of viral replication of influenza (1). Favipiravir is a pyrazinecarboxamide derivative, thought to act as a selective viral RNA-dependent RNA polymerase inhibitor. Favipiravir is a prodrug which is metabolized to its active form, favipiravir-ribofuranosyl-5’-triphosphate. Favipiravir has demonstrated *in vitro* activity against SARS-CoV-2 (2). An early open labelled control study compared Favipiravir and Lopinavir/ritonavir. It found that the favipiravir arm had a shorted viral clearance median time and significant improvement in chest CT compared to the control arm (improvement rate 91.43% compared to 62.22% - p=0.004) (3). A systematic review and meta-analysis were performed by Hassanipour et.al. (4). It showed a significant clinical improvement in favipiravir compared to control during the 7 days post hospitalization (RR = 1.24, 95% CI: 1.09–1.41; P = 0.001). Viral clearance was higher with favipiravir but it was not significant. Transfer to ICU and adverse events were not found to be significant. In those with mild to moderate COVID-19 there was no beneficial effect to mortality.

Composition & dose

In this study, patients will be given 1800mg of favipiravir (nine 200mg tablets) at H0 and 1800mg of favipiravir (9 200mg tablet) at H12. Thereafter the patient will take 800mg BD (four 200mg tablets) for a further 6 days.

Favipiravir will either be obtained by the sponsor or the local teams from a reliable manufacturer.

Pharmacokinetic characteristics

Oral favipiravir reaches maximal concentrations at 2 hours post oral administration and has a half-life of 2-5.5 hours. Protein binding occurs at 54%. The active parent drug is metabolised to the inactive metabolite T-705M1 mainly by aldehyde oxidase and partially by xanthine oxidase. PK analysis of continuous doses of favipiravir in nonhuman primate models exhibit nonlinear pharmacokinetics over time, with decreasing trough levels at D4, relative to D2.

Pharmacokinetic schedule: only if there is evidence of accelerated viral clearance

All patients will need to be admitted for 12 hours for intense sampling. The timings are:

- Pre-dose D0H0, then
- 1, 2, 3, 4, 6, 9, 12 and 24h

Note that blood will be taken on D3 and D7 for routine haematology and biochemistry. The EDTA sample will be used to measure the drug concentrations.

Toxicity

A review of the safety of favipiravir from 29 studies demonstrates a favourable safety profile. Side-effects which tend to be mild, include diarrhoea, as well as blood abnormalities including raised uric acid levels, low neutrophil count and raised liver function tests. Early concerns regarding QTc prolongation, were not born out in a study of healthy Japanese individuals using moxifloxacin as a positive control, where favipiravir showed no effect on QT intervals.

Animal models have demonstrated teratogenicity, and no human studies on pregnant or lactating women have been conducted.

Contraindications & cautions

Known hypersensitivity. See later section on drug-specific contraindications.

Patients should avoid excessive exposure to sunlight or artificial UV light

Drug interactions

Favipiravir inhibits the formation of acetaminophen sulphate *in vitro* and *in vivo*, resulting in a small magnitude increase in acetaminophen levels. As a result daily acetaminophen (paracetamol) should not exceed 3g/ day (5).

Reference

1. Furuta, Y., Takahashi, K., Fukuda, Y., Kuno, M., Kamiyama, T., Kozaki, K., Nomura, N., Egawa, H., Minami, S., Watanabe, Y., Narita, H. and Shiraki, K., 2002. In Vitro and In Vivo Activities of Anti-Influenza Virus Compound T-705. Antimicrobial Agents and Chemotherapy, 46(4), pp.977-981.
2. Wang M, Cao R, Zhang L, Yang X, Liu J, Xu M et al. Remdesivir and chloroquine effectively inhibit the recently emerged novel coronavirus (2019-nCoV) in vitro. Cell Research. 2020;30(3):269-271.
3. Cai, Q., Yang, M., Liu, D., Chen, J., Shu, D., Xia, J., Liao, X., Gu, Y., Cai, Q., Yang, Y., Shen, C., Li, X., Peng, L., Huang, D., Zhang, J., Zhang, S., Wang, F., Liu, J., Chen, L., Chen, S., Wang, Z., Zhang, Z., Cao, R., Zhong, W., Liu, Y. and Liu, L., 2020. Experimental Treatment with Favipiravir for COVID-19: An Open-Label Control Study. Engineering, 6(10), pp.1192-1198.
4. Hassanipour, S., Arab-Zozani, M., Amani, B., Heidarzad, F., Fathalipour, M. and Martinez-de-Hoyo, R., 2021. The efficacy and safety of Favipiravir in treatment of COVID-19: a systematic review and meta-analysis of clinical trials. Scientific Reports, 11(1).
5. Zhao Y, Harmatz JS, Epstein CR, Nakagawa Y, Kurosaki C, Nakamura T, Kadota T, Giesing D, Court MH, Greenblatt DJ. Favipiravir inhibits acetaminophen sulfate formation but minimally affects systemic pharmacokinetics of acetaminophen. Br J Clin Pharmacol. 2015 Nov;80(5):1076-85. doi: 10.1111/bcp.12644. Epub 2015 Jun 8. PMID: 25808818; PMCID: PMC4631180.

#### 16.9 Molnupiravir

Rationale

Molnupiravir is a small molecule ribonucleoside prodrug of N-Hydroxycytidine (NHC). It is phosphorylated intracellularly into NHC triphosphate (NHC-TP). NHC-TP incorporates into viral RNA by viral RNA polymerase causing an accumulation of deleterious errors into the viral genome. This causes “error catastrophe” by increasing the rate of mutation in the viral genome to a level lethal to the virus and causing extinction (1).

Studies of mice infected with SARS-CoV-1 found that EDII-2801 (an orally bioavailable NHC-prodrug) prophylactically and therapeutically improved pulmonary function, and reduced virus titre (2). An *in vivo* study on human lung-only mice found that molnupiravir dramatically inhibited the replication of SARS-CoV-2 (3). A Phase 1 safety trial of doses up to 1600MG per day found molnupiravir was well tolerated with the incidence of adverse effects was highest in the placebo arm (4). Another Phase 1 trial concluded molnupiravir was safe and well tolerated and advised 800MG BD for 5 days (5).

A Phase 2 trial compared viral clearance to a range of doses and found that by day 5, virus was not isolated for those receiving 400 or 800MG BD of molnupiravir (0/42 and 0/53 respectively), compared to 11.1% (6/54) in the placebo group (p = 0.03).

The drug manufacturer, Merck, Sharpe & Dohme (MSD), performed a double blind randomised, placebo-controlled trial, which gave molnupiravir within 5 days of symptoms to non-hospitalised, unvaccinated adults with mild/moderate COVID-19 and at least 1 risk factor for severe COVID-19. Dose was 800MG BD for 5 days. Primary efficacy end point was hospitalisation due to any cause or death at day 29. Interim results showed that 28 out of 285 (7.3%) patients given molnupiravir had been hospitalised or died, compared to 53 out of 377 (14.1%) in the placebo group (-6.8% difference, −11.3 to −2.4; P=0.001). In addition the all-randomised analysis also showed that molnupiravir treatment was associated with greater reductions from baseline in mean viral load than placebo at days 3, 5 and 10 (6).

However, when the data was presented to the FDA for approval, further analysis showed lower effectiveness. A press release showed data from all enrolled patients and found that the risk of hospitalisation in the molnupiravir group was 9.7% (68/699) compared to 6.8% (48/709) in the control group. The absolute risk reduction was 3.0% (0.1 to 5.9; p-value =0.0218). A 50% decrease from the interim results (7).

Composition & dose

Each Lagevrio® capsule contains 200 mg of molnupiravir, recommended dose is 800mg taken orally every 12 hours for 5 days. Molnupiravir is being made by MSD, as well as under license by a number of generics manufacturers.

Molnupiravir will either be obtained by the sponsor or the local teams from a reliable manufacturer (including high-quality generics manufacturers).

Should be given within 5 days of onset of symptoms.

Pharmacokinetic characteristics

Molnupiravir is a prodrug metabolised to the ribonucleoside analogue N-Hydroxycytidine (NHC) and phosphorylated to the active ribonucleoside triphosphate (NHC-TP). Following twice daily oral administration of 800 mg molnupiravir, median time to peak plasma NHC concentrations is 1.5 hours. The effective half-life of NHC is approximately 3.3 hours. It can be taken with or without food.

Pharmacokinetic sampling schedule: (only if there is evidence of accelerated viral clearance)

The timings are:

- Pre-dose D0H0, then
- 1, 2, 3, 4, 6, 9, 12 and 24h post dose

Note that blood will be taken on D3 and D7 for routine haematology and biochemistry. The EDTA sample will be used to measure the drug concentrations.

Toxicity

Highest dose tested in safety trials was 800MG BD, found to have a 0.9% probability of having 30% excess toxicity compared to control therefore considered safe (5).

Contraindications

Hypersensitivity

Cautions

No dose adjustment needed for renal impairment however the pharmacokinetics have not been evaluated with eGFR <30 mL/min, also not evaluated with hepatic impairment.

As per USA FDA advice, those on molnupiravir need to be aware of avoiding pregnancy. Females of child bearing potential need to use a reliable method of contraception for the duration of the treatment, and also for 4 days after the final dose of molnupiravir. Males of reproductive potential, if they are sexually active with females of child bearing potential, should use a reliable form of contraception during the treatment and at least 3 months after the final dose.

Therefore, in addition to the baseline pregnancy test, a repeat test on women of childbearing potential who are allocated to Molnupiravir will be done on D14.

Drug Interactions

None identified

References

1.	Singh AK, Singh A, Singh R, Misra A. Molnupiravir in COVID-19: A systematic review of literature. Diabetes & Metabolic Syndrome: Clinical Research & Reviews. 2021;15(6):102329.

2.	Sheahan TP, Sims AC, Zhou S, Graham RL, Hill CS, Leist SR, et al. An orally bioavailable broad-spectrum antiviral inhibits SARS-CoV-2 and multiple endemic, epidemic and bat coronavirus. Biorxiv. 2020.

3.	Wahl A, Gralinski LE, Johnson CE, Yao W, Kovarova M, Dinnon KH, et al. SARS-CoV-2 infection is effectively treated and prevented by EIDD-2801. Nature. 2021;591(7850):451-7.

4.	Painter WP, Holman W, Bush JA, Almazedi F, Malik H, Eraut NC, et al. Human safety, tolerability, and pharmacokinetics of molnupiravir, a novel broad-spectrum oral antiviral agent with activity against SARS-CoV-2. Antimicrobial agents and chemotherapy. 2021;65(5):e02428-20.

5.	Khoo SH, Fitzgerald R, Fletcher T, Ewings S, Jaki T, Lyon R, et al. Optimal dose and safety of molnupiravir in patients with early SARS-CoV-2: a Phase I, open-label, dose-escalating, randomized controlled study. Journal of Antimicrobial Chemotherapy. 2021;76(12):3286-95.

6.	Jayk Bernal A, Gomes da Silva MM, Musungaie DB, Kovalchuk E, Gonzalez A, Delos Reyes V, et al. Molnupiravir for oral treatment of Covid-19 in nonhospitalized patients. New England Journal of Medicine. 2021.

7.	Merck and Ridgeback Biotherapeutics Provide Update on Results from MOVe-OUT Study of Molnupiravir, an Investigational Oral Antiviral Medicine, in At Risk Adults With Mild-to-Moderate COVID-19 [press release]. 26/November/2021 2021.

#### 16.9  Nirmatrelvir/ritonavir (e.g. PAXLOVID™)

Rationale

PAXLOVID™ is a therapeutic combination of 2 compounds, Nirmatrelvir and ritonavir, taken at the same time as separate pills. Nirmatrelvir (PF-07321332) is an oral covalent 3CL protease inhibitor of SARS-CoV-2 and ritonavir is an inhibitor of HIV-1 and HIV-2 protease. Ritonavir inhibits cytochrome P450A and CYP2D6 which inhibits the metabolism of nirmatrelvir.

Coronoviruses contain two proteases, main protease – Mpro (also known as 3CL protease – 3CLpro) and papain-like protease. Pfizer developed a SARS-CoV Mpro inhibitor in 2002 (PF-00835231) which demonstrated in vitro and in vivo activity against SARS-CoV-2 (1). Other studies also found successful in vitro inhibition of SARS-CoV-2 replication in derived alveolar basal epithelial cells expressing ACE2 (2) and human bronchial epithelial cells (3). This compound had insufficient bioavailability so the oral version PF-07321332 was developed (4).

A Phase 2/3 EPIC-HR randomised double blind control trial from Pfizer looked at non-hospitalised patients at risk of progressing to severe illness. Intervention was ritonavir and nirmatrelvir started within three or five days of symptom onset. Primary endpoint was COVID-19-related hospitalisation or all cause death up to day 28. For those receiving PAXLOVID™ within 3 days, 3/389 (0.7%) were hospitalised with zero deaths., compared to the placebo 27/385 hospitalised with 7 deaths, p<0.0001. For treatment within 5 days, 6/607 (6.7%) were hospitalised with no deaths, compared to 41/612 (with 10 deaths) in the placebo group (p<0.0001) (5).

Composition & dose

Administered as 300MG of nirmatrelvir (two 150MG tablets) with one 100 MG tablet of ritonavir given twice daily for 5 days.

Nirmatrelvir/ritonavir will either be obtained by the sponsor or the local teams from a reliable manufacturer (including high-quality generics manufacturers).

Pharmacokinetic characteristics

Ritonavir is administered with nirmatrelvir as a pharmacokinetic enhancer resulting in higher systemic concentrations and longer half-life of nirmatrelvir. This supports a twice daily administration. Nirmatrelvir (when given with ritonavir) is eliminated renally, with a half-life of 6.05 hours, peak concentration is at 3 hours. Ritonavir is metabolised in the liver, with a half-life of 6.15 hours, peak concentration is at 3.98 hours.

Pharmacokinetic sampling schedule: (only if there is evidence of accelerated viral clearance)

The timings are:

- Pre-dose D0H0, then
- 1, 2, 3, 4, 6, 9, 12 and 24h post dose

Note that blood will be taken on D3 and D7 for routine haematology and biochemistry. The EDTA sample will be used to measure the drug concentrations.

Toxicity

Repeat-dose toxicity studies up to 1 month duration of nirmatrelvir in rats and monkeys resulted in

no adverse findings. Ritonavir can cause retinal toxicity with long term use (6), although this is not relevant for the short-term use in the study. Ritonavir alone does not cause any issues related to HIV drug-resistance.

Contraindications

In our study, nirmatrelvir/ritonavir is contraindicated in:

hypersensitivity to nirmatrelvir/ritonavir or any of its ingredients, including ritonavir.

co-administration with drugs highly dependent on CYP4503 A.

co-administration with potent CYP450 3A inducers

More details regarding concomitant medication can be found at:

<https://www.covid19-druginteractions.org/checker>

<https://crediblemeds.org/>

<https://compendium.ch/fr/Patient> - this is a Swiss website dealing with drug interactions

<https://www.bnf.org/products/bnf-online/> - available only in the UK

Cautions

Caution should be given in patients with pre-existing liver disease/hepatitis or live enzyme abnormalities. Dose reduction for renal impairment (eGFR ≥ 30 to <60 mL/min) is 150MG nirmatrelvir and 100 MG ritonavir.

Drug Interactions

Contraindicated with drugs that are highly dependent on CYP3A for clearanceor are potent CYP3A inducers such as

References

1.	Boras B, Jones RM, Anson BJ, Arenson D, Aschenbrenner L, Bakowski MA, et al. Discovery of a novel inhibitor of coronavirus 3CL protease as a clinical candidate for the potential treatment of COVID-19. BioRxiv. 2021:2020.09. 12.293498.

2.	de Vries M, Mohamed AS, Prescott RA, Valero-Jimenez AM, Desvignes L, O’Connor R, et al. A comparative analysis of SARS-CoV-2 antivirals characterizes 3CLpro inhibitor PF-00835231 as a potential new treatment for COVID-19. Journal of Virology. 2021;95(10):e01819-20.

3.	Barilli A, Visigalli R, Ferrari F, Di Lascia M, Riccardi B, Puccini P, et al. Organic Cation Transporters (OCTs) in EpiAirway™, a Cellular Model of Normal Human Bronchial Epithelium. Biomedicines. 2020;8(5):127.

4.	Owen DR, Allerton CM, Anderson AS, Aschenbrenner L, Avery M, Berritt S, et al. An oral SARS-CoV-2 Mpro inhibitor clinical candidate for the treatment of COVID-19. Science. 2021:eabl4784.

5.	Mahase E. Covid-19: Pfizer’s paxlovid is 89% effective in patients at risk of serious illness, company reports. British Medical Journal Publishing Group; 2021.

6.	Tu Y, Poblete RJ, Freilich BD, Zarbin MA, Bhagat N. Retinal toxicity with ritonavir. International journal of ophthalmology. 2016;9(4):640.

#### 16.9  Sotrovimab

Rationale

Sotrovimab (VIR-7831) is an Fc-engineered IgG1 human monoclonal antibody developed from a parental antibody isolated from a survivor of the SARS outbreak in 2003 (1). Sotrovimab targets a highly conserved epitope in the SARS-CoV-2 spike protein at a region that does not compete with angiotensin-converting enzyme 2 binding.

In-vitro testing has shown that sotrovimab is able to retain activity against alpha, beta, gamma, delta and kappa variants, and even omicron pseudotyped virus encoding the then most prevalent haplotype of the Omicron spike. The same study showed that Syrian Golden hamsters infected with wildtype SARS-CoV-2 had significantly decreased total viral load and infectious virus levels with sotrovimab (2).

A multicentre randomized, double-blind, multicentre, placebo-controlled, phase 3 study tested sotrovimab on non-hospitalised patients with symptomatic, mild to moderate COVID-19 and at least 1 risk factor for disease progression. Symptom onset was within 5 days, median age was 53. Primary efficacy outcome was all-cause hospitalization longer than 24 hours for acute illness management or death through day 29. Primary outcome was significantly reduced with sotrovimab (6/528 – 1%) compared to placebo (30/529 – 6%) by 79% (95% CI, 50% – 91%; P<.001) (3).

N.B. – REGN-COV2 (the initial positive control monoclonal antibody) was shown in vitro studies to fail to neutralise omicron (4), although the in-vivo significance of this is yet to be determined.

Composition & dose

Recommended dose is 500MG sotrovimab administered as a single intravenous infusion over 60 minutes. Patients should be monitored for at least 1 hour after administration.

0.9% Sodium Chloride of 5% dextrose for injection, one vial of sotrovimab is 500MG/8ML.

Pharmacokinetic characteristics

The mean systemic clearance is 125 mL/day, and the median terminal half-life is approximately 49 days. The geometric mean maximum plasma concentration (Cmax) after a 1-hour sotrovimab intravenous infusion to be 117.6 mcg/mL, and the geometric mean Day 29 concentration to be 24.5 mcg/mL.

Toxicity

Serious hypersensitivity reactions, including anaphylaxis, have been reported with administration of

sotrovimab. If signs or symptoms of a clinically significant hypersensitivity reaction or anaphylaxis occur during infusion, immediately discontinue administration and initiate appropriate medications and/or supportive care.

Contraindications

Hypersensitivity

Cautions

Renal impairment is not expected to impact the pharmacokinetics of sotrovimab since monoclonal antibodies with molecular weight >69 kDa do not undergo renal elimination. The effect of hepatic insufficiency is unknown. The effect of other covariates (e.g., sex, race, body weight, disease of severity on the pharmacokinetics is also unknown.

Pharmacokinetic schedule: only if there is evidence of accelerated viral clearance

As a type B intervention (positive control), intensive pharmacokinetic sampling will not be conducted on those receiving sotrovimab.

Drug Interactions

No formal drug interaction studies have been performed with sotrovimab. Sotrovimab is not renally

excreted or metabolized by cytochrome P450 (CYP) enzymes; therefore, interactions with concomitant medications that are renally excreted or that are substrates, inducers, or inhibitors of CYP enzymes are unlikely.

References

1.	Pinto D, Park Y-J, Beltramello M, Walls AC, Tortorici MA, Bianchi S, et al. Cross-neutralization of SARS-CoV-2 by a human monoclonal SARS-CoV antibody. Nature. 2020;583(7815):290-5.

2.	Cathcart AL, Havenar-Daughton C, Lempp FA, Ma D, Schmid M, Agostini ML, et al. The dual function monoclonal antibodies VIR-7831 and VIR-7832 demonstrate potent in vitro and in vivo activity against SARS-CoV-2. bioRxiv. 2021.

3.	Gupta A, Gonzalez-Rojas Y, Juarez E, Casal MC, Moya J, Falci DR, et al. Effect of the Neutralizing SARS-CoV-2 Antibody Sotrovimab in Preventing Progression of COVID-19: A Randomized Clinical Trial. medRxiv. 2021.

4.	Wilhelm A, Widera M, Grikscheit K, Toptan T, Schenk B, Pallas C, et al. Reduced neutralization of SARS-CoV-2 omicron variant by vaccine sera and monoclonal antibodies. MedRxiv. 2021.

#### 16.9  Fluoxetine

Rationale

Fluoxetine is a selective serotonin reuptake inhibitor introduced in the 1980s and the most globally widely used antidepressant (1). Recently it has been classified as a Functional Inhibitor of Acid SphingoMyelinAse (FIASMA) and considered for anti-viral properties. Acid SphingoMyelinAse is a glycosome which catalyses the degradation of sphingomyelin to phosphorycholine and ceramide; ceramide causes membrane changes mediating viral uptake (2). ASM has shown to be activated by SARS-CoV-2. FIASMA’s have been demonstrated to displace ASM from lysosomal membranes, causing partial degradation, reducing the viral membrane fusion with the host cell (3).

Fluoxetine has been shown to have *in vitro* activity against SARS-CoV-2 with an EC50 of 2.6 µM demonstrated at a concentration equivalent to normal treatment for depression (0.8ug/ml) (4). Fluoxetine has also been shown to be effective against the alpha and beta variants (5), and can be effective without cytotoxic effect (6). *in silico* data determined that 40MG per day would provide at least 85% of patients with the trough target plasma concentrations needed to reach the target EC90 concentration within 3 days (7).

Retrospective observational tries have demonstrated the potential use of fluoxetine against COVID-19. Patients already on fluoxetine taking it within the first 48 hours of admission were found to have a lower risk of intubation or death (combined endpoint) (8). Another study expanded this to within 10 days before or 7 days after COVID-19 diagnosis and found a reduction in death with fluoxetine (9). Finally, a Hungarian study gave anxious patients fluoxetine on admission and found in the fluoxetine group 13.6% died compared to 30.8% not receiving fluoxetine (10).

There have been two clinical trials on another FIASMA – fluvoxamine. A double blind randomised controlled trial of 152 participants used a fluvoxamine dose of 100MG TDS for 15 days for SARS-CoV-2 non-hospitalised patients. Of the 80 participants that received fluvoxamine, none of them died, compared to 6 in the control group (11). The TOGETHER trial used a dose of 100MG BD for 10 days, 741 received fluvoxamine and 756 received the placebo. There were 17 deaths in the fluvoxamine group compared to 25 in the placebo, p=0.24 (12). Hospitalisation was reduced with fluvoxamine, however the endpoint defined staying in the emergency department for > 6 hours as hospitalisation, when this was removed the difference was not significant.

The ANTICOV clinical trial is proposing fluoxetine 40MG/day + inhaled budesonide for 7 days. In Silico study recommended 40 MG/OD dose with provide target plasma concentrations in 3 days.

Dose

In this study, patients will be given 40MG OD on D0 and for a further 6 days.

Pharmacokinetic characteristics

Fluoxetine is well absorbed after oral intake with a large volume of distribution. Following a single oral 40 mg dose, peak plasma concentrations of fluoxetine from 15 to 55 ng/mL are observed after 6 to 8 hours. Food does not appear to affect systemic bioavailability. Fluoxetine is metabolized extensively in the liver to norfluoxetine, which has similar pharmacological activity. The main route of elimination is hepatic, to inactive metabolites excreted by the kidney.

After acute administration, the elimination half-life of fluoxetine is 1 to 3 days and after chronic administration 4 to 6 days. Norfluoxetine is longer with an elimination half-life of 4 to 16 days after acute and chronic administration. Steady state plasma levels are attained after 4 to 5 weeks of continuous drug administration.

Pharmacokinetic sampling schedule: (only if there is evidence of accelerated viral clearance)

The timings are:

- Pre-dose D0H0, then
- 1, 2, 3, 4, 6, 9, 12 and 24h post dose

Note that blood will be taken on D3 and D7 for routine haematology and biochemistry. The EDTA sample will be used to measure the drug concentrations.

Adverse effects

Common adverse effects of fluoxetine affect the gastrointestinal system (nausea, diarrhoea and anorexia) and the nervous system (insomnia, somnolence and anxiety). Care must be taken in those with existing mental health problems. Serotonin syndrome can be prevented by avoiding overuse, and should not be an issue at these doses for these durations.

Contraindications

Known hypersensitivity

Use of MAOIs and thioridazine – fluoxetine must not be given within 14 days of discontinuation, and a 5-week gap must be given before starting an MAOI or thioridazine

Other antidepressants (e.g. Selective Serotonin Reuptake Inhibitors (SSRIs)), which increase the risk of serotonin syndrome.

Cautions

Since fluoxetine is metabolised in the liver hepatic impairment may cause a prolonged half-life.

Caution should be taken with those with a pre-existing psychiatric condition.

References

1.	Rossi A, Barraco A, Donda P. Fluoxetine: a review on evidence based medicine. Annals of General Hospital Psychiatry. 2004;3(1):1-8.

2.	Kornhuber J, Hoertel N, Gulbins E. The acid sphingomyelinase/ceramide system in COVID-19. Molecular Psychiatry. 2021:1-8.

3.	Carpinteiro A, Edwards MJ, Hoffmann M, Kochs G, Gripp B, Weigang S, et al. Pharmacological inhibition of acid sphingomyelinase prevents uptake of SARS-CoV-2 by epithelial cells. Cell Reports Medicine. 2020;1(8):100142.

4.	Zimniak M, Kirschner L, Hilpert H, Seibel J, Bodem J. The serotonin reuptake inhibitor Fluoxetine inhibits SARS-CoV-2. BioRxiv. 2020.

5.	Fred MS, Kuivanen S, Ugurlu H, Casarotto PC, Levanov L, Saksela K, et al. Antidepressant and antipsychotic drugs reduce viral infection by SARS-CoV-2 and fluoxetine show antiviral activity against the novel variants in vitro. bioRxiv. 2021.

6.	Schloer S, Brunotte L, Goretzko J, Mecate-Zambrano A, Korthals N, Gerke V, et al. Targeting the endolysosomal host-SARS-CoV-2 interface by clinically licensed functional inhibitors of acid sphingomyelinase (FIASMA) including the antidepressant fluoxetine. Emerging microbes & infections. 2020;9(1):2245-55.

7.	Eugene AR. Fluoxetine pharmacokinetics and tissue distribution suggest a possible role in reducing SARS-CoV-2 titers. F1000Research. 2021;10(477):477.

8.	Hoertel N, Sánchez-Rico M, Vernet R, Beeker N, Jannot A-S, Neuraz A, et al. Association between antidepressant use and reduced risk of intubation or death in hospitalized patients with COVID-19: results from an observational study. Molecular psychiatry. 2021:1-14.

9.	Oskotsky T, Marić I, Tang A, Oskotsky B, Wong RJ, Aghaeepour N, et al. Mortality Risk Among Patients With COVID-19 Prescribed Selective Serotonin Reuptake Inhibitor Antidepressants. JAMA network open. 2021;4(11):e2133090-e.

10.	Németh ZK, Szûcs A, Vitrai J, Juhász D, Németh JP, Holló A. Fluoxetine use is associated with improved survival of patients with COVID-19 pneumonia: A retrospective case-control study. Ideggyogyaszati Szemle. 2021;74(11-12):389-96.

11.	Lenze EJ, Mattar C, Zorumski CF, Stevens A, Schweiger J, Nicol GE, et al. Fluvoxamine vs placebo and clinical deterioration in outpatients with symptomatic COVID-19: a randomized clinical trial. Jama. 2020;324(22):2292-300.

12.	Reis G, dos Santos Moreira-Silva EA, Silva DCM, Thabane L, Milagres AC, Ferreira TS, et al. Effect of early treatment with fluvoxamine on risk of emergency care and hospitalisation among patients with COVID-19: the TOGETHER randomised, platform clinical trial. The Lancet Global Health. 2021.

#### 16.9  Fluvoxamine

Rationale

Fluvoxamine is a selective serotonin reuptake inhibitor developed in 1983 and approved by the U.S. FDA in 1994. It is used to treat major depressive disorder and Obsessive-Compulsive Disorder. Recently it has been classified as a Functional Inhibitor of Acid SphingoMyelinAse (FIASMA) and considered for anti-viral properties. Acid SphingoMyelinAse is a glycosome which catalyses the degradation of sphingomyelin to phosphorycholine and ceramide; ceramide causes membrane changes mediating viral uptake (1). ASM has shown to be activated by SARS-CoV-2. FIASMA’s have been demonstrated to displace ASM from lysosomal membranes, causing partial degradation, reducing the viral membrane fusion with the host cell (2).

A double blind randomised controlled trial of 152 participants used a fluvoxamine dose of 100MG TDS for 15 days for SARS-CoV-2 non-hospitalised patients. Of the 80 participants that received fluvoxamine, none of them died, compared to 6 in the control group (3). The TOGETHER trial used a dose of 100MG BD for 10 days. 741 received fluvoxamine and 756 received the placebo. There were 17 deaths in the fluvoxamine group compared to 25 in the placebo, p=0.24. Hospitalisation was reduced with fluvoxamine (119 [16%] of 756 vs 79 [11%] of 741); relative risk [RR] 0·68; 95% Bayesian credible interval [95% BCI]: 0·52–0·88). However, the endpoint defined staying in the emergency department for > 6 hours as hospitalisation, when this was removed the difference was not significant (4).

A quasi-randomised trial was performed in California after COVID-19 outbreak. During mass testing, patients were given the choice of fluvoxamine therapy. 50-100 MG loading dose, then 50MG BD for 14 days, follow up at 7 and 14 days. The fluvoxamine group were 5% non-white compared to 27% no therapy group. 38% of fluvoxamine patients were asymptomatic at day 0 compared to 58% in no therapy. Hospitalisation up to day 14 was 0/65 (0%) for fluvoxamine and 6/48 (12.5%) for no therapy (P=0.005) with 2 needing mechanical ventilation and one dying. At day 14 - ongoing symptoms were present in 0% with fluvoxamine compared with 60% (29 of 48) with no therapy (P < 0.001). The most common ongoing symptom were anxiety (n = 19), however it was not shown which group was more anxious. No serious adverse events occurred with fluvoxamine (5).

Dose

In this study, patients will be given 100mg TDS starting on D0 and for a further 6 days, as tolerated.

Traditionally treatment is initiated at the lowest possible dose (50 mg) given once daily at bedtime, and then increased to 100 mg daily at bedtime after a few days, as tolerated.

Effective daily dose is usually between 100 mg and 200 mg, up to a maximum of 300 mg.

In the clinical trials listed above dosing was 100mg TDS, 100mg BD and 50mg BD (with loading dose) respectively – a large range.

Pharmacokinetic characteristics

Fluvoxamine is well absorbed after oral administration. Following a single 100 mg oral dose, peak plasma levels of 31-87 ng/mL were attained 1.5 to 8 hours post-dose. Peak plasma levels are directly proportionate to dose of single oral doses.

Following single doses, the mean plasma half-life is 15 hours, and slightly longer (17-22 hours), during repeated dosing. Steady-state plasma levels are usually achieved within 10-14 days.

Fluvoxamine undergoes extensive hepatic transformation, to at least nine metabolites, which are excreted by the kidney.

Pharmacokinetic sampling schedule: (only if there is evidence of accelerated viral clearance)

The timings are:

- Pre-dose D0H0, then
- 1, 2, 3, 4, 6, 9, 12 and 24h post dose

Note that blood will be taken on D3 and D7 for routine haematology and biochemistry. The EDTA sample will be used to measure the drug concentrations.

Adverse effects

Common adverse effects of fluvoxamine are gastrointestinal complaints (nausea, constipation, anorexia, diarrhoea and dyspepsia) and central nervous system complaints (somnolence, dry mouth, nervousness, insomnia and agitation). Care must be taken in those with existing mental health problems. Serotonin syndrome can be prevented by avoiding overuse.

Contraindications

Known hypersensitivity

Use of MAOIs and thioridazine – fluvoxamine must not be given within 14 days of discontinuation, and a 5-week gap must be given before starting an MAOI or thioridazine

Other antidepressants (e.g. Selective Serotonin Reuptake Inhibitors (SSRIs), which increase the risk of serotonin syndrome.

Cautions

Patients with hepatic or renal insufficiency should begin treatment with a low dose and becarefully monitored.

Caution should be taken with those with a pre-existing psychiatric condition

References

1.	Kornhuber J, Hoertel N, Gulbins E. The acid sphingomyelinase/ceramide system in COVID-19. Molecular Psychiatry. 2021:1-8.

2.	Carpinteiro A, Edwards MJ, Hoffmann M, Kochs G, Gripp B, Weigang S, et al. Pharmacological inhibition of acid sphingomyelinase prevents uptake of SARS-CoV-2 by epithelial cells. Cell Reports Medicine. 2020;1(8):100142.

3.	Lenze EJ, Mattar C, Zorumski CF, Stevens A, Schweiger J, Nicol GE, et al. Fluvoxamine vs placebo and clinical deterioration in outpatients with symptomatic COVID-19: a randomized clinical trial. Jama. 2020;324(22):2292-300.

4.	Reis G, dos Santos Moreira-Silva EA, Silva DCM, Thabane L, Milagres AC, Ferreira TS, et al. Effect of early treatment with fluvoxamine on risk of emergency care and hospitalisation among patients with COVID-19: the TOGETHER randomised, platform clinical trial. The Lancet Global Health. 2021.

5.	Seftel D, Boulware DR, editors. Prospective cohort of fluvoxamine for early treatment of coronavirus disease 19. Open Forum Infectious Diseases; 2021: Oxford University Press US.

#### 16.9 AZD7442 (Evusheld)

Rationale

AZD7442 is a combination of two long acting antibodies – tixagevimab and cilgavimab. They were both derived from B cells donated by patients who recovered from SARS-CoV-2. Individually they prevent the spike protein from being attached to the ACE2 receptor, together they extend the half-life and reduce effector functions. In non-human primate models, prophylactic AZD7442 prevented infection from SARS-CoV-2, and therapeutic administration caused accelerated virus clearance from lungs. A phase 1 study in healthy participants found that a 300 MG IM injection (IM injections of 150MG of tixagevimab and 150MG of cilgavimab), provided SARS-CoV-2 neutralising titres 10-fold higher than that of convalescent serum for at least 3 months. They remained 3 fold higher after 9 months (1).

A phase 3 double blind placebo-controlled study (PROVENT trial) used Evusheld as immunoprophylaxis on unvaccinated adults without prior infection. The intervention group was given a single 300 MG AZD7442 dose for prevention of symptomatic positive SARS-CoV-2 RT-PCR. Primary efficacy analysis showed that the relative risk ratio was 0.77 (0.46 – 0.90) vs placebo (P<0.001) (2).

Another Phase 3 randomised double-blind controlled trial (STORM CHASER) gave unvaccinated adults with confirmed exposure to SARS-CoV-2 (within the last 8 days) a single 300 MG AZD7442 dose. AZD7442 reduced the risk of developing symptomatic COVID-19 by 73% (27 – 90%) (3).

The TACKLE phase 3, randomised, double blind, placebo-controlled trial used a 600MG IM dose of AZD7442 (IM injections of 300MG of tixagevimab and 300MG of cilgavimab). Eligible patients were those with mild-to-moderate COVID-19 and symptomatic for no longer than seven days. Primary end point was severe COVID-19 or death. In the intervention arm there were 18 recorded events (18/407) and 37 in the placebo arm (37/415) (4).

Composition & dose

Recommended dose by the US FDA is 300MG of tixagevimab and 300 MG of cilgavimab administered as consecutive IM injections at different sites. Patients should be monitored for at least 1 hour after administration.

Pharmacokinetic characteristics

Peak plasma time is 14 days for both antibodies, peak plasma concentration is 16.5 mcg/mL for tixagevimab and 15.3 mcg/ml for cilgavimab. The half-life is 87.9 days for tixagevimab and 82.9 days for cilgavimab.

Pharmacokinetic schedule: only if there is evidence of accelerated viral clearance

As a type B intervention (positive control), intensive pharmacokinetic sampling will not be conducted on those receiving Evusheld.

Toxicity

Serious hypersensitivity reactions, including anaphylaxis, have been observed with Human immunoglobulin G1 (IgG1) monoclonal antibodies like Evusheld. If signs and symptoms of a clinically significant hypersensitivity reaction or anaphylaxis occur while taking Evusheld, immediately discontinue administration and initiate appropriate medications and/or supportive care.

Contraindications

Hypersensitivity

Cautions

A higher proportion of subjects who received Evusheld versus placebo have reported myocardial infarction and cardiac failure serious adverse events. All of these subjects had cardiac risk factors or a previous history of cardiovascular disease, there was no clear temporal pattern. A causal relationship between Evusheld and these events has not been established.

Drug Interactions

No formal drug interaction studies have been performed with Evusheld. Tixagevimab and cilgavimab are not renally excreted or metabolized by cytochrome P450 (CYP) enzymes. Interactions with concomitant medications that are renally excreted or that are substrates, inducers, or inhibitors of CYP enzymes are unlikely.

References

1.	Loo Y-M, McTamney PM, Arends RH, Abram ME, Aksyuk AA, Diallo S, et al. The SARS-CoV-2 monoclonal antibody combination, AZD7442, is protective in non-human primates and has an extended half-life in humans. Science Translational Medicine. 2022:eabl8124.

2.	Levin MJ, Ustianowski A, De Wit S, Launay O, Avila M, Seegobin S, et al., editors. LB5. PROVENT: Phase 3 Study of Efficacy and Safety of AZD7442 (Tixagevimab/Cilgavimab) for Pre-exposure Prophylaxis of COVID-19 in Adults. Open Forum Infectious Diseases; 2021: Oxford University Press US.

3.	CHASER S. AstraZeneca: Update on AZD7442 STORM CHASER trial in post-exposure prevention of symptomatic COVID-19.

4.	Mode D, Stockholm LC. AstraZeneca: AZD7442 reduced risk of developing severe COVID-19 or death in TACKLE Phase III outpatient treatment trial.

### 17 Appendix 3 – Sample size simulations

Sample size projections were done via simulation. Each simulation used a pharmacodynamic model of viral clearance (4) that was fitted to prospectively collected viral load data in 46 individuals. For simplicity, we assumed there was no site dependent effects.

Interim analyses were performed after 50 patients were enrolled and then for every subsequent 25 patients. In the simulation, out of the 5 interventions 4 were ineffective (no increase in viral clearance) and 1 was effective (10% increase in viral clearance). Figures 6 & 7 show the type 1 and 1-type 2 errors for different choices of stopping rule thresholds, respectively.

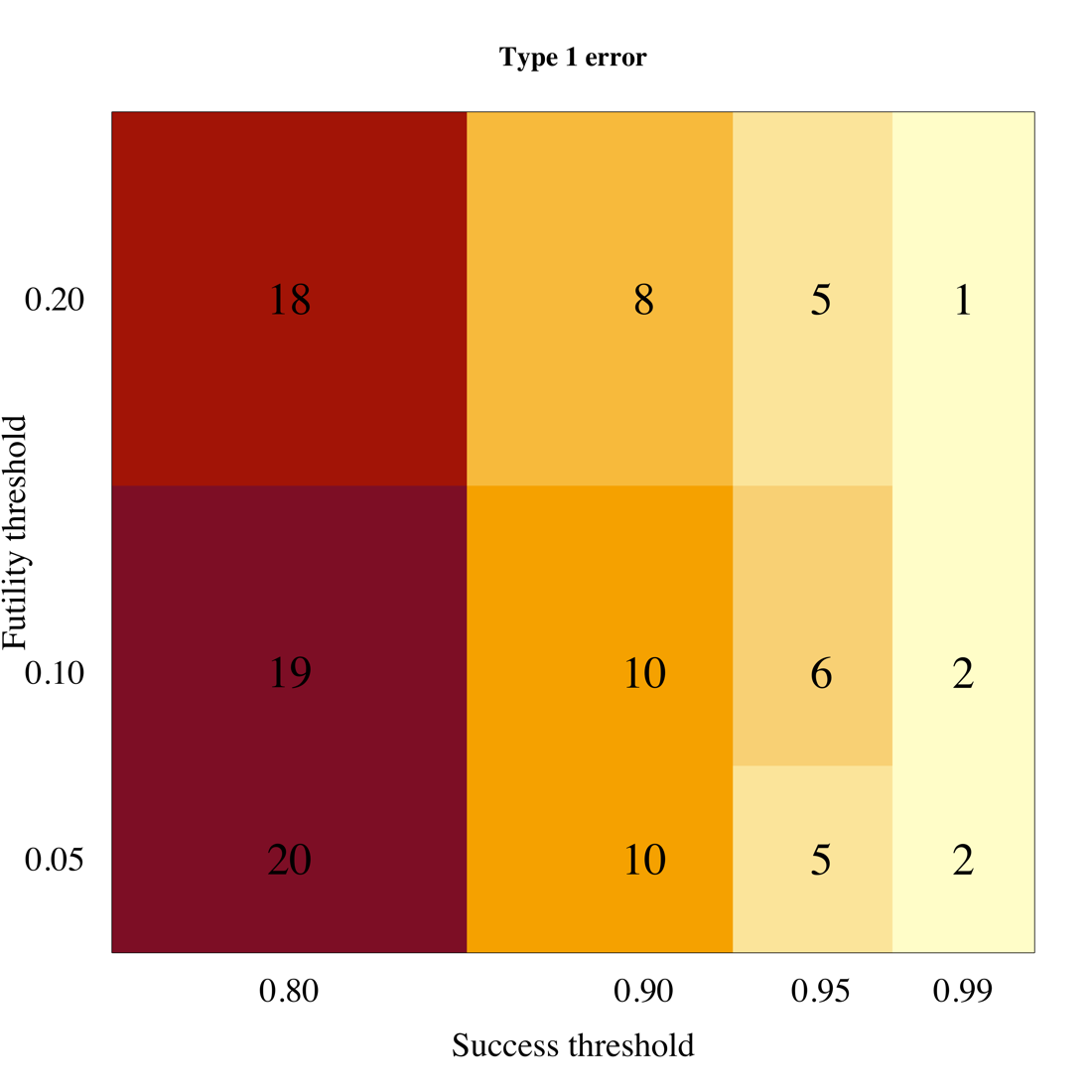

Figure 6 Estimated type 1 error

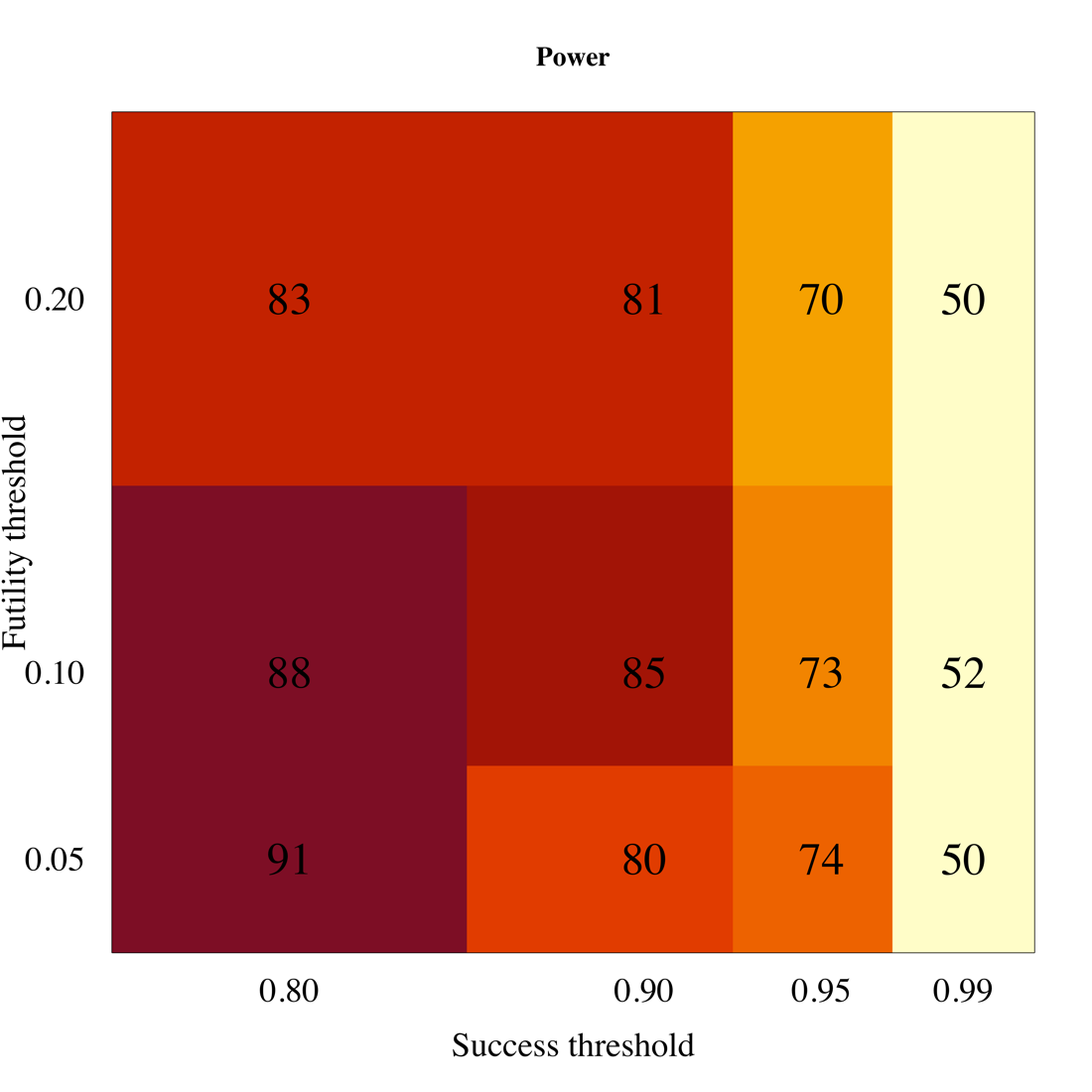

Figure 7 Estimated power (1-type 2 error)

### 18 Appendix 4 – Exclusion criteria

The exclusion criteria in the Master Protocol are expanded upon here. The exclusion criteria in **bold** below have a corresponding section in this appendix where more details are given. The patient may not enter the study if ANY of the following apply:

- **Taking any concomitant medications or drugs**†
- **Presence of any chronic illness/ condition requiring long term treatment, or other significant comorbidity**
- **Laboratory abnormalities discovered at screening**
- For females: pregnancy, actively trying to become pregnant, or lactation
- **Contraindication to taking, or known hypersensitivity reaction to any of the proposed therapeutics**
- Currently participating in another COVID-19 therapeutic or vaccine trial
- Evidence of pneumonia (although imaging is NOT required)

† healthy women on the oral contraceptive pill are eligible to join the study

1. Taking any concomitant medications or drugs

The study will exclude all patients taking **regular drugs** including herbal drugs for the treatment of COVID-19, but some patients may take over the counter drugs or other vitamin and herbal supplements if considered safe in the judgement of the site investigator upon screening assessment. Patients should be counseled to end and avoid all non-essential medications during study participation. In certain circumstances where a patient is on a regular medication, which does not reflect an excluded underlying illness and in the decision of the study PI is not likely to interfere with the study outcomes, or predispose the patient to any increased risk, a decision can be made to enrol. In all cases, the risks/ benefits of this decision should be documented, as well as the person(s) making the decision.

Drug interactions change rapidly as new data become available. If in any doubt, please refer to some of the web sites below:

<https://www.covid19-druginteractions.org/checker>

<https://crediblemeds.org/>

<https://compendium.ch/fr/Patient> - this is a Swiss website dealing with drug interactions

<https://www.bnf.org/products/bnf-online/> - available only in the UK

<https://www.uptodate.com/drug-interactions/?source=responsive_home#di-druglist> – this requires a subscription

2. Presence of any chronic illness/ condition requiring long term treatment, or other significant comorbidity

The protocol excludes almost all individuals with a chronic illness of any severity and those with an illness that requires long term treatment. In addition, the following are considered significant comorbidities due to their known association with worse outcomes in COVID-19 according to the MOPH guideline; 1. Chronic respiratory disease, 2. Cardiovascular disease, 3. Chronic kidney disease, 4. Stroke, 5. Obesity, 6. Cancer, 7. Diabetes.

BMI ≥ 35 kg/m^2^

Diet controlled diabetes mellitus

Reduced kidney function – eGFR < 70 mls/min/1.73m^2^*

Any known underlying liver disease, HIV infection and other immunocompromised condition.

Asplenia

Any underlying bleeding disorder e.g. haemophilia, von Willibrand’s disease

Suffering with any other disease which in the opinion of the investigator poses undue risk to the potential patient

*eGFR for males is calculated based on the Chronic Kidney Disease Epidemiology Collaboration (Andrew S Levey, Lesley A Stevens et al. 2009)

<https://qxmd.com/calculate/calculator_251/egfr-using-ckd-epi>

3. Laboratory abnormalities discovered at screening

These are:

haemoglobin < 8 g/dL

platelet count < 50,000/uL

ALT > x 2 ULN

total bilirubin > 1.5 x ULN

eGFR < 70 mls/min/1.73m^2^

4. Contraindication to taking, or known hypersensitivity reaction to any of the proposed therapeutics:

Where a contraindication is mentioned in the above sections, or an above contraindication incorporates drug-specific contraindications below, it is not repeated again in this section, unless further information is given which warrants some repetition. For a complete list of drug-specific contraindications please see the section for that drug in Appendix 2.

To repeat, a known hypersensitivity reaction to ANY of the proposed therapeutics below (even if the patient were to be randomised to a different treatment arm or control) is an exclusion criterion from entering into the study.

The following are relevant contraindications (i.e. in potentially eligible patients):

##### Hydroxychloroquine

patients with pre-existing retinopathy of the eye

patients with known hypersensitivity to any 4-aminoquinoline compounds

epilepsy

Prolonged QTc (see Hydroxychloroquine specific appendix)

##### Remdesivir

No additional contraindications

##### Ivermectin

No additional contraindications

##### Nitazoxanide

No additional contraindications

##### Miglustat

For females of childbearing age, contraception should be used throughout the study and for 2 weeks post last dose

##### Lopinavir/ritonavir

Concomitant use with drugs that are metabolised by CYP 3A

##### REGN-COV2

No additional contraindications

##### Nebulised Unfactionated Heparin (UFH)

- Heparin allergy or Heparin-induced thrombocytopaenia
- Known prolonged APTT > 120ms or bleeding diathesis
- Pulmonary bleeding or uncontrolled bleeding

##### Favipiravir

- Gout or history of gout
- For females of childbearing age, they should use effective contraception during the study and for 7 days after the last dose. For males, contraception should be used during the study and for 7 days post last dose and they should abstain from sex with a pregnant women.

##### Molnupiravir

As per USA FDA advice, those on molnupiravir need to be aware of avoiding pregnancy.  Females of child bearing potential need to use a reliable method of contraception for the duration of the treatment, and also for 4 days after the final dose of molnupiravir.  Males of reproductive potential, if they are sexually active with females of child bearing potential, should use a reliable form of contraception during the treatment and at least 3 months after the final dose

##### Nirmatrelvir/ritonavir

No additional contraindications

##### Sotrovimab

No additional contraindications

##### Fluoxetine

No additional contraindications

##### Fluvoxamine

No additional contraindications

##### AZD7442(Evusheld)

No additional contraindications

##### References for exclusion criteria

Andrew S Levey, Lesley A Stevens, Christopher H Schmid, Yaping Lucy Zhang, Alejandro F Castro, Harold I Feldman, John W Kusek, Paul Eggers, Frederick Van Lente, Tom Greene and J. Coresh (2009). "A new equation to estimate glomerular filtration rate." Annals of Internal Medicine **150**: 604-612.

Williamson, E. J., A. J. Walker, K. Bhaskaran, S. Bacon, C. Bates, C. E. Morton, H. J. Curtis, A. Mehrkar, D. Evans, P. Inglesby, J. Cockburn, H. I. McDonald, B. MacKenna, L. Tomlinson, I. J. Douglas, C. T. Rentsch, R. Mathur, A. Y. S. Wong, R. Grieve, D. Harrison, H. Forbes, A. Schultze, R. Croker, J. Parry, F. Hester, S. Harper, R. Perera, S. J. W. Evans, L. Smeeth and B. Goldacre (2020). "Factors associated with COVID-19-related death using OpenSAFELY." Nature **584**(7821): 430-436.
