## Supplementary materials for "Pharmacometric assessment of the *in vivo* antiviral activity of ivermectin in early symptomatic COVID-19"

**Table of Contents:**

1: List of sites and investigators (PLATCOV Collaborative Group)

2: Acknowledgements

3: Adverse Events

4: Serious Adverse Events

5: Ivermectin visual side effects

6: Virus variant determination

7: Ivermectin dosing table

8: Randomization

9: Statistical analysis

10: Supplementary Figures

11: References

**List of Sites and Investigators (PLATCOV Collaborative Group)**

**Sites**

1. Hospital for Tropical Diseases (HTD), Faculty of Tropical Medicine, Mahidol University, 420/6 Rajvithi Road, Bangkok, 10400, Thailand
2. Vajira Hospital (VJ), Navamindradhiraj University, 681 Samsen road, Dusit, Bangkok, 10300, Thailand
3. Bangplee Hospital (BP), 88/1 Moo 8 Tambon Bang Phli Yai, Amphoe Bangplee, Samut Prakan 10540, Thailand

**Investigators**

Co-principal investigators:

Nicholas J White^1,2^

William HK Schilling^1,2^

Faculty of Tropical Medicine:

Site and Country Principal investigator:

Weerapong Phumratanaprapin^4^

Accountable Investigator:

Viravarn Luvira^4^

Co-Investigators:

James J Callery^1,2^

Nicholas PJ Day^1,2^

Sasithon Pukrittayakamee^1,4^

Simon Boyd^1,2^

Cintia Cruz^1,2^

Arjen M Dondorp^1,2^

Walter RJ Taylor^1,2^

James A Watson^1,2^

Watcharapong Piyaphanee^4^

Kittiyod Poovorawan^1,4^

Thundon Ngamprasertchai^4^

Tanaya Siripoon^4^

Borimas Hanboonkunupakarn^1,4^

Kesinee Chotivanich^1,4^

Podjanee Jittamala^1,3^

Mallika Imwong^1,5^

Janjira Thaipadungpanit^1,4^

Vajira Hospital:

Site Principal investigator:

Vasin Chotivanich^7^

Co-investigators:

Wiroj Ruksakul^7^

Chunlanee Sangketchon^8^

Bangplee Hospital:

Site Principal investigator:

Pongtorn Hanboonkunupakarn^6^

Co-investigator:

Sakol Sookprome^6^

Universidade Federal de Minas Gerais

Mauro M Teixeira^9^

1. Mahidol Oxford Tropical Medicine Research Unit, Faculty of Tropical Medicine, Mahidol University, Bangkok, Thailand

2. Centre for Tropical Medicine and Global Health, Nuffield Department of Medicine, Oxford University, Oxford, UK

3. Department of Tropical Hygiene, Faculty of Tropical Medicine, Mahidol University, Bangkok, Thailand

4. Department of Clinical Tropical Medicine, Faculty of Tropical Medicine, Mahidol University, Bangkok, Thailand

5. Department of Molecular Tropical Medicine and Genetics, Faculty of Tropical Medicine, Mahidol University, Bangkok, Thailand

6. Bangplee Hospital, Ministry of Public Health, Thailand

7. Faculty of Medicine, Navamindradhiraj University, Bangkok,Thailand

8. Faculty of Science and Health Technology, Navamindradhiraj University, Bangkok,Thailand

9. Department of Biochemistry and Immunology, Universidade Federal de Minas Gerais, Minas Gerais, Brazil

**Acknowledgements**

We thank the data safety and monitoring board (DSMB): Tim Peto, Andre Siqueira, and Panisadee Avirutnan, and the trial steering committee (TSC): Nathalie Strub-Wourgraft, Martin Llewelyn, Deborah Waller, and Attavit Asavisanu. We thank Sompob Saralamba and Tanaphum Wichaita for developing the RShiny randomisation app and Mavuto Mukaka for statistical support. We would also like to thank all the staff at the Clinical Trials Unit (CTU) at MORU. We thank the PCR Expert group (Janjira Thaipadungpanit, Audrey Dubot-Pérès, Clare Ling and Elizabeth Batty), Thermo Fisher for their excellent support with this project, and all the hospital staff at the Hospital of Tropical Diseases (HTD), Bangplee (BP) and Vajira (VJ) hospitals, as well as those involved in sample processing in MORU and the processing and analysis at the Faculty of Tropical Medicine, molecular genetics laboratory. We would also like to thank the MORU Clinical Trials Support Group (CTSG) for data management and logistics, and the purchasing, administration and support staff at MORU. Finally, we would like to thank the patients with COVID-19 who volunteered to be part of the study.

**Adverse events (AE)**

**Table 1: Summary of adverse events (grade 3 and above)**

|  | **All grades** | | | **Grade 3-4** | | |
| --- | --- | --- | --- | --- | --- | --- |
|  | **Ivermectin (n=46)** | **Casirivimab/ imdevimab**  **(n=10)** | **No study drug (n=45)** | **Ivermectin (n=46)** | **Casirivimab/ imdevimab**  **(n=10)** | **No study drug (n=45)** |
| Any adverse event (Grade ≥ 3) | 0 | 0 | 2 |  |  |  |
| Serious adverse event reported | 0 | 0 | 2 |  |  |  |
| Symptoms |  |  |  |  |  |  |
| Fever |  |  |  | 0 | 0 | 0 |
| Headache |  |  |  | 0 | 0 | 0 |
| Dizziness |  |  |  | 0 | 0 | 0 |
| Blurred vision |  |  |  | 0 | 0 | 0 |
| Fatigue |  |  |  | 0 | 0 | 0 |
| Cough |  |  |  | 0 | 0 | 0 |
| Difficulty breathing |  |  |  | 0 | 0 | 0 |
| Chest pain |  |  |  | 0 | 0 | 0 |
| Running nose |  |  |  | 0 | 0 | 0 |
| Loss of smell or taste |  |  |  | 0 | 0 | 0 |
| Abdominal pain |  |  |  | 0 | 0 | 0 |
| Loss of appetite |  |  |  | 0 | 0 | 0 |
| Nausea |  |  |  | 0 | 0 | 0 |
| Vomiting |  |  |  | 0 | 0 | 0 |
| Diarrhoea |  |  |  | 0 | 0 | 0 |
| Arthralgia |  |  |  | 0 | 0 | 0 |
| Myalgia |  |  |  | 0 | 0 | 0 |
| Itching |  |  |  | 0 | 0 | 0 |
| Skin rash |  |  |  | 0 | 0 | 0 |
| Laboratory abnormalites |  |  |  |  |  |  |
| Creatinine |  |  |  | 0 | 0 | 0 |
| BUN |  |  |  | 0 | 0 | 0 |
| Sodium |  |  |  | 0 | 0 | 0 |
| eGFR |  |  |  | 0 | 0 | 0 |
| Potassium |  |  |  | 0 | 0 | 0 |
| ALT/SGPT |  |  |  | 0 | 0 | 0 |
| AST/SGOT |  |  |  | 0 | 0 | 0 |
| Total bilirubin |  |  |  | 0 | 0 | 0 |
| Direct bilirubin |  |  |  | 0 | 0 | 0 |
| Alkaline Phosphatase |  |  |  | 0 | 0 | 0 |
| LDH |  |  |  | 0 | 0 | 0 |
| Creatinine phosphokinase (CPK) |  |  |  | 0 | 0 | 2* |
| Anemia |  |  |  | 0 | 0 | 0 |
| Leukocytopenia |  |  |  | 0 | 0 | 0 |
| Neutropenia |  |  |  | 0 | 0 | 0 |
| Thrombocytopenia |  |  |  | 0 | 0 | 0 |

*Both patients were also classified as serious adverse events and are detailed in the serious adverse events table (Table: 2). All AEs solicited were reported between days 0-7 and at day 28.

**Serious Adverse Events**

**Table 2: Summary of Serious Adverse Events**

| **Number** | **Study arm** | **Final diagnosis** | **Relationship to trial drug** | **Resolved** |
| --- | --- | --- | --- | --- |
| 1 | No study drug | Reduction in activities of daily living after COVID-19 infection ^1^ | Not related | Yes |
| 2 | No study drug | COVID-induced rhabdomyolysis ^2^ | Not related | Yes |
| 3 | No study drug | COVID-induced rhabdomyolysis ^2^ | Not related | Yes |

^1^Participant was impeded in activities of daily living one day post-discharge from the ward (day 8) and was readmitted for further investigation complaining of right-sided chest pain and lethargy. Clinical observations incl. oxygen saturations, physical examination and electrocardiogram were unremarkable, laboratory investigations including inflammatory markers and D-Dimers were in the normal range. A SARS-CoV-2 PCR was negative. CT pulmonary angiogram showed no radiological evidence of pulmonary embolus,or pneumonitis. The patient’s symptoms quickly resolved and the patient was discharged the following day.

^2^Both participants had an acute rise in creatinine phosphokinase (CPK) during admission. Myoglobin was not detected on urinalysis. A diagnosis of COVID-induced rhabdomyolysis was made by the clinical team based on symptoms of myalgia and a raised CPK without another identifiable aetiology. Nephrology review advised supportive treatment only. Symptoms resolved, the CPK normalized and was within normal limits on follow-up.

**Ivermectin Visual Side Effects**

Six patients receiving ivermectin complained of visual disurbances although none of these met the pre-defined AE criteria of ≥3. Ivermectin at high doses is known to cause transient visual side effects. (1, 2). Six patients in the trial experienced transient visual symptoms. These were all reviewed by a qualified medical physician, in discussion with the PLATCOV Safety Team and the DSMB and were not considered to be safety concerns to the participants. As these side effects have been previously well documented (1, 2), transient and cause no lasting damage, randomisation into the ivermectin arm was not halted for safety reasons (although individual patients did switch to alternative treatments). Of note, two other participants who did not receive ivermectin also experienced similar transient visual changes. These occurred in a participant who received no study drug and another participant who received remdesivir. These cases were discussed with the DSMB committee who were in agreement with the study team’s decision.

| Case | History |
| --- | --- |
| 1 | On discharge, the participant reported they had been having episodes of unilateral dark grey/black shadowing of the lower half of the right eye’s visual field after four doses of ivermectin. Episodes lasted five seconds and only happened once a day, several hours after receiving ivermectin. They were reviewed by an ophthalmologist whose examination was normal. Symptoms resolved following cessation of ivermectin. |
| 2 | The participant experienced visual changes in both eyes after three doses of ivermectin. There was sudden peripheral blurring/fogging lasting two seconds which self-resolved. Later, widespread black dots developed bilaterally, again lasting two seconds and again self-resolved. Bedside examination was normal. Ivermectin was stopped at the patient’s request. |
| 3 | The participant developed a headache, then later blurred vision bilaterally after one dose of ivermectin. Three hours after dosing there was a bilateral throbbing pain in of the head (no aura) which was relieved with paracetamol and sleep. The next morning (18 hours post-administration), there was bilateral fogging of both eyes. Symptoms lasted 30 seconds to one minute and self-resolved. There was no pain, floaters or flashing, and it was unrelated to position. Examination was normal. Ivermectin was discontinued and there were no further issues. |
| 4 | The participant informed the clinical staff on day five that they had been experiencing daily episodes of blurred vision (appearance of clouding) since being started on ivermectin. This was initially in the left eye but then later both eyes. The episodes lasted about 10 minutes. They were worse on lying down and the timing was related to the administration of ivermectin. They received five doses of ivermectin in total. Eye examination was normal. Episodes spontaneously resolved following cessation of ivermectin . |
| 5 | The participant informed the ward staff on day five that they have been having daily transient visual changes since being given ivermectin (symptoms similar to case 4, both participants were on the study ward together). Symptoms consisted of short episodes of visual blurring in both eyes with some associated dizziness which improved without treatment. |
| 6 | The participant informed the study team at the day 28 follow up that they had been experiencing intermittent blurring of vision from day 3 of the study. Symptoms had subsequently begun to resolve on discharge (day 7). The participant was offered an ophthalmology appointment but declined further follow-up. |

**Virus variant determination**

Viral genetic variants were identified using real-time PCR genotyping with the TaqMan™ SARS-CoV-2 Mutation Panel. The variants circulating in Thailand from 30^th^ September 2021 to 18^th^ April 2022 were Delta (B.1.617.2), and the Omicron BA.1 (B.1.1.529) and Omicron BA.2 subvariants (B.1.1.529). All samples were tested for four canonical mutations of the circulating variants. Those with mutation S.T19R.ACA.AGA were designated Delta (B.1.617.2) if no other mutations in the panel were identified. Those with mutation S.Q493R.CAA.CGA were designated as Omicron BA.1 (B.1.1.529) if no other mutations in the panel were identified. Those with mutations S.Q493R.CA.A.CGA, S.T376A.ACT.GCT and S.V213G.GTG.GGG were designated Omicron BA.2 (B.1.1.529) if S.T19R.ACA.AGA was absent.

**Ivermectin dosing table**

The dose in this study was **600 micrograms/kg body weight given once daily for 7 days** (taken with food).

| **Weight in kg** | **Number of 6 mg tablets** | **Dose in mg** | **mg/kg** |
| --- | --- | --- | --- |
| 40 - <50 | 4 | 24 | 0.49 - 0.6 |
| 50 - <60 | 5 | 30 | 0.51 - 0.6 |
| 60 - <70 | 6 | 36 | 0.52 - 0.6 |
| 70 - <80 | 7 | 42 | 0.53 - 0.6 |
| 80 - <90 | 8 | 48 | 0.54 - 0.6 |
| 90 - <100 | 9 | 54 | 0.55 - 0.6 |
| ≥ 100 | 10 | 60 | ≤ 0.6 |

**Randomization**

The randomization sheets were generated by the trial statistician (James Watson).

All new randomization sheets and all updates of existing randomization sheets were done using a pre-written R script which was stored on the randomization Dropbox folder (owner is MORU, under custodianship of the head of MORU IT; this file is a full ‘Professional’ version with history recorded and only the trial statistician and head of IT had access. The file took the following inputs:

- Site codes (e.g. “th001”) for which to generate randomization sheets;
- The set of arms available for randomization in that site;
- The number of arm repeats per block (this is set to the minimum integer such that in each block there is an integer number for each arm);
- The randomization data file from each site (which has the patient numbers for subjects already randomized) named data-XXX.csv (where XXX is the site code), if this does not yet exist a blank csv (headers only) is generated.

This R script is run every time a new site becomes active and every time the set of available arms changes. The output is a csv file named rand-XXX.csv (where XXX is the site code). This overwrites the pre-existing file (which can be retrieved from the Dropbox version history). Each time the randomization script is run, this is recorded on a log file*.*

The randomization is done according to the following constraints:

- Blocks of 2*number of available arms;
- Additional ‘fuzziness’ by swapping one patient allocation per block at random (this can be swapped for any of the available arms) – this avoids knowing which arm the last patient per block will receive.

Each time an authorized member of the study team logs onto the web-app this is logged (timestamp and username).

Each time a new patient is randomized this is logged on to the file data-XXX.csv (where XXX is the site code) with the following information:

- Subject number
- Screening number
- Age
- Sex
- Member of study team username
- Timestamp

At the start of the trial (30^th^ September 2021), randomization to casirivimab/imdevimab was set at 10% (positive control), and the other arms had equal uniform randomization ratios of 22.5%. This was changed on the 30^th^ October 2021 so that all arms had equal randomization ratios. This explains why there are slightly fewer than expected patients randomized to casirivimab/imdevimab.

**Statistical Analysis**

The primary analysis consists of fitting Bayesian hierarchical (mixed effects) linear models to the serial log_10_ viral load data up until day 7 (the day 14 data were not used). All models encode residual error as a *t*-distribution with degrees of freedom estimated from the data. The *t*-distribution was chosen for robustness as the residual error is clearly non-Gaussian. The *t*-distribution error model also makes the model robust against model mis-specification (particularly for the linear models) (3). All models include correlated individual random effect terms for both the intercept (baseline viral load) and the slope. All changes to the slope are defined as multiplicative changes on the log scale (a value of 0 equals no change).

The treatment effect is defined as the proportional change (expressed as a multiplicative term) in the population slope of the daily change in log_10_ viral load. The data are modelled on the log_10_ copies per mL scale, after conversion from Ct values using the standard curve generated from the 12 control concentrations (synthetic samples with known viral densities) from each 96 well plate. The standard curve transformation is done by fitting a linear mixed effects model (random slope and random intercept for each plate) to the control data: regressing the Ct values on the known log viral densities. This borrows information across plates and allows for batch effects.

For all models, we adjusted the intercept and slope for the enrolling site (3 sites in total, the reference site is the Hospital of Tropical Diseases which recruited >90% of patients) and for the variant called (Delta is reference: BA.1 and BA.2 are the two alternatives). A subset of models also adjusted the slopes and intercepts for:

- Age
- Number of vaccine doses
- Result of serology antibody test
- Days since symptom onset

All models except model 1 adjust for human RNase P (proxy for the number of human cells in the sample).

In total we fit 9 separate models:

1. Model 1 is linear with no RNase P adjustment; adjustment for site & variant; weakly informative priors (WIP)
2. **Model 2 is linear with RNase P adjustment; adjustment for site & variant; WIP. This is the main model used to report treatment effects.**
3. Model 3 is non-linear; RNase P adjustment; adjustment for site & variant; WIP
4. Model 4 is linear with RNase P adjustment; adjustment for site & variant; non-informative priors (NIP)
5. Model 5 is non-linear with RNase P adjustment; adjustment for site & variant; NIP
6. Model 6 is linear with RNase P adjustment; full covariate adjustment; WIP
7. Model 7 is non-linear with RNase P adjustment; full covariate adjustment; WIP
8. Model 8 is linear with RNase P adjustment; full covariate adjustment; NIP
9. Model 9 is non-linear with RNase P adjustment; full covariate adjustment; NIP

Model 1 is a base model without RNase P adjustment; models 2-9 all have RNase P adjustment and are all combinations of linear & non-linear models, with or without full covariate adjustment; and with either weakly informative priors or non-informative priors.

We compared model fits using the *loo* (approximate leave-one-out cross validation) package.

The statistical analysis plan provides a detailed overview of the model structures. Comparison of treatment effect under all 9 models is given in Figure S5.

All data, models and analytical output are on the linked GitHub repository: <https://github.com/jwatowatson/PLATCOV-Ivermectin>

This includes all data used in the analysis for full reproducibility of the results.

**Supplementary Figures**

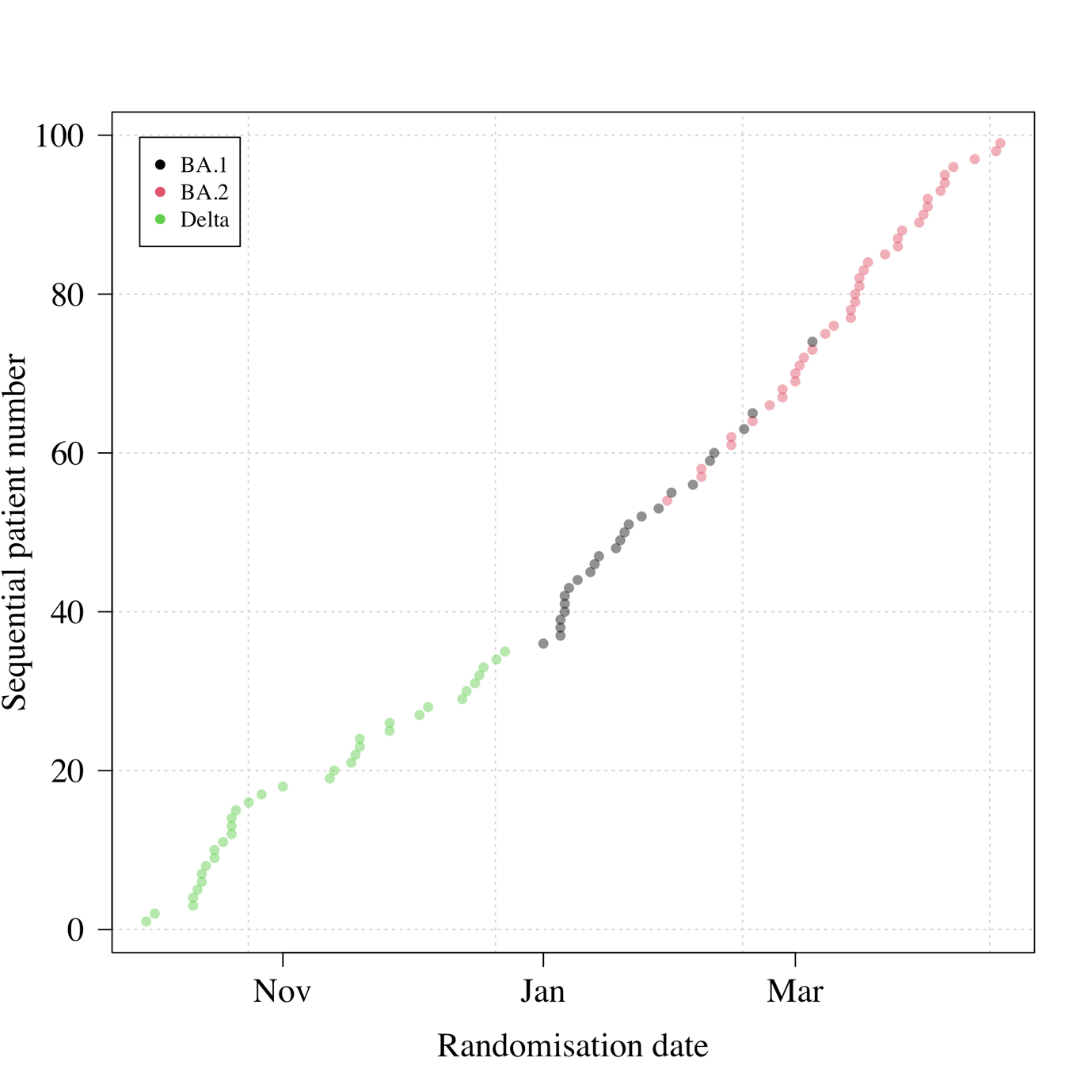

SFigure 1: Randomization dates and virus variants of all patients with available PCR data (n=99). This excludes two patients: one who had no detectable virus (likely an enrolment error); and one who left the study following randomization (no swabs were taken). All patients included in the analysis had their virus genotyped (none were imputed).

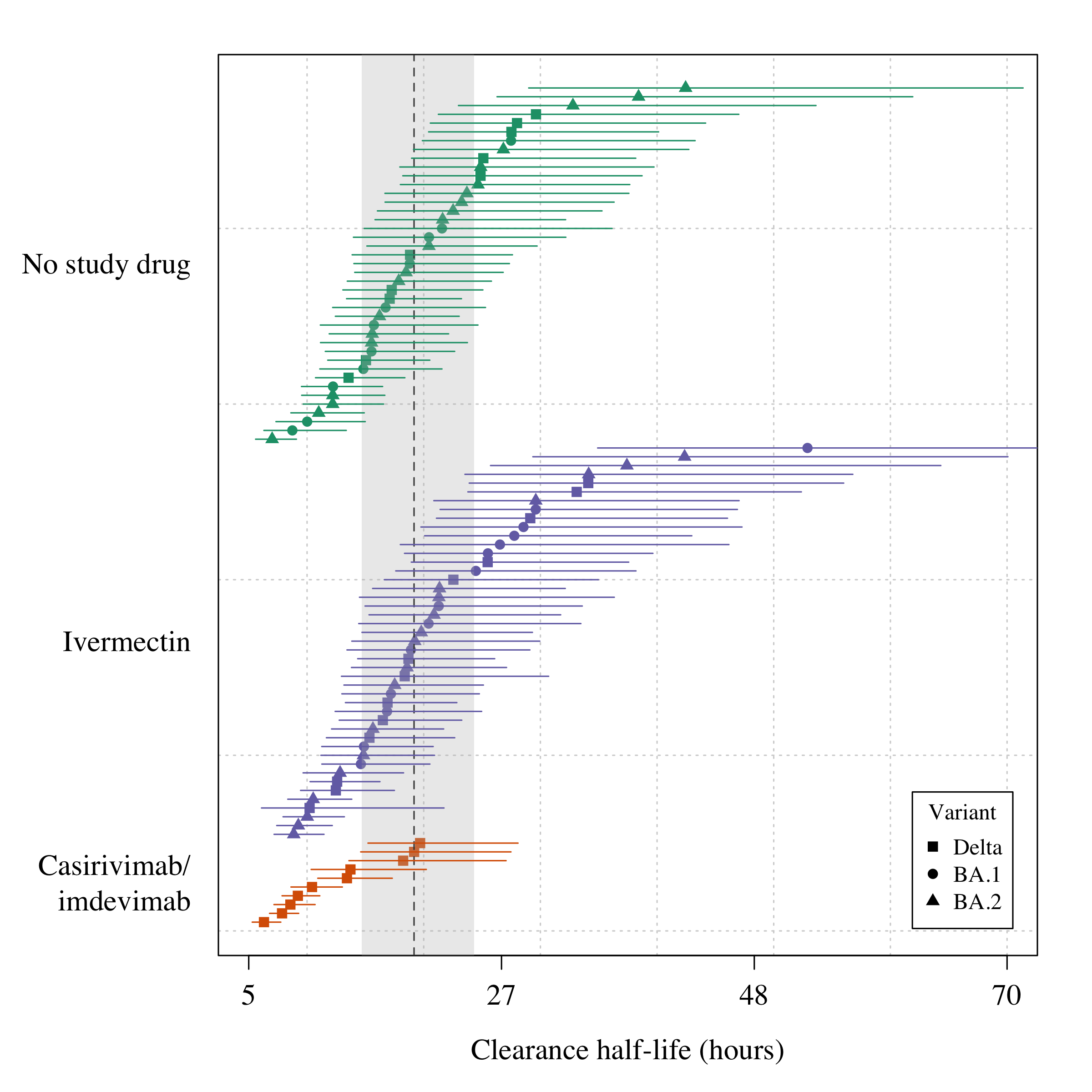

SFigure 2: Estimated clearance half-lives in all patients in the mITT population (n=96), grouped by treatment arm and in order of decreasing rate of clearance. The mean posterior estimated half-lives are shown by the squares/circles/triangles (corresponding to the Delta, BA.1, and BA.2 variants, respectively). The vertical dashed line shows the posterior mean population clearance half-life (the grey shaded area is the 95% credible interval).

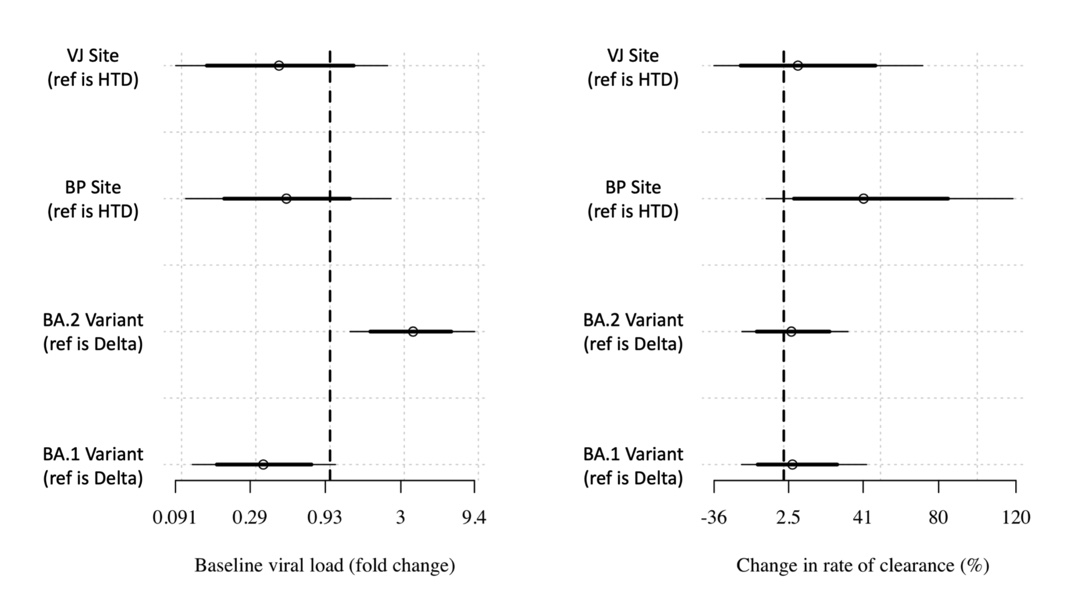

SFigure 3: Covariate effects estimated for the main analytical linear model.

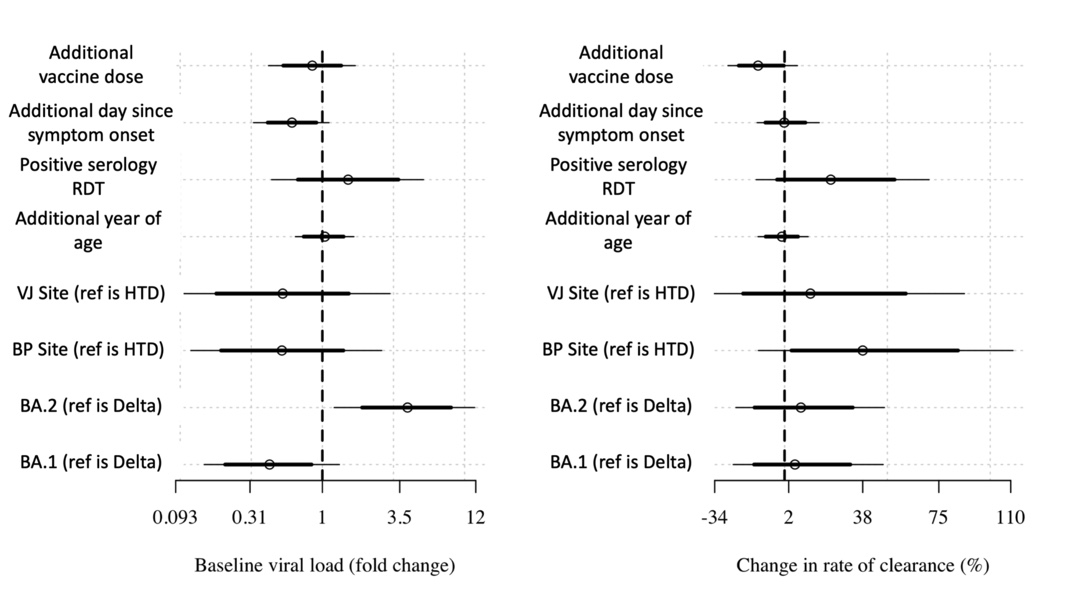

SFigure 4: Covariate effects on intercept (left) and slope (right) for the linear model with additional covariate adjustment.

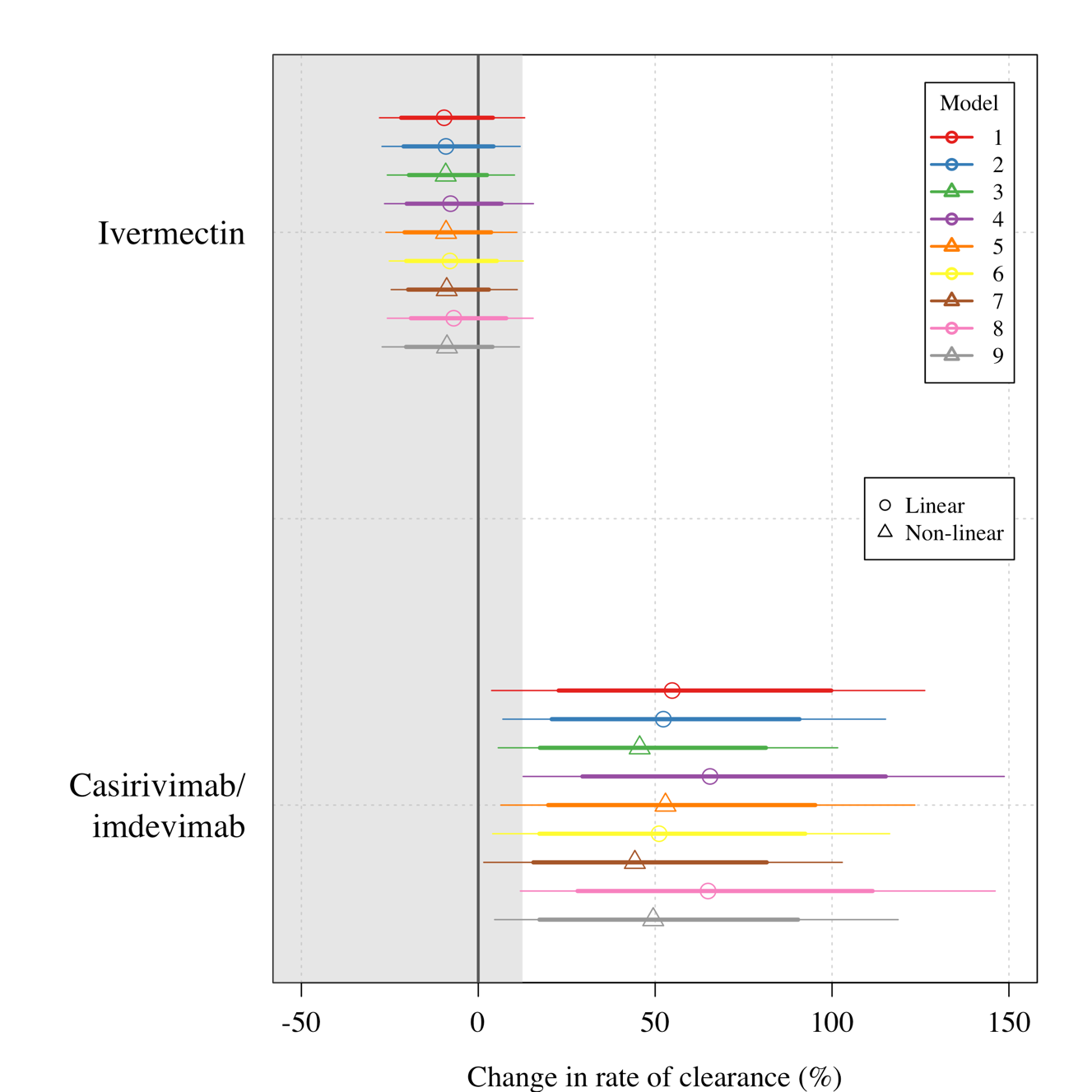

SFigure 5: Treatment effect estimates for all nine models fit to the data. Circles/triangles show the mean posterior estimates (circles: linear models; triangles: non-linear models); thick lines: 80% credible intervals; thin lines: 95% credible intervals. A description of each model is given in the statistical analysis section above.

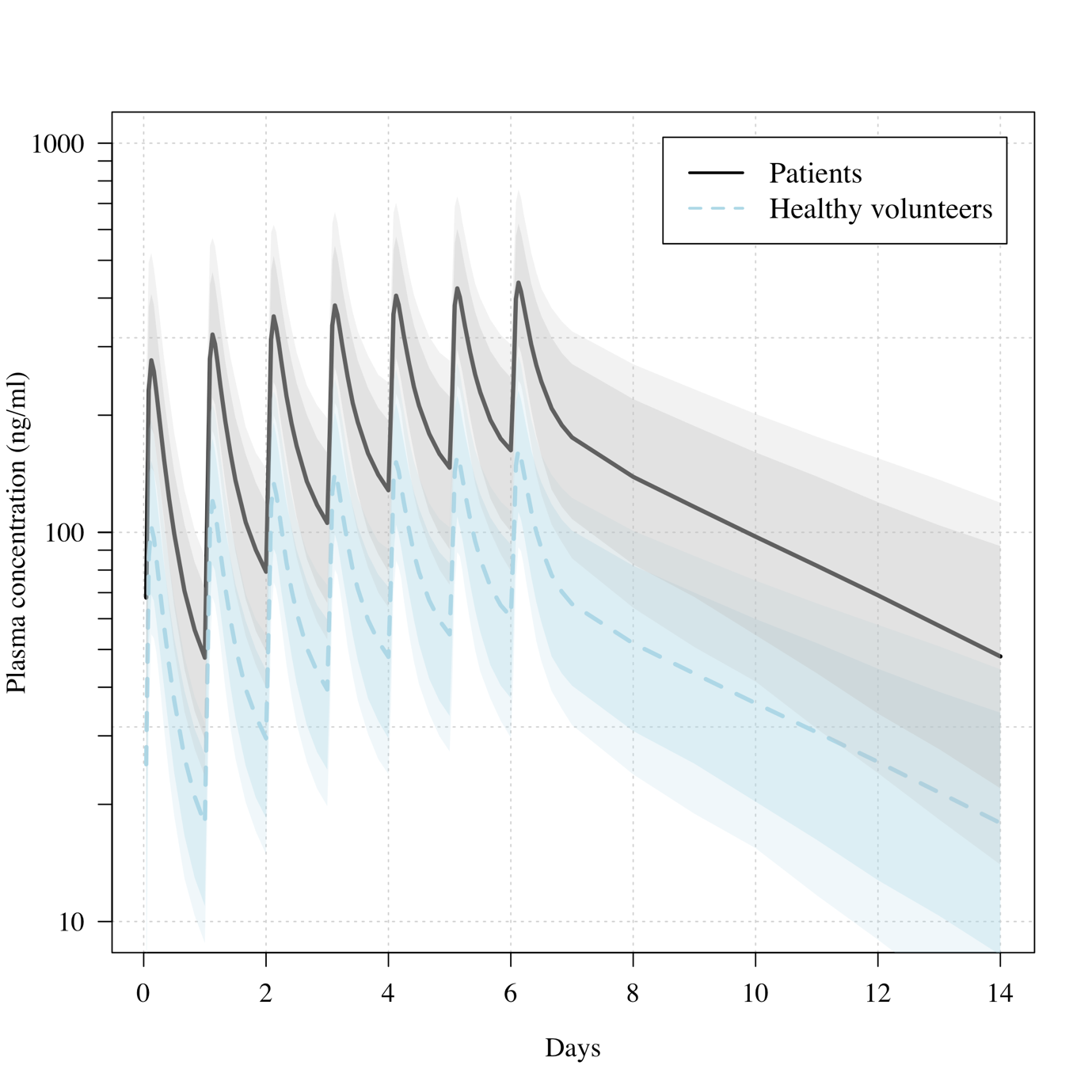

SFigure 6: Predicted ivermectin plasma concentrations over time under the population PK model fit to data from healthy volunteers (4) and the ivermectin patients in the PLATCOV study. Mean predicted concentrations with 80 and 95% confidence intervals are shown for daily dosing of 600μg/kg ivermectin in a 70kg adult over one week for patients (thick line) and healthy volunteers (dashed line). The mean relative bioavailability in patients compared to healthy volunteers was estimated as 2.6.

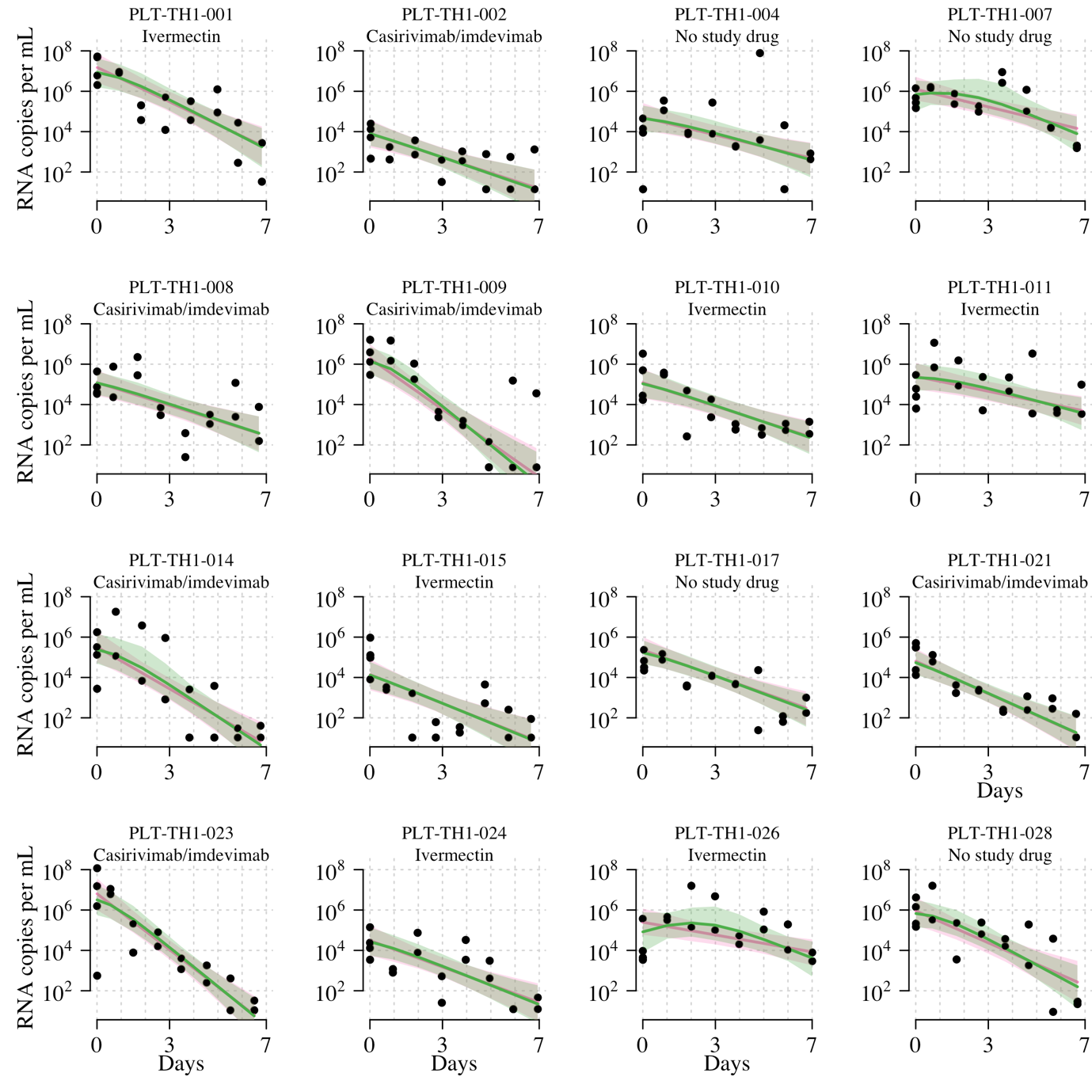

SFigure 7: Individual fits under the two main Bayesian hierarchical models (pink: linear model with RNase P adjustment; green: non-linear model with RNase P adjustment). Lines show mean fits; shaded areas show 95% credible intervals around fits. Black circles show viral load measurements for each independent swab. Only follow-up data included in the mITT analysis dataset are shown (e.g. for patients who switched medication after day 2, we only show data up until the switch i.e. the data that were used for the analysis). Site codes: TH1=HTD, TH57=BP, TH58=VJ.

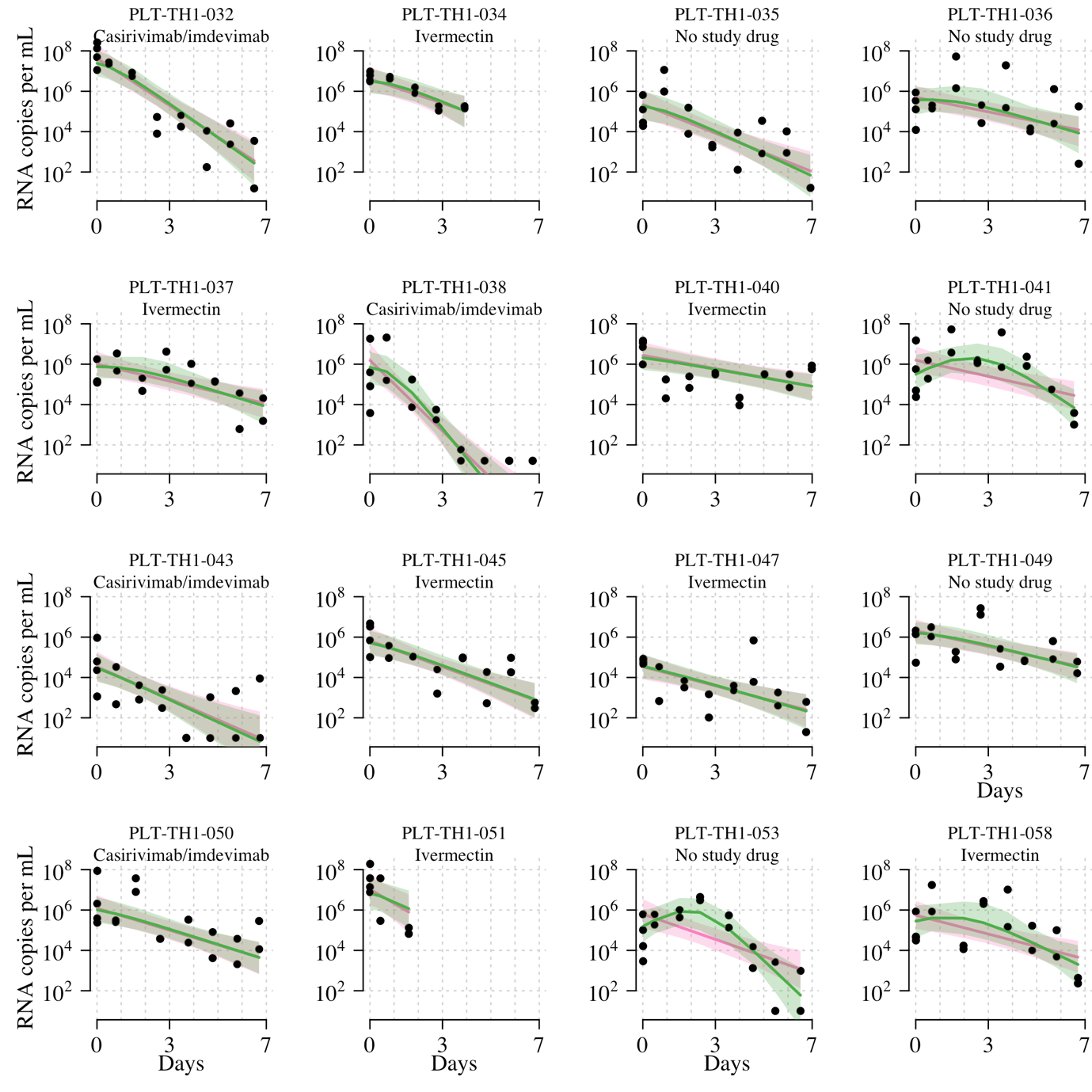
 *SFigure 7 cont.*

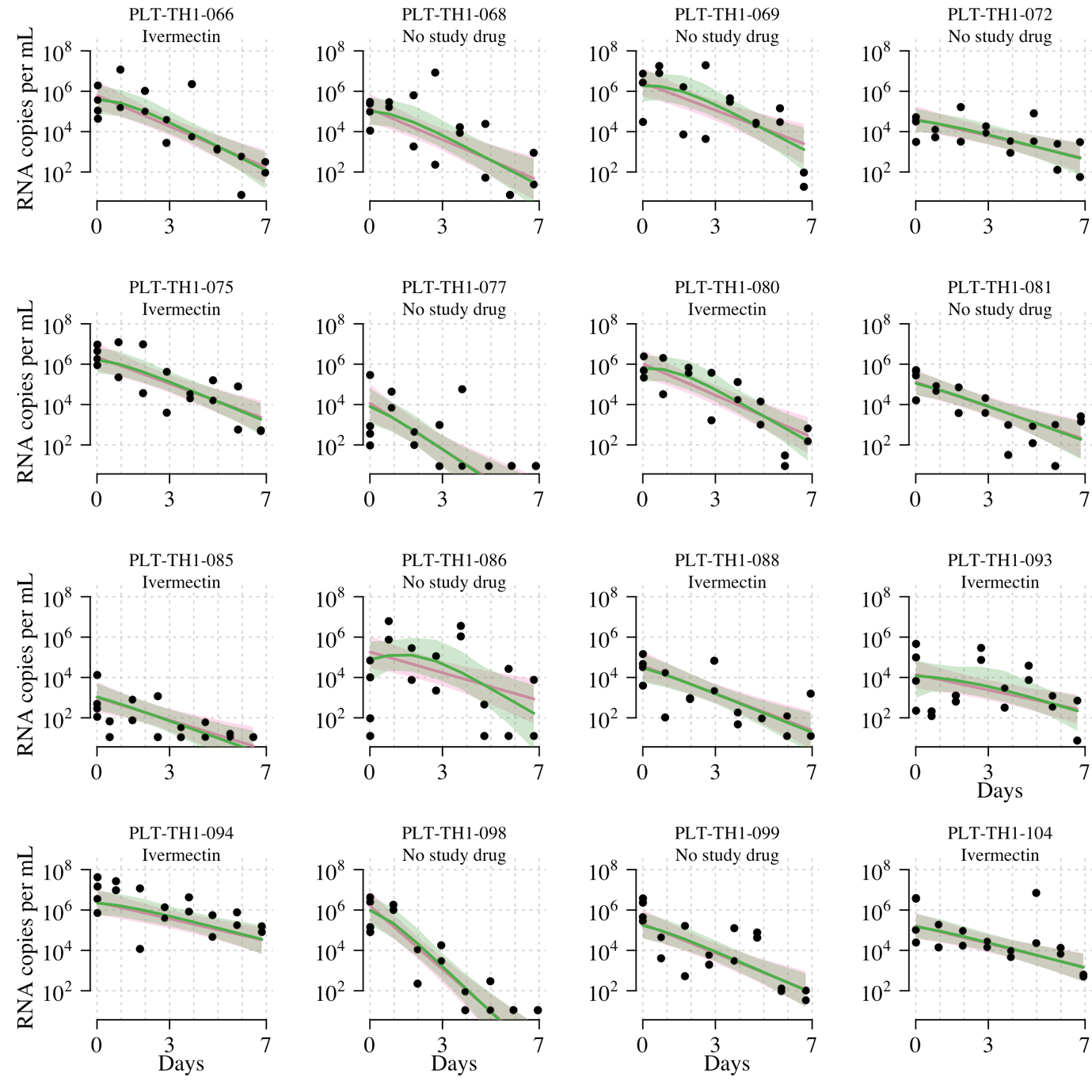

*SFigure 7 cont.*

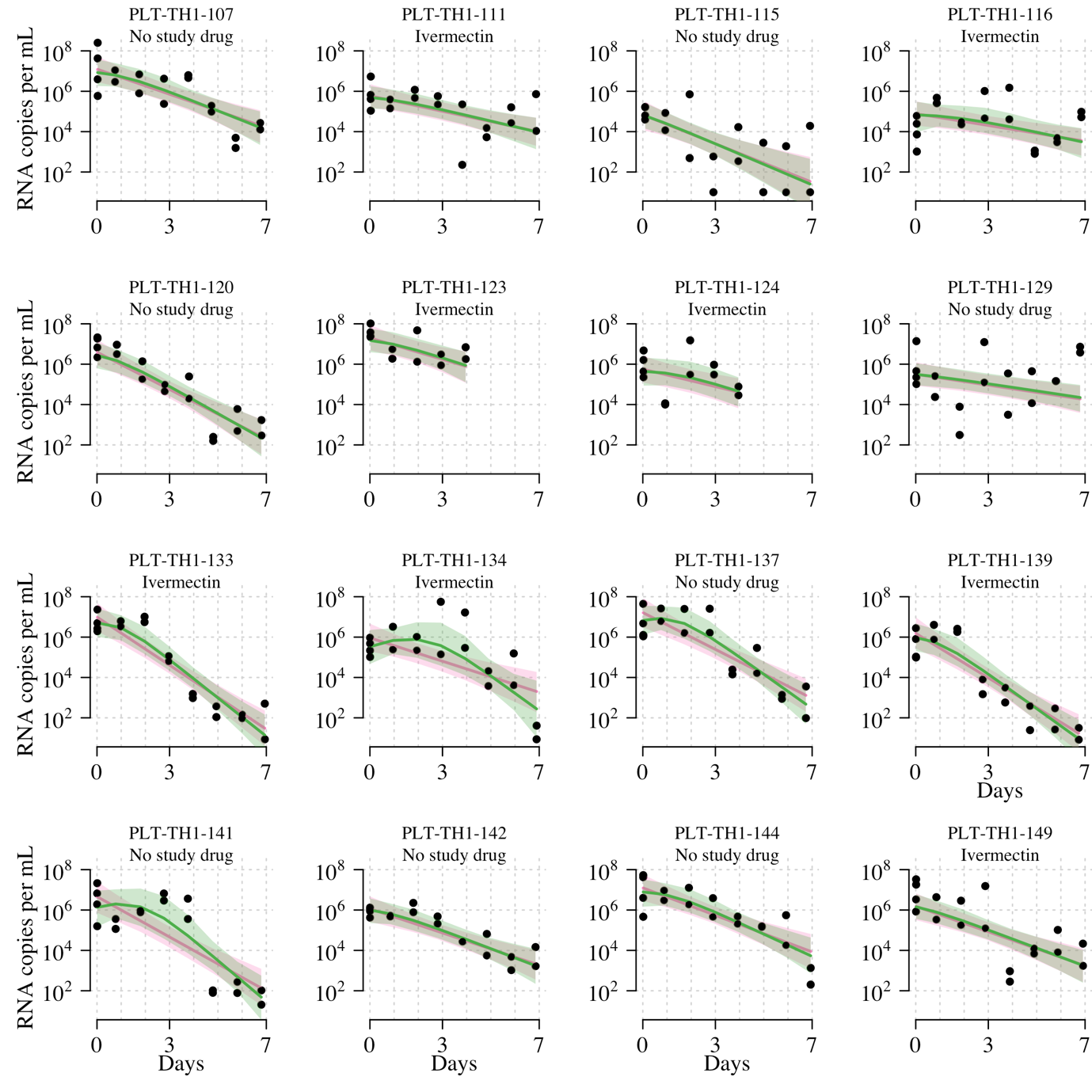

*SFigure 7 cont.*

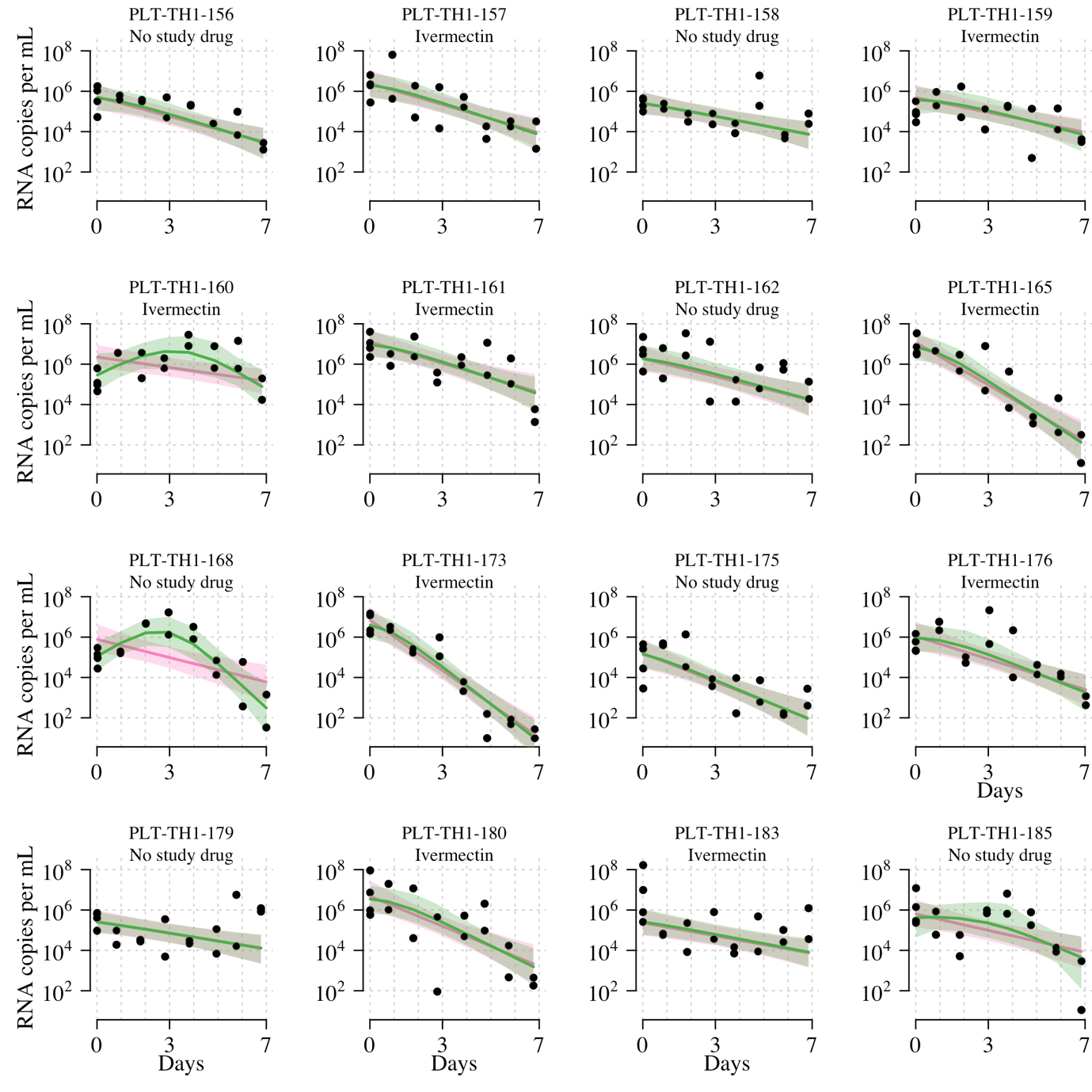

*SFigure 7 cont.*

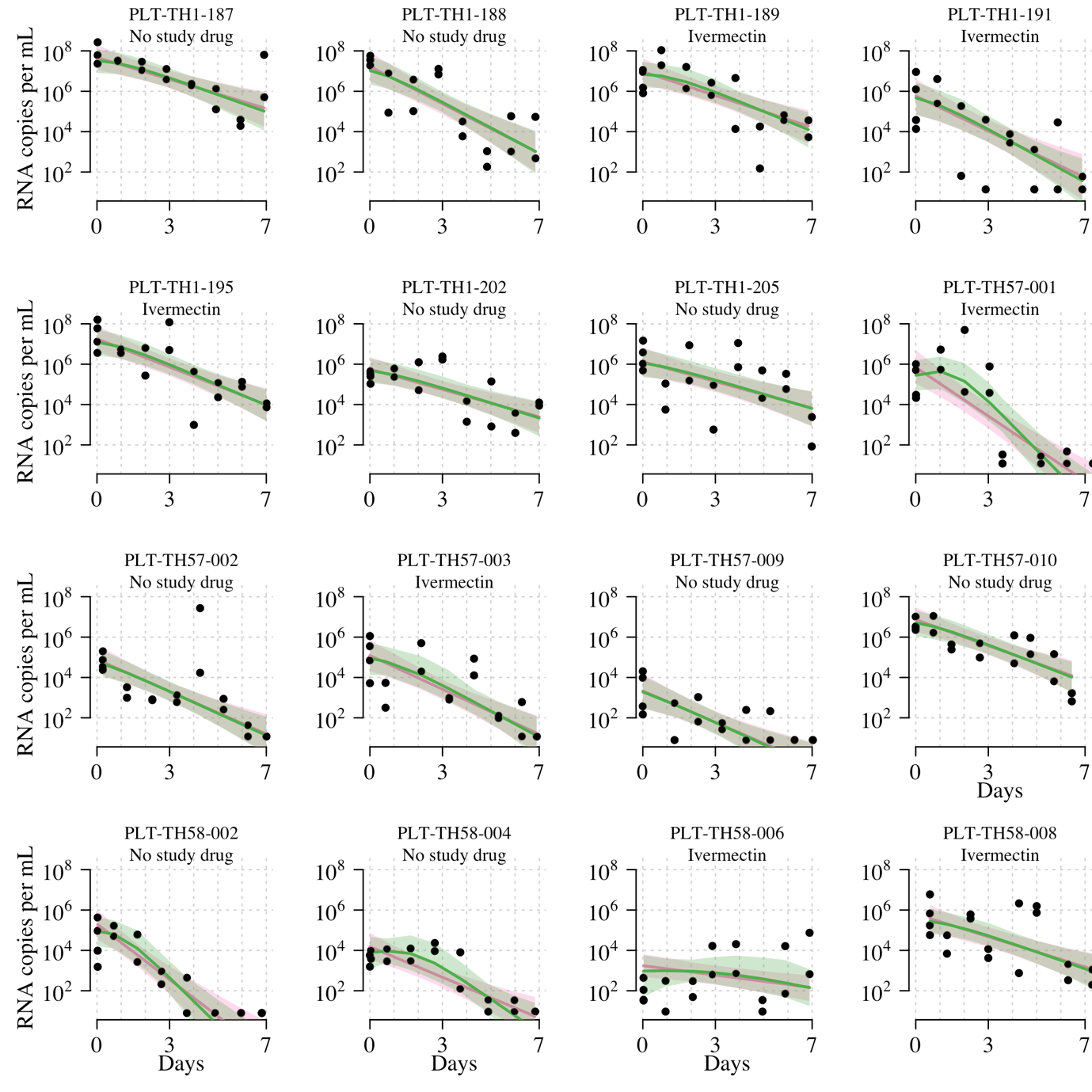

*SFigure 7 cont.*

**References:**

1. Navarro M, Camprubí D, Requena-Méndez A, Buonfrate D, Giorli G, Kamgno J, et al. Safety of high-dose ivermectin: a systematic review and meta-analysis. Journal of Antimicrobial Chemotherapy. 2020;75(4):827-34.

2. Smit MR, Ochomo EO, Aljayyoussi G, Kwambai TK, Abong'o BO, Chen T, et al. Safety and mosquitocidal efficacy of high-dose ivermectin when co-administered with dihydroartemisinin-piperaquine in Kenyan adults with uncomplicated malaria (IVERMAL): a randomised, double-blind, placebo-controlled trial. The Lancet Infectious Diseases. 2018;18(6):615-26.**3.**

3. Lange, K. L., Roderick J. A. Little, & Jeremy M. G. Taylor. (1989). Robust Statistical Modeling Using the t Distribution. *Journal of the American Statistical Association*, *84*(408), 881–896. https://doi.org/10.2307/2290063

4. Kobylinski KC, Jittamala P, Hanboonkunupakarn B, Pukrittayakamee S, Pantuwatana K, Phasomkusolsil S, Davidson SA, Winterberg M, Hoglund RM, Mukaka M, van der Pluijm RW, Dondorp A, Day NPJ, White NJ, Tarning J. Safety, Pharmacokinetics, and Mosquito-Lethal Effects of Ivermectin in Combination With Dihydroartemisinin-Piperaquine and Primaquine in Healthy Adult Thai Subjects. *Clin Pharmacol Ther*. 2020;107(5):1221-30.
